# PReCePT Devolved Nations Evaluation Report

**DOI:** 10.1101/2024.07.30.24311213

**Authors:** Hannah B Edwards, Carlos Sillero Rejon, Christalla Pithara-McKeown, Frank De Vocht, Hugh McLeod, Sabi Redwood, Liz Hill, Brent Opmeer, David Odd, Karen Luyt

## Abstract

This study set out to evaluate the longer-term sustainability, effectiveness, and cost-effectiveness of the National PReCePT Programme (NPP) in England, and explore trends and MgSO4 guidance implementation practices in the devolved nations, Scotland and Wales.

We found that the majority of improvement in MgSO4 use seemed to take place in the first year or two following the NPP. Benefits were largely sustained over the 4 years of follow-up, with an overall appearance of plateau in recent years. There was some indication of a slight declining trend in use coinciding with the COVID-19 pandemic, that continued to the end of 2022 (the end of the currently available data). Regional disparities in use of MgSO4 reduced since the NPP was launched.

We estimated that the NPP was associated with around £597,000 net monetary benefit (NMB) from a lifetime societal perspective, with an 89% probability of being cost-effective for babies with less than 30 weeks’ gestation. This NMB increased to £4.2m when including babies up to 32 weeks’ gestation.

By 2022, MgSO4 use in Wales had caught up with levels in England, with levels in Scotland not far behind. The NMB of implementing MgSO4 for babies up to 32 weeks’ gestation in the three nations has increased over time, generating approximately £125m in England, £8m in Scotland and £5m in Wales in 2022.

Consequently, the benefit forgone for not achieving optimal MgSO4 uptake has also reduced over time, although there remains considerable scope for improving performance in each nation. The improvements in implementing MgSO4 have generated health gains and cost savings associated with CP prevention. Investing additional resources in implementing MgSO4 further would be likely to be cost-effective in all three nations.

Our analysis highlighted how devolved nation activities were (directly or indirectly) shaped by PReCePT methodology. Qualitative interviews with clinical leads involved in implementing MgSO4 in Scotland and Wales – where the NPP was not implemented – shed light on the separate but similar initiatives implemented there, explaining the increasing trends also observed in the devolved nations (e.g. the Maternity and Children Quality Improvement Collaborative (MCQIC) Preterm Perinatal Wellbeing Package (PPWP) in Scotland, improvement interventions mirroring PERIPrem in Wales, and British Association for Perinatal Medicine Toolkits in both nations).

Challenges and enablers were linked to perinatal team relationships; local leadership with protected time and funding; access to national performance data; staff clarity and confidence on guidance and administration of treatment; opportunities for and commitment to co-creating meaning around the intervention; skills, competencies and resources available to adopters; and engagement in continuous improvement activities (e.g. audit and feedback, benchmarking and missed case reviews). Findings reiterate the need for local champions with backfill funding and protected time, and regional and national capacity building and support structures. These reflect findings from the corresponding interviews with English teams.

The essential next step in this quality improvement journey is to better quantify, in this same population, the health and societal benefits associated with cases of cerebral palsy prevented from the improvements achieved in use of MgSO4.

## 1. Background

Since 2015 the World Health Organisation (WHO)^1^ and the UK National Institute for Health and Care Excellence (NICE)^2^ have recommended administration of magnesium sulphate (MgSO4) in preterm deliveries <30 weeks’ gestation, with the option to extend up to 34 weeks’ gestation as a core part of maternity care. This follows strong evidence that when given antenatally to women in preterm labour, MgSO4 reduces the risk of cerebral palsy (CP) in preterm babies by around 30% (relative risk 0.68, 95% CI 0.54 to 0.87, derived from individual participant data meta-analysis (5 trials, 4601 babies))^3^. Yet by 2017 only 64% of eligible women in England received it, with high regional variation in uptake indicating inequalities in perinatal care^4^.

The cost of these preventable cases of CP is high. As well as the significant impact on affected individuals and their families^5^, CP is associated with lifetime societal costs around €800,000 per affected individual (2006 prices)^6^. On top of this, the NHS spends £1.8 billion annually on clinical negligence litigation for avoidable CP due to newborn brain injury. These cases account for half of the total NHS litigation expenditure, on average costing £10 million per case^7^.

MgSO4 is likely to be a highly cost-effective intervention. The cost of the drug is around £1, with associated costs (staff, consumables and monitoring) of administering it around £340. For births <30 weeks gestation, it is estimated that one case of CP can be prevented for every 37 mothers treated (NNT). We can estimate from other literature that in England, around 200 cases of CP per year could be avoided by consistent administration of MgSO4 during labour^3^.

In 2014-2015, a quality improvement framework and toolkit were developed and implemented in co-creation with five maternity units in the South-West of England^8^. Building on this successful pilot, in 2018, NHS England rolled out a scaled-up version of this intervention as the National PReCePT (Prevention of cerebral palsy in preterm labour) Programme (NPP). This was a quality improvement (QI) programme for maternity units, providing practical clinical guidance and learning resources, midwife backfill funding, and QI support to improve maternity staff awareness and increase use of MgSO4 for mothers in preterm labour. The aim was to reach ≥85% uptake across all maternity units in England, and the programme was delivered by regional Academic Health Science Networks (AHSNs).

Evaluation of the first 12 months of the programme found it to be both effective and cost-effective^9^. However, it is unknown whether the improvements achieved in the first year of the programme have been sustained over time, and experience from comparable international programmes suggests that initial gains may be lost in subsequent years. Sustained impact also implies that additional health benefits are realised without additional costs, thus further increasing the cost-effectiveness of the programme. The primary aim of this study was to evaluate the longer-term impact of the NPP in terms of effectiveness and cost-effectiveness (quantitative and health economic evaluation).

Another key aim was to compare MgSO4 use in England with that in the devolved nations of Scotland and Wales. Strategic and operational delivery of perinatal healthcare is not identical across the three nations, but there is much overlap. As England was exposed to the NPP and Scotland and Wales were not, this created the conditions for a “natural experiment” comparing performance between nations. In addition, in order to better understand how the devolved nations were responding to the NICE guidance, and whether the PReCePT programme in England may have, directly or indirectly, affected clinical practice in Scotland and Wales, this evaluation also aims to qualitatively assess explanatory mechanisms for differences in uptake of MgSO4 (or lack thereof) between England and the devolved nations (qualitative evaluation).

We aim to combine learning from the Devolved Nations study to that gained from the PReCePT evaluations to provide recommendations on implementation of improvement interventions for scale and spread of evidence-based perinatal interventions i.e. an implementation framework for future national programmes to accelerate getting research evidence into clinical practice. Increased understanding of the mechanisms behind the success of the PReCePT QI approach, compared to approaches adopted in the devolved nations, can help identify successful implementation strategies and practices, informing future practice.

## 2. Aims and objectives

### 2.1 Aims for the Quantitative evaluation

#### Primary aim

Estimate the sustained effectiveness of the NPP in improving MgSO4 use in preterm babies (<30 weeks gestation) in England, over the first four years since launch.

#### Secondary aims

- Compare MgSO4 use in England with that in the devolved nations Scotland and Wales.
- Explore the potential impact of the COVID-19 pandemic on MgSO4 use.
- Estimate the impact of the NPP on preterm babies <34 weeks gestation.
- Identify any difference in outcomes for the subgroup of maternity units that took part in the PReCePT RCT^10^.
- Identify sociodemographic factors associated with access to MgSO4 at the individual level.

### 2.2 Aims for the Health Economic evaluation

- Estimate the cost-effectiveness of the NPP in improving MgSO4 use in preterm babies (<30 and <32 weeks gestation) in England, over the four years since launch.
- Estimate the net monetary benefit of optimal MgSO4 implementation (assuming 95% of eligible mothers were treated) in the three nations.
- Estimate the potential impact of running NPPs in Scotland and Wales, based on the English NPP experience, in terms of effectiveness, implementation costs and cost-effectiveness.

### 2.3 Aims for the Qualitative evaluation

- Describe and explain enablers and barriers to implementing NICE guidance on MgSO4 use in the devolved nations.
- Describe and explain strategies adopted by those with responsibilities to promote administration of MgSO4 in response to the NICE clinical guidance in the devolved nations.
- Compare strategies, enablers and challenges experienced by the devolved nations with those emerging from the PReCePT NPP evaluation.

## 3. Methods

### 3.1 Quantitative evaluation

#### Study design

This was an observational study including a quasi-experimental or “natural experiment” design.

#### Population

The population of interest were NHS maternity units in England, Scotland and Wales, as the intervention was delivered at the maternity unit level. Within maternity units, analyses were performed on data on babies born preterm, before 30 weeks of completed gestation, and subsequently admitted to an NHS neonatal unit. In a secondary analysis, this population was extended to preterm babies up to 34 weeks.

#### Data source

De-Identified pseudonymised data were provided by the National Neonatal Research Database (NNRD) which holds routinely collected patient data on babies admitted to an NHS neonatal unit.

#### Sample size and power calculation

All maternity units in England, Scotland and Wales were included, except the five units in England that took part in the original PReCePT pilot study^8^ which were therefore not part of the subsequent NPP. For the primary analysis of sustained effectiveness on English data (predicted n=150 maternity units) it was estimated that there would be 91.7% power to detect an effect size of 6.3 percentage points (with a standard error of 0.0187) difference in uptake from pre- to post-implementation (effect estimates from the original study, assuming the effect had sustained). The minimum effect size detectable with 80% power would be 5.3 percentage points difference.

#### Statistical analysis

Population characteristics, MgSO4 use by nation and time-period, and reasons MgSO4 was not given were described.

For consistency with nationally reported data, MgSO4 uptake was defined as the number of mothers recorded as receiving MgSO4 divided by the total number of eligible mothers, excluding missing values from the denominator. This was computed per month per unit, and reported as a percentage. Only data on singletons and the first born (i.e. one infant) from each multiple birth were included in the calculation.

The primary effectiveness analysis was a multivariable linear regression model to estimate the difference in mean MgSO4 uptake from before (the 1 year period before) to after (the 4 years follow-up) implementation of the NPP in England. This difference was calculated both across the four years follow-up overall, and cumulatively by year of follow-up. The model adjusted for the underlying time trend expressed as number of months before or after implementation, and mother and baby characteristics aggregated nationally per month (mean maternal age, reported smokers, white British ethnicity, Index of Multiple Deprivation (IMD) decile, type of birth (c-section versus vaginal delivery), proportion of multiple births, baby’s birthweight adjusted for gestational age as a z-score). It also adjusted for the slope change, to account for the ceiling effect and reduction in rate of change at high levels. The model was weighted on the number of eligible births per unit per month. Data on paternal age and ethnicity were explored as potential confounding factors, but were excluded due to high levels of missing data and expected collinearity with maternal age and ethnicity.

The main model was at the national-level. The original plan was to use data aggregated to the maternity-unit level, but regression diagnostics showed that nationally-aggregated data resulted in a superior model fit, largely because national averages have higher sample sizes per month, so are far less affected by extreme monthly proportions of 0% and 100%. Both maternity-unit-level and individual-level analyses were also performed as sensitivity analyses. The sensitivity analyses on individual-level and maternity-unit level data additionally adjusted for level of maternity unit (Neonatal Intensive Care Unit (NICU) versus Special Care Baby Unit (SCBU) or Local Neonatal Unit (LNU)), and regional clustering by AHSN. Potential interaction was explored between MgSO4 uptake and level of maternity unit (NICU versus SCBU or LNU), as performance could plausibly differ by unit type.

A secondary analysis used a Bayesian structural time-series model to evaluate the impact of the NPP. This method was originally developed to evaluate the impact of marketing campaigns^11^. It compares the observed uptake trend in England post-NPP, with a counterfactual trend (what might have happened without the NPP) based on a synthetic control group of weighted Scottish and Welsh data. The difference between the observed and counterfactually expected trends in the post-NPP period can be interpreted as the causal impact of the intervention. This method can reduce bias in natural experiment studies where there is no randomization of exposure^11–13^. For this model, the national NPP launch date (May 2018) was taken as the intervention start date to approximate an ‘intention to treat’ analysis. Post-hoc sensitivity analyses with transformations and different distributional assumptions were performed on this model to optimise the fit, due to the change in rate of change near the ceiling.

Logistic regression modelling was used for individual-level analysis of sociodemographic predictors of MgSO4 receipt. Maternal age, ethnicity, index of multiple deprivation (IMD) decile, North vs South of England, and history of smoking were included in the model. The odds of receiving treatment were described for the four years pre-PReCePT (2014-2018) and the four years post-PReCePT (2018-2022).

Statistical software Stata version 17 and R version 4.3.1 were used for all statistical analyses.

### 3.2 Health Economics

#### 3.2.1 Economic Evaluation of the National PReCePT Programme

##### 3.2.1.1 Data and assumptions for the economic evaluation

Data and assumptions used for the economic evaluation of the NPP are described in the following sections.

###### Cost-effectiveness of MgSO4 treatment

There is evidence of the cost-effectiveness of MgSO4 to prevent CP and neurodisabilities^14–16^. We adopted Bickford and colleagues’ results on treatment cost-effectiveness, converting their estimated costs to GBP currency and 2019 prices (see Appendix 1)^14^. We used a combined estimate with lifetime incremental savings of £19,054 and incremental QALYs of 0.24 for babies with less than 32 weeks gestation.

###### Cost of the National PReCePT Programme

The mean implementation cost per unit of the NPP was estimated from data supplied by the national programme team and the PReCePT Study team and reported previously^9^ ^17^. NPP costs included those relating to national programme management, funding support from AHSNs, and the funded backfill of clinical time for midwives. On average, the NPP costed £6,044 per maternity unit. The total cost of the NPP at a national level was estimated at £936,747.

###### Effectiveness of the National PReCePT Programme

We estimated the NPP effectiveness as the difference between the predicted level of MgSO4 use over time compared to a counterfactual level of MgSO4 use representing what may have occurred in the absence of the NPP. The predicted level of MgSO4 use over time was obtained from the multivariable linear regression for the difference in mean MgSO4 uptake from before (the one-year before) to after (the four-year follow-up) implementation of the NPP in England. The counterfactual assumes a continuation of the pre-NPP trend in MgSO4 uptake. Therefore, our main measure of NPP effectiveness was the area-between-the-curves between the predicted MgSO4 uptake and the counterfactual uptake.

Our main analysis uses a linear distribution to estimate the counterfactual based on the pre-NPP predicted trend. In a sensitivity analysis, we used a beta distribution to estimate this counterfactual to account for both an upper and lower bound for MgSO4 uptake as a proportion. Uptake is limited to a maximum of 100% and a diminishing rate of increase in uptake.

Our main analysis includes the change in the proportion of patients treated with babies of less than 30 weeks gestation. Secondary analyses included babies of less than 32 weeks gestation to align with the cost-effectiveness evidence of MgSO414-16, and babies of between 32 and 34 weeks gestation.

###### Population data

The size of the eligible population was based on anonymised patient-level data from the UK NNRD as the number of preterm babies at a national level for the periods of interest. The eligible population was calculated monthly for babies of less than 30 weeks gestation, babies of less than 32 weeks gestation and babies of between 32 and 34 weeks gestation.

###### Perspective and time horizons

MgSO4 has shown to be a cost-effective treatment in the prevention of CP in preterm births from a societal perspective and lifetime perspective^14–16^. We calculated the evidence of the implementation from a health care perspective. Therefore, the economic evaluation will account for the health gains and cost-savings from a societal perspective and lifetime perspective.

###### Cost-effectiveness thresholds

We use a threshold of £20,000 per QALY, following NICE guidelines^18^.

##### 3.2.1.2 Analysis

###### Policy cost-effectiveness analysis

We estimated the policy cost-effectiveness of the NPP over the four years since its launch by combining analysis of the costs and effectiveness of the NPP to increase MgSO4 implementation with the lifetime societal cost and health gains of MgSO4 treatment. This analysis used a framework we previously developed to conduct economic evaluations of implementation initiatives^19^:

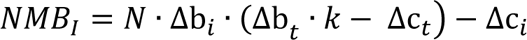

Where NMBI is the net monetary benefit (NMB) of the implementation initiative (i.e., NPP) at a willingness-to-pay-threshold (k). A positive net monetary benefit will indicate that the implementation initiative was cost-effective. N is the size of the eligible population obtained from the population data. Δbi is the estimate of the NPP effectiveness as described above. Δci is the estimated cost of the NPP as described above. Δct and

Δbt are the net cost and health gains of MgSO4 treatment per patient estimated from previous work on the cost-effectiveness of MgSO4 for the prevention of CP.^14^

We calculated the net increment of the number of patients that received MgSO4 (*Δbi*\**N*), the implementation cost-effectiveness per additional patient treated (*Δci/(Δbi*\**N)*), and the NMB of the NPP.

These statistics were calculated for each of the measures of effects: the area-between-the-curves estimate with a linear counterfactual (main analysis) and a beta counterfactual (sensitivity analysis). A secondary analysis included babies less than 32 weeks gestation. We did not perform a policy cost-effectiveness analysis for the sub-group analysis accounting for babies between 32 and 34 weeks gestation as there is no available literature reporting evidence of the lifetime cost-effectiveness of MgSO4 treatment for these patients.

###### Sampling uncertainty

We conducted a probabilistic analysis using Monte Carlo simulation with 10,000 samples drawn from parameter distributions. Point estimates, probabilistic distribution assumptions, and parameter source estimates are reported in Appendix 1. We plotted cost-effectiveness planes and cost-effectiveness acceptability curves for willingness-to-pay thresholds from zero to £100,000 per QALY gained for the policy cost-effectiveness of the NPP intervention.

###### Statistical software

All data management and analyses will be performed in Stata version 18 and R.

#### 3.2.2 Net monetary benefit of optimal MgSO4 treatment

##### 3.2.2.1 Data and assumptions

Data and assumptions used for estimating the net monetary benefit of optimal implementation of MgSO4 treatment are described in the following sections.

###### Population data

We calculated the monthly number of pre-term babies from January 2014 to December 2022 based on anonymised patient-level data from the UK NNRD for each nation (England, Scotland and Wales). We also calculated the number of babies treated with MgSO4 as the product between the monthly MgSO4 uptake and the monthly number of pre-term babies.

###### Cost-effectiveness of MgSO4 treatment

As noted above, we adopted Bickford and colleagues’ results on treatment cost-effectiveness, converting their estimated costs to GBP currency and 2019 prices (see Appendix 1). We used a combined estimate with lifetime incremental savings of £19,054 and incremental QALYs of 0.24 for babies with less than 32 weeks gestation.

###### Willingness-to-pay thresholds

We use a threshold of £20,000 per QALY, following NICE guidelines^18^.

##### 3.2.2.2 Analysis

###### Net monetary benefit of optimal MgSO4 implementation analysis

Using the framework described above we estimated the following statistics for each nation:

a. The cumulative lifetime NMB of MgSO4 treatment for all the patients each month.
b. The cumulative lifetime NMB of optimal MgSO4 treatment for all the patients each month, assuming that optimal implementation would be achieved if 95% of the babies were treated at the time. This level of optimal uptake was determined by the clinicians in the research group and reflects uptake of MgSO4 which could be achievable and sustained over time.
c. The NMB not generated due to MgSO4 not being implemented optimally, which is the difference between the NMB of the current implementation (a) and the NMB of optimal implementation (b).

We estimated these values monthly from January 2014 to December 2022. We plotted these results to study the trajectories of these three statistics. We also calculated these values for the first year of the observed data (2014) and the last year (2022).

###### The cost-effectiveness of future implementation analysis

The NMB not generated due to MgSO4 not being implemented optimally indicates the ‘value of perfect implementation’, or how much could have been invested to achieve the optimal uptake of MgSO4. For this reason, we estimated the NMB of optimal implementation for a year for the three nations as the NMB not generated in 2022. This will inform an upper threshold of how much could be invested in implementation to achieve optimal uptake of MgSO4 in a year and be viewed as good value for money.

We also calculated the potential cost-effectiveness for different implementation programme scenarios with different implementation effectiveness (increment proportion of patients treated) and different implementation costs for a year for the three nations. We plotted these results in heatmaps to visualise the cost-effectiveness of hypothetical scenarios with different levels of implementation cost and implementation effectiveness.

We focused on three scenarios depending on the implementation effectiveness: low performance (1% increment of MgSO4 uptake), mid-performance (5%), and high performance (10%). For these three scenarios, the implementation costs were assumed to be similar to the NPP in England: £6,044 per unit. Therefore the costs were assumed £936,747 for England, £84,609 for Scotland and £54,392 for Wales. We also accounted for the uncertainties of the MgSO4 cost-effectiveness including the 5^th^ and 95^th^ percentiles from the probabilistic analysis.

Positive NMB indicate that the implementation would be cost-effective and, therefore, good value for money to achieve the assumed level of implementation effectiveness.

###### Subgroup analyses

Analysis was calculated for two groups of patients: babies with less than 30 weeks gestation, and babies between 30 and 32 weeks gestation. We also run the analysis for the total number of babies (<32 weeks gestation). We also report the uptake at Operational Delivery Network (ODN) level in England to explore the differences in performance between networks.

### 3.3 Qualitative evaluation

To understand the strategies, implementation processes and factors affecting implementation in the two devolved nations, we carried out semi-structured, remote (MS Teams) interviews with key staff working in perinatal strategic and implementation lead roles i.e. individuals involved in the design, planning and/or implementation of national and/or regional programmes and initiatives targeting maternity and neonatal safety in general, and of MgSO4 for neuroprotection specifically. Recruitment was guided by the concept of information power^20^ which proposes that the number of participants recruited to the study is guided by the richness of responses and research participants’ depth of knowledge specific to the topic of interest.

The topic guide (Appendix 3) was informed by the Normalisation Process Theory, a theoretical framework which helps us understand the dynamics of implementing, embedding and integrating a complex intervention such as quality improvement and perinatal optimisation interventions including magnesium sulphate in routine practice ^21^. Our previous evaluation of the National PReCePT Programme and the PReCePT study described how the mechanisms described by the theory aligned with the Quality Improvement methodology used by PReCePT QI. Appendix 4 illustrates how the four mechanisms overlap with the four primary drivers of PReCePT QI as presented in the PReCePT Implementation Guide. By using the same framework, we can compare what has taken place in the two nations to PReCePT activities in relation to the four implementation mechanisms.

The interviews provided data on the interventions set up in the two devolved nations, and shed light on:

1. The strategies adopted to communicate and create understanding among adopters of the intervention (coherence),
2. which people were involved in implementation and how they were brought together to discuss and make plans around the intervention (cognitive participation),
3. strategies adopted to make it possible for people to administer the intervention and make it part of everyday practices and routines (collective action), and
4. the ways in which actors monitored and appraised the intervention and used data for improvement (reflexive action).

All potential participants were approached by steering group members in the first instance, and later on other participants (snowball sampling). Individuals were introduced to the study and its aims, and provided with a Participant Information Sheet and invitation letter. Potential participants were given the researcher’s (CPM) contact information and approached for further information and making arrangements for an interview. Information on the voluntary and anonymous nature of participation, audio-recording and transcribing the interview and including data in an open research repository was provided in the PIS. All participants provided verbal consent before taking part in the interview.

Data collection took place between November 2022 and July 2023 and lasted between 28 and 58 minutes. All interviews were audio recorded and fully transcribed by a University of Bristol approved transcription company.

Transcripts were imported into NVIVO QSR, qualitative data management software, for analysis. Analysis used the framework method (Gale, Heath et al. 2013). Our analysis framework matrix was informed by the Normalisation Process Theory coding manual (May, Albers et al. 2022). Two researchers, one member of the research team and one independent member of the wider ARC research team, independently coded three transcripts and met to discuss their coding and resolve any discrepancies in the coding of the data. Coding of transcripts was initially deductive based on the NPT coding manual, followed by inductive analysis to identify themes emerging from the participants’ interviews. No themes distinct from the NPT were identified. Interim findings were also regularly presented to the steering group and wider team to ensure accuracy of interpretation and internal consistency of codes. A framework matrix including rows (participants), columns (themes) and “cells” of summarised data enabled comparative analysis of data across, as well as within, cases to inform an understanding of factors affecting implementation.

## 4. Results

### 4.1 Quantitative evaluation

#### 4.1.1 Baseline characteristics

Across January to December 2017, the year before NPP roll-out, a total of 4091 babies under 30 weeks gestational age were admitted to neonatal units in England, 296 in Scotland, and 182 in Wales. The majority of the births were in maternity units with a NICU. In total there were 150 maternity units in England (3 with no neonatal service, 106 with an associated SCBU/LNU, 41 with an associated NICU); 18 maternity units in Scotland (3 with no neonatal service, 6 with a SCBU/LNU, 9 with a NICU); and 12 maternity units in Wales (2 with no neonatal service, 7 with a SCBU/LNU, 3 with a NICU). The mean number of eligible births per hospital per month was 2.8 in England, 2.3 in Scotland, and 2.1 in Wales.

Other than in the number of babies admitted, study populations were largely comparable across the three nations. Differences included: Wales had a lower proportion of the extremely preterm babies <25 weeks, Scotland and Wales both had a higher proportion of mothers of white British ethnicity compared to England; Wales had a higher proportion of mothers with a history of smoking; and Scotland had a higher proportion of caesarean section births. (Table 1)

**Table 1:**
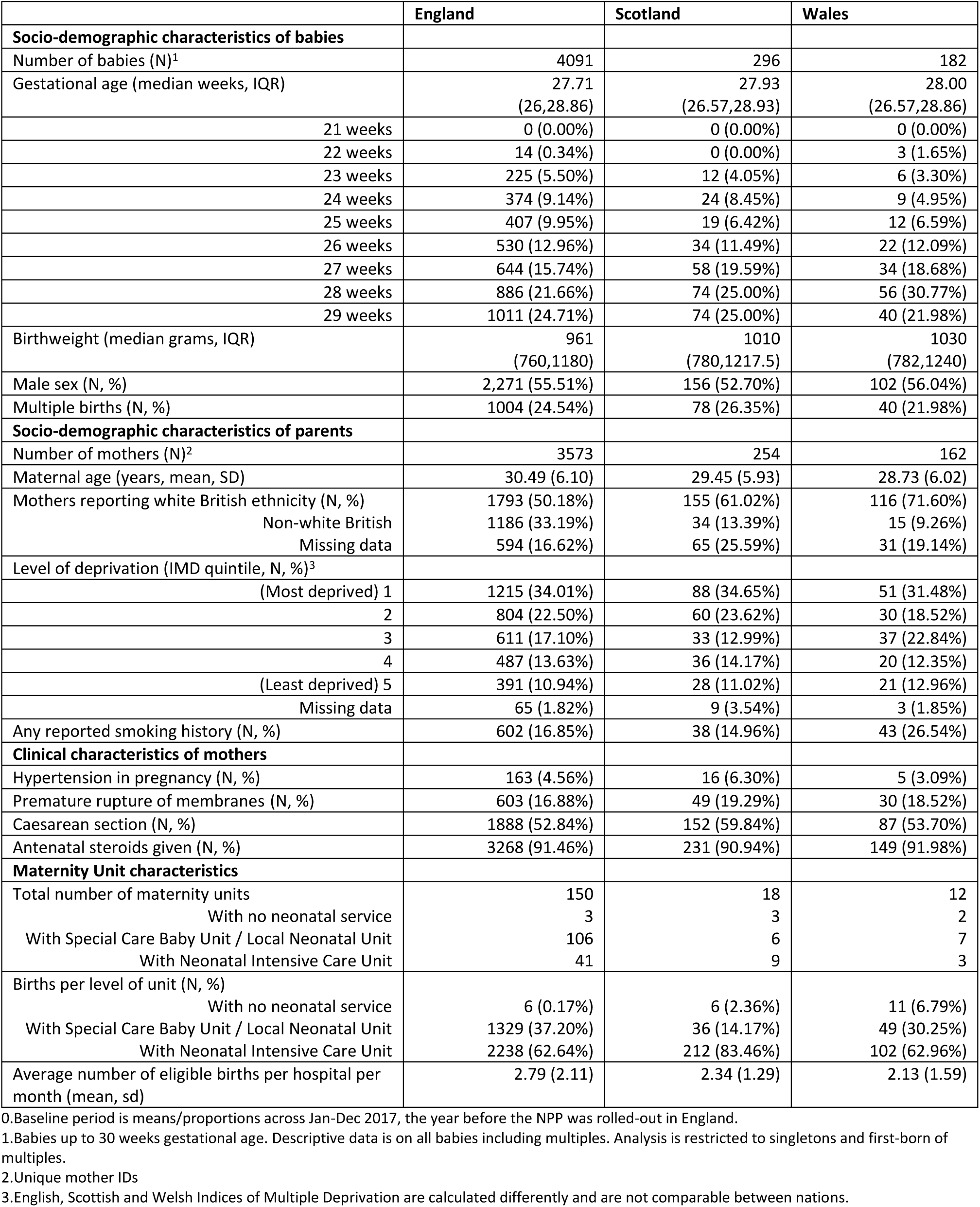
Baby, mother, and maternity unit characteristics by nation at baseline^0^.

#### 4.1.2 Historical trends

MgSO4 reporting in the NNRD started to become reliable around 2014, and at that point uptake (proportion of eligible mothers receiving treatment) was recorded at around 20% in England, 40% in Scotland, and 10% in Wales. The three nations appeared to follow broadly comparable increasing trends in uptake, with improvements becoming more gradual as the proportion receiving MgSO4 approached high levels (a natural ceiling effect). There was visual suggestion that post-NPP, uptake may have improved more rapidly in England compared to the devolved nations. However, due to relatively small numbers, there was high variation in monthly uptake for Scotland and Wales, which limits formal comparison of trends. (Figure 1)

**Figure 1:**
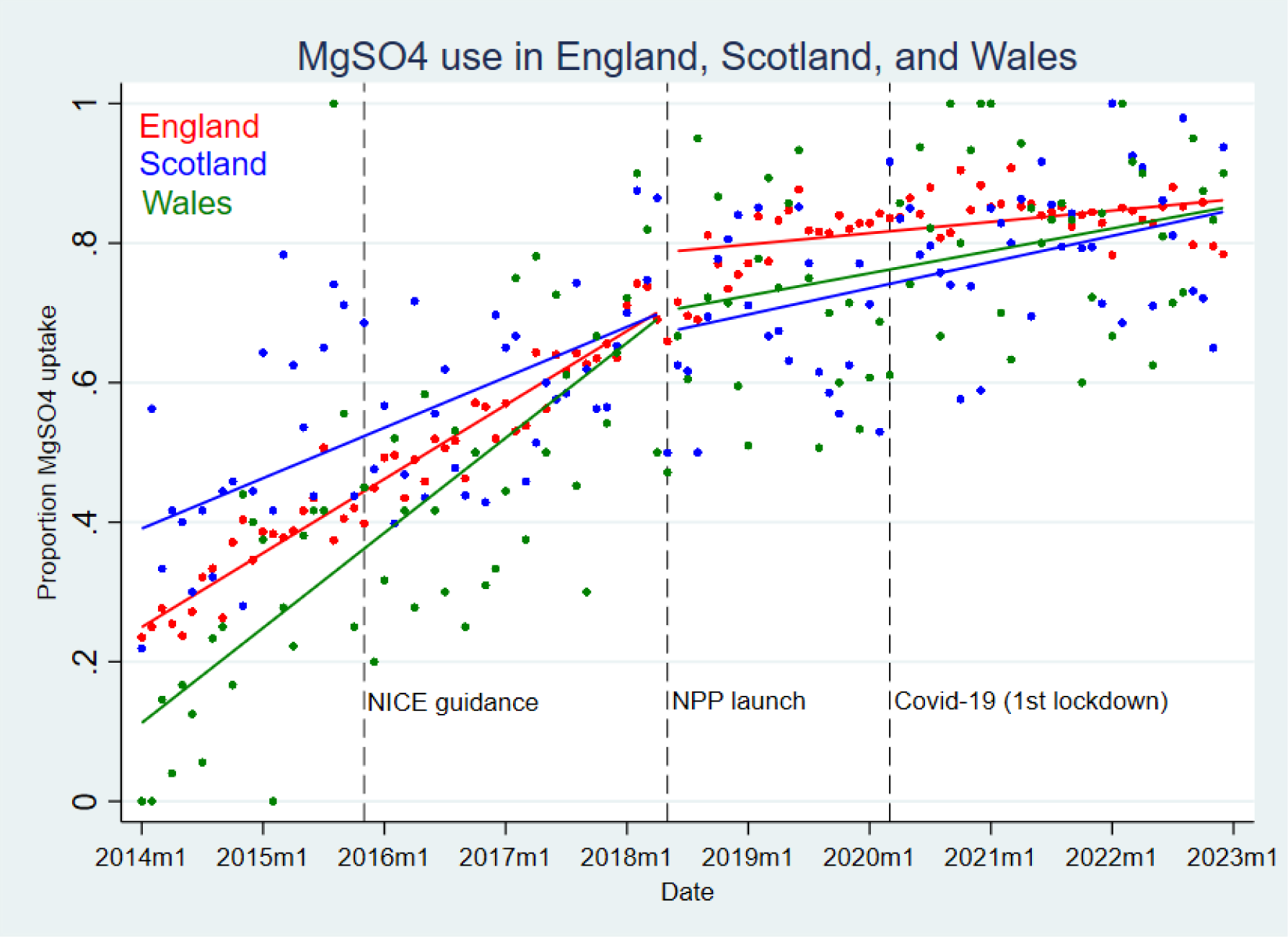
MgSO4 uptake in England, Scotland and Wales, 2014 to 2022

#### 4.1.3 Overall trends in uptake pre- and post-NPP

In England, overall MgSO4 uptake rose from 65.8% in 2017 to 85.5% in 2022. The corresponding increases in the devolved nations were from 62.3% to 81.4% in Scotland, and from 61.6% to 86.6% in Wales. The amount of missing data fell from around 5% in 2017 to under 1% in 2022. Imminent delivery was the most commonly recorded reason for not giving MgSO4 at both time points: this accounted for around 15% of eligible babies in 2017, going down to around 10 percentage points in England and Wales in 2022. The number recorded as not offered MgSO4 fell from around 7% to around 1% across all three nations. (Table 2, Figure 2)

**Figure 2:**
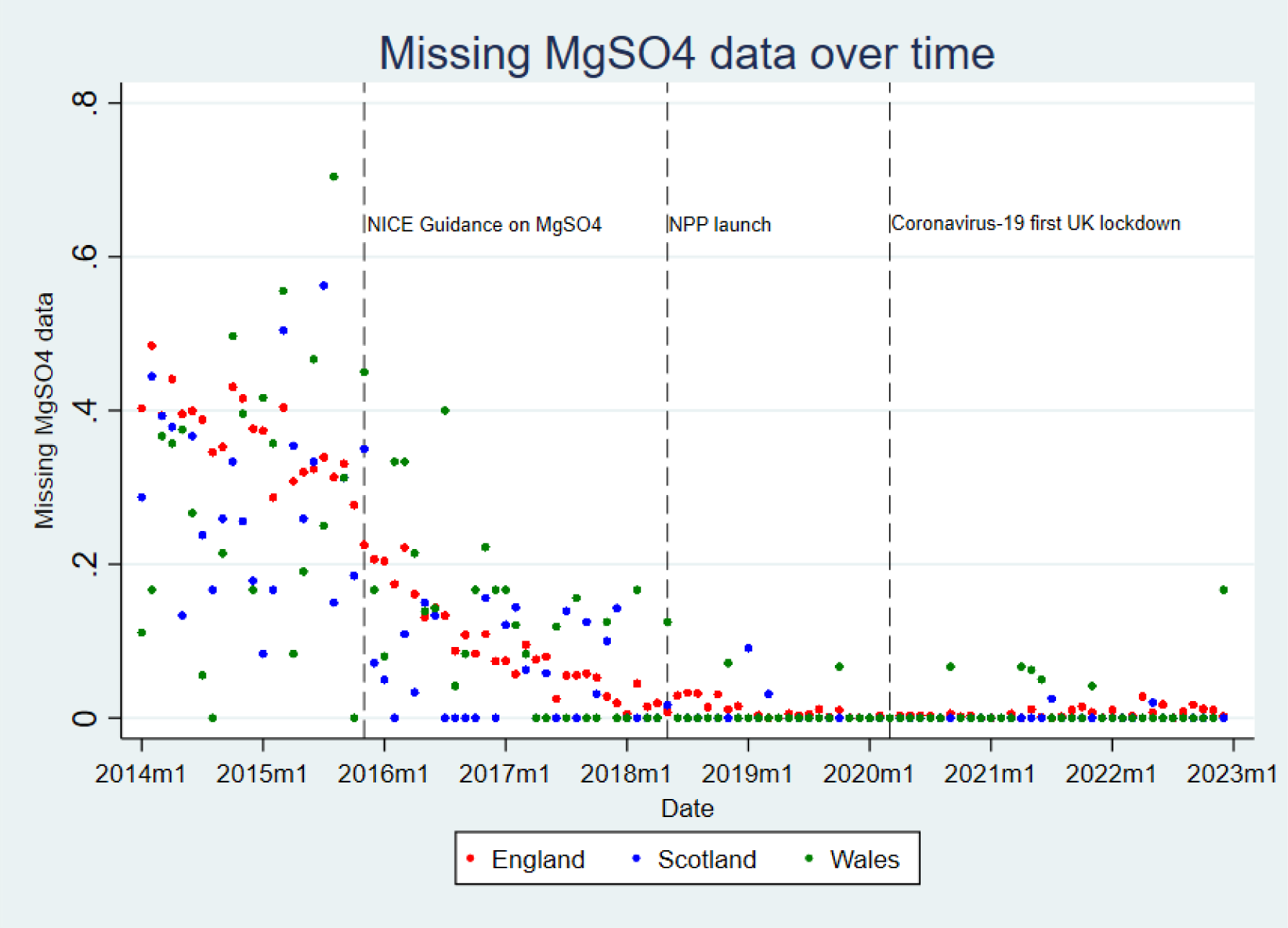
Missing MgSO4 data in England, Scotland and Wales, 2014 to 2022

**Table 2:** MgSO4 uptake in England, Scotland, and Wales, pre- and post-NPP^0^.

|  | England |  | Scotland |  | Wales |  |
| --- | --- | --- | --- | --- | --- | --- |
|  | 2017<sup>1</sup> | 2022<sup>2</sup> | 2017<sup>1</sup> | 2022<sup>2</sup> | 2017<sup>1</sup> | 2022<sup>2</sup> |
| Total number of eligible births<sup>0</sup> | 3573 | 3286 | 254 | 253 | 162 | 135 |
| Total number of mothers **given** MgSO<sub>4</sub> (%) | 2223<br>(62.22%) | 2786<br>(84.78%) | 149<br>(58.66%) | 205<br>(81.03%) | 93<br>(57.41%) | 116<br>(85.93%) |
| Total number of mothers **not given** MgSO<sub>4</sub> (%) | 1158<br>(32.41%) | 474<br>(14.42%) | 90<br>(35.57%) | 47<br>(18.58%) | 58<br>(35.80%) | 18<br>(13.33%) |
| Total number with **missing** MgSO<sub>4</sub> data (%) | 192<br>(5.37%) | 26<br>(0.79%) | 15<br>(5.91%) | 1<br>(0.40%) | 11<br>(6.79%) | 1<br>(0.74%) |
| **MgSO<sub>4</sub> uptake<sup>3</sup>** (sd) | 65.75%<br>(0.47) | 85.46%<br>(0.35) | 62.34%<br>(0.49) | 81.35%<br>(0.39) | 61.59%<br>(0.49) | 86.57%<br>(0.34) |
| Reason MgSO<sub>4</sub> not given (%)<sup>4</sup> |  |  |  |  |  |  |
| Contraindicated | 11 (0.31%) | 5 (0.15%) | 2 (0.79%) | 1 (0.40%) | 0 (0.0%) | 1 (0.74%) |
| Declined | 3 (0.08%) | 3 (0.09%) | 0 (0.00%) | 1 (0.40%) | 1 (0.62%) | 0 (0.00%) |
| Delivery imminent | 548 (15.34%) | 329 (10.01%) | 42 (16.54%) | 38 (15.02%) | 23 (14.20%) | 13 (9.63%) |
| Not appropriate | 126 (3.53%) | 23 (0.70%) | 14 (5.51%) | 1 (0.40%) | 12 (7.41%) | 0 (0.00%) |
| Not offered | 240 (6.72%) | 32 (0.97%) | 18 (7.09%) | 2 (0.79%) | 14 (8.64%) | 2 (1.48%) |
| Data missing | 230 (6.44%) | 82 (2.50%) | 14 (5.51%) | 4 (1.58%) | 8 (4.94%) | 2 (1.48%) |

0. All data on singletons and first born of multiples <30 weeks gestation and admitted to an NHS Neonatal unit
1. Across the year Jan-Dec 2017
2. Across the year Jan-Dec 2022
3. Uptake percentage calculated excluding missing values from the denominator, to fit with national audit reporting practices.
4. Percentage calculated out of total cases

#### 4.1.4 Estimate of improvement from before to after the NPP (main analysis)

The adjusted estimate was for an average 5.8. percentage point increase in MgSO4 uptake in England over the four years since the NPP launch, compared to the one year before the NPP (95% CI 2.7 to 8.9, p<0.001). The majority of the gains appeared to take place as a step-change in the first year or two of the programme. There was some indication of additional gains in years 3-4 (at which point the improvement became statistically significant), but confidence intervals overlap with estimates from the first two years, so the possibility of additional later gains is less clear. On average across the follow-up period, the post-intervention slope appeared to plateau (slope = 0.02, 95% CI -0.06 to 0.10, p=0.622).

These estimates were robust to sensitivity analyses (including: analysis at the maternity unit and individual level; excluding a ‘fuzzy’ implementation start window of +/-2 months; excluding the final 2 months of data (due to potential incomplete data from some units); and using a longer pre-NPP comparison period of 4 years). Main model diagnostics showed a good fit.

There was a suggestion of greater gains both in the 40 units in the PReCePT RCT, and when including babies up to 34 weeks gestational age. However, the confidence intervals of these estimates overlapped with those of the main result, so the evidence of additional benefit in these groups is unclear. There was some evidence that lower-level maternity units improved more than higher-level units (SCBUs and LNUs: 9.1 percentage points change, 95%CIs 3.9 to 14.3, p=0.001. NICUs: 4.1 percentage points change, 95%CIs 0.6 to 7.6, p=0.022. Test for interaction p=0.020). (Table 3, Figure 3)

**Figure 3:**
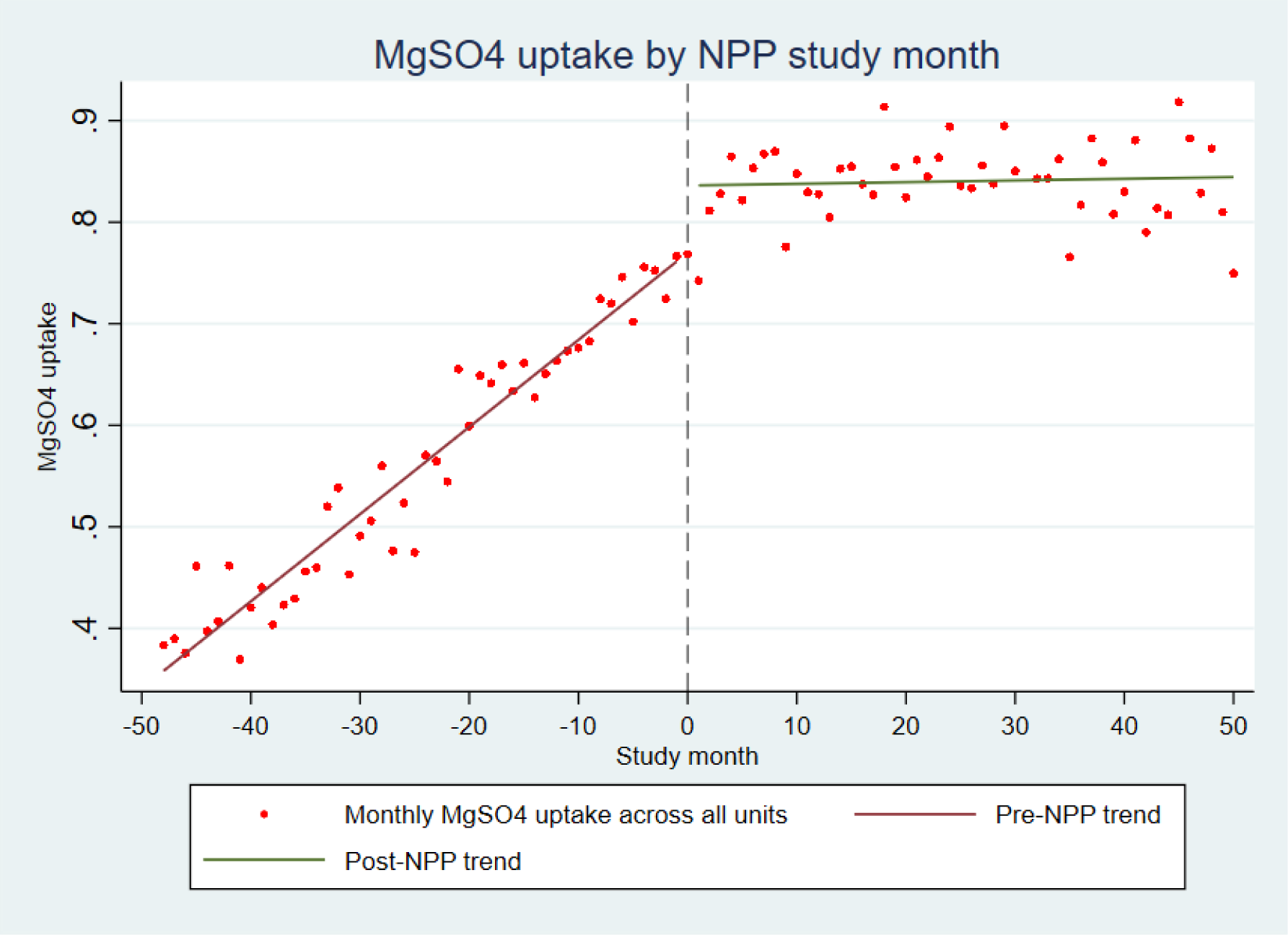
MgSO4 uptake in England by NPP study month

**Table 3:** Change in MgSO4 uptake in England from before to after the NPP^0^.

|  | Step-change in MgSO<sub>4</sub> uptake after intervention (percentage points)<sup>1</sup> | 95% CI | p-value | Change in slope post-intervention compared to pre-intervention | 95% CI | p-value | Overall slope post-intervention | 95% CI | p-value |
| --- | --- | --- | --- | --- | --- | --- | --- | --- | --- |
| Unadjusted<sup>2</sup> | 11.80 | 10.01 to 13.59 | <0.001 | - | - | - |  |  |  |
| Fully adjusted<sup>3</sup> | 5.77 | 2.69 to 8.86 | <0.001 | -0.87 | -1.18 to -0.57 | <0.001 | 0.02 | -0.06 to 0.10 | 0.622 |
| **Cumulative increase per-year across the follow-up period<sup>2</sup>** |  |  |  |  |  |  |  |  |  |
| Year 1 | 3.06 | -1.28 to 7.40 | 0.167 | -0.76 | -1.48 to -0.05 | 0.037 | 0.19 | -0.31 to 0.69 | 0.450 |
| Year 1-2 | 3.52 | -0.24 to 7.28 | 0.067 | -0.57 | -0.91 to -0.23 | <0.001 | 0.25 | 0.09 to 0.41 | 0.002 |
| Year 1-3 | 5.58 | 2.10 to 9.06 | 0.002 | -0.86 | -1.25 to -0.48 | <0.001 | 0.05 | -0.05 to 0.16 | 0.302 |
| Year 1-4 | 5.77 | 2.69 to 8.86 | <0.001 | -0.87 | -1.17 to -0.57 | <0.001 | 0.02 | -0.06 to 0.10 | 0.622 |
| **Secondary, sensitivity and sub-group analyses<sup>2</sup>** |  |  |  |  |  |  |  |  |  |
| Analysis at maternity-unit level<sup>4</sup> | 5.83 | 2.82 to 8.83 | <0.001 | -0.83 | -1.47 to -0.18 | 0.012 | 0.003 | -0.05 to 0.06 | 0.923 |
| Analysis at individual-level<sup>4</sup> | 5.91 | 2.50 to 9.31 | 0.001 | -0.76 | -1.17 to -0.34 | <0.001 | 0.02 | -0.03 to 0.07 | 0.449 |
| Excluding implementation start window (2 months each side of start) | 6.59 | 2.93 to 10.25 | <0.001 | -0.86 | -1.17 to -0.54 | <0.001 | -0.01 | -0.09 to 0.07 | 0.820 |
| Excluding final 2 months (?data quality) | 5.78 | 2.64 to 8.92 | <0.001 | -0.82 | -1.14 to -0.50 | <0.001 | 0.04 | -0.04 to 0.12 | 0.346 |
| Comparing 4 years pre with 4 years post | 6.03 | 3.70 to 8.36 | <0.001 | -0.74 | -0.82 to -0.65 | <0.001 | 0.01 | -0.07 to 0.08 | 0.895 |
| PRECePT cRCT units only (n=40) | 8.62 | 2.37 to 14.87 | 0.007 | -0.79 | -1.37 to -0.20 | <0.001 | 0.04 | -0.09 to 0.16 | 0.563 |
| Including babies up to 34 weeks gestation | 8.67 | 6.38 to 10.96 | <0.001 | -0.69 | -0.90 to -0.48 | <0.001 | 0.046 | -0.02 to 0.11 | 0.157 |

0 All data on singletons and first born of multiples <30 weeks gestation and admitted to an NHS Neonatal unit
1 Percentage point difference in uptake between mean across the 12m pre-NPP, and mean across the four years post-NPP
2 Crude regression of uptake post-implementation compared with pre-implementation.
3 Fully adjusted model: includes interaction between pre-post period and study month to capture change in slope as well as step-change. Adjusted for covariates as monthly aggregates: mean maternal age, proportion who identified as of white British ethnicity, mean IMD decile, proportion of multiple births, proportion with pregnancy hypertension, reported smokers, type of birth (c-section versus vaginal delivery), birthweight adjusted for gestational age as a z-score. Model weighted on unit size.
4 Additionally adjusted for level of birth unit, and regional clustering by AHSN.

#### 4.1.5 Secondary impact analysis (Bayesian structural time series model)

The default model showed a mean English uptake across the post-intervention period of 84%, and a predicted counterfactual mean uptake of 71% (95% Bayesian Credible Interval 66% to 76%). Subtracting the predicted from the observed gives an estimate of the causal impact of the NPP intervention of 13.4 percentage points absolute increase in MgSO4 uptake (95% BCI 8.8 to 17.9, p=0.001), or 19.0% (95% BCI 11.6%, 27.1%) relative change. As there was departure from the parallel trends assumption (English, Scottish and Welsh trends did not behave in a similar manner in the pre-intervention period), the estimated impact here needs to be interpreted with caution, and is likely to be overestimating the effect.

Post-hoc exploration of a model with a polynomial transformation (to account for the ceiling effect in the data) gave findings comparable to the main regression analyses, of 3.3 percentage points absolute increase (95% CI - 0.72, 7.2), or a 4% relative increase (95% CI -1%, +9%, p=0.058) compared to the counterfactual of England with no NPP. However, as a post-hoc analysis, this too must be interpreted with caution. (Figure 4)

**Figure 4:**
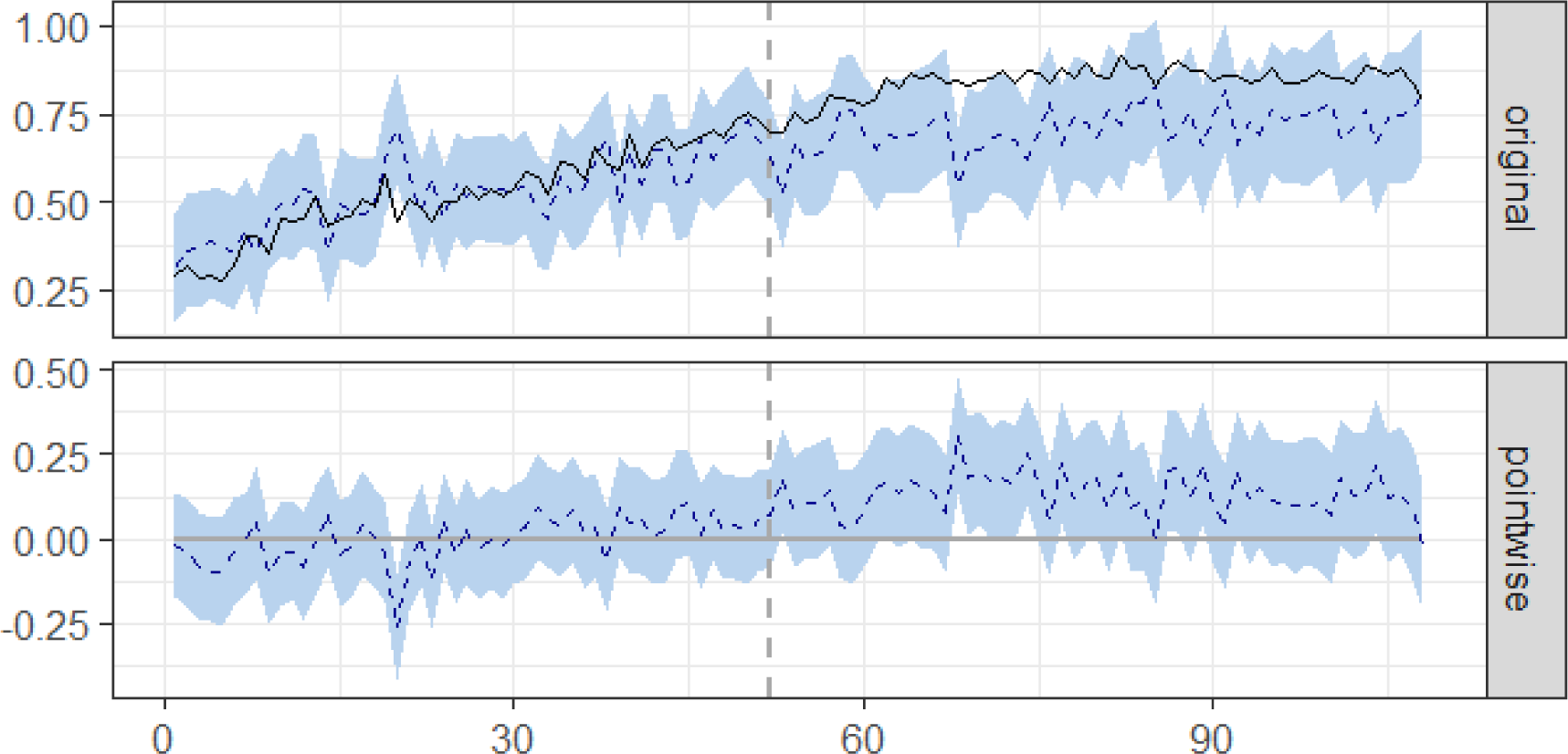
Bayesian causal impact plot of observed vs counterfactual predicted uptake

#### 4.1.6 Impact of the COVID-19 pandemic

The post-pandemic trend in England is suggestive of slight decline in uptake from the start of 2020 to the end of 2022 (most recent data). This period includes the peaks of the COVID-19 pandemic and UK lockdowns. Post-hoc exploration (in NNRD data) found that use of antenatal steroids (another protective treatment for preterm babies) had an almost identical decline over this same period. (Figure 5)

**Figure 5:**
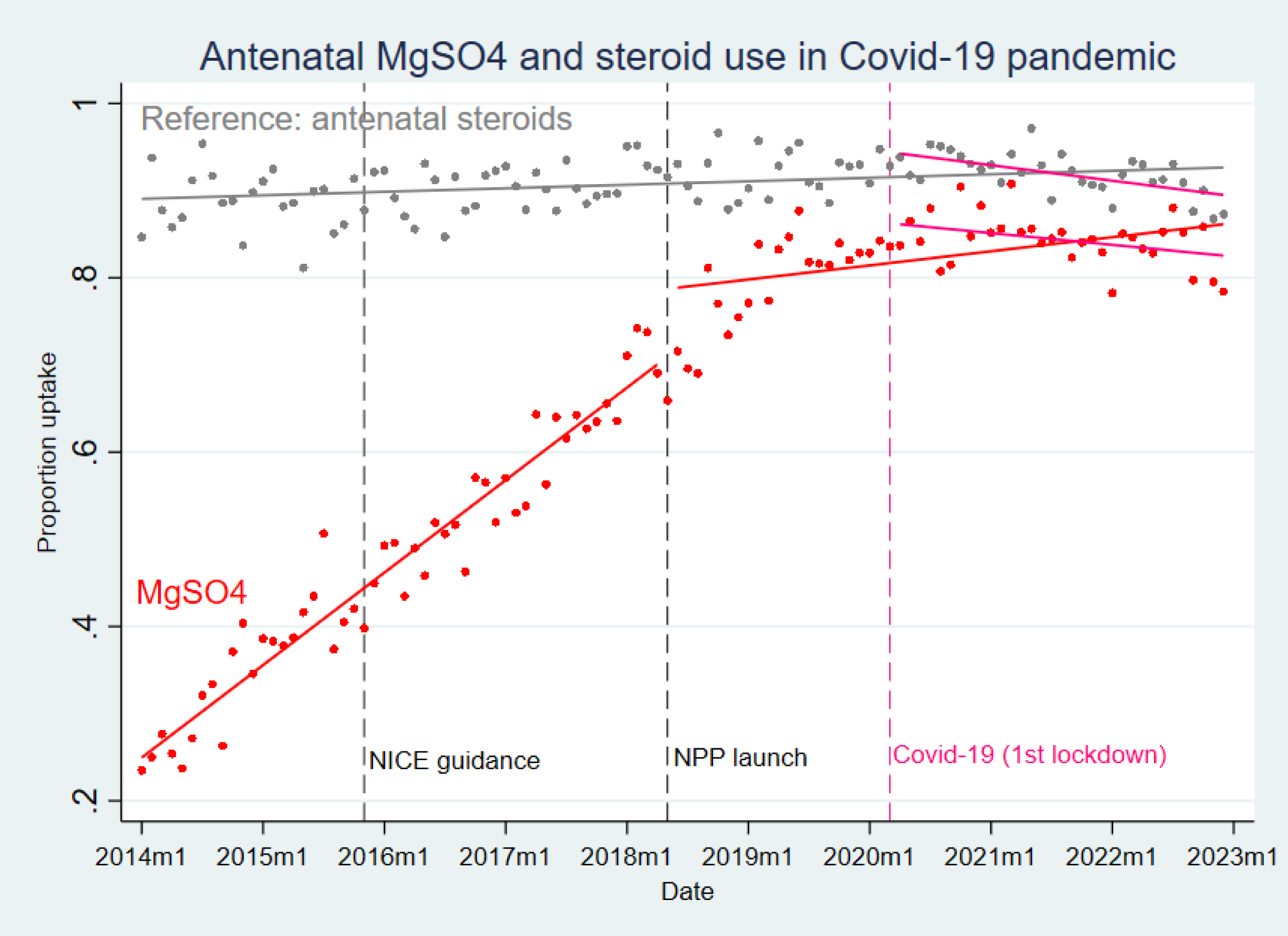
Impact of Coronavirus-19 pandemic on MgSO4 uptake in England

#### 4.1.7 Sociodemographic predictors of receiving MgSO4 in England

Individual-level analysis indicated historical disparities in receipt of MgSO4. Up to 2018, mothers who lived in the North as compared to the South of England (OR 0.62, 95% CI 0.57 to 0.67, p<0.001), or with a history of smoking (OR 0.72, 95% CI 0.64 to 0.80, p<0.001), had significantly lower odds of receiving MgSO4. In the four years since the NPP roll-out (2018 to 2022), the North/South divide in care had significantly attenuated (OR 0.93, 95% CI 0.83 to 1.03, p=0.167). Individual-year analysis indicated that this disparity had equalised by 2019. A history of smoking was however still predictive of lower odds of receiving MgSO4 (OR 0.66, 95% CI 0.58 to 0.76, p<0.001). Post-hoc exploration of reasons MgSO4 was not given did not indicate any differences in reasons between those with, versus without, a history of smoking. Smoking was itself socially patterned with younger mothers, white British mothers, and mothers from more deprived areas or from the North of England all more likely to have a history of smoking,

Across all time, older maternal age was predictive of fractionally higher odds of receiving MgSO4 (OR 1.01, 95% CI 1.00 to 1.01, p=0.009, indicating an average increase of around 1% in odds of receiving MgSO4 for each increase in year of age). There was no evidence that maternal ethnicity or area deprivation were predictive of odds of receiving MgSO4 after adjusting for other factors. (Table 4)

**Table 4:** Sociodemographic predictors of receiving MgSO4 in England.

|  | Pre-NPP<sup>0</sup> |  |  | Post-NPP<sup>1</sup> |  |  |
| --- | --- | --- | --- | --- | --- | --- |
|  | OR | 95% CIs | p-value | OR | 95% CIs | p-value |
| IMD decile<br>(increasing deprivation) | 1.01 | 0.99 to 1.03 | 0.180 | 1.01 | 0.99 to 1.04 | 0.162 |
| Maternal ethnicity<br>(white British) | 1.07 | 0.97 to 1.17 | 0.170 | 0.94 | 0.84 to 1.05 | 0.243 |
| Maternal age (years) | 1.00 | 0.99 to 1.01 | 0.287 | 0.99 | 0.99 to 1.01 | 0.928 |
| History of smoking | 0.72 | 0.64 to 0.80 | <0.001 | 0.66 | 0.58 to 0.76 | <0.001 |
| North of England vs South | 0.62 | 0.57 to 0.67 | <0.001 | 0.93 | 0.83 to 1.03 | 0.167 |

0. 2014 to May 2018 1. May 2018 to Dec 2022

### 4.2 Health Economic results

#### 4.2.1 NPP economic evaluation results

##### Main analysis

Our analysis of the impact of the NPP is illustrated in Figure 6, which shows the difference between the predicted level of MgSO4 use over time compared to a counterfactual level of MgSO4 use representing what may have occurred in the absence of the NPP. Our main analysis uses a linear distribution to estimate this counterfactual based on the pre-NPP predicted trend (Figure 6a) and a sensitivity analysis uses a beta distribution to estimate this counterfactual (Figure 6b).

**Figure 6.**
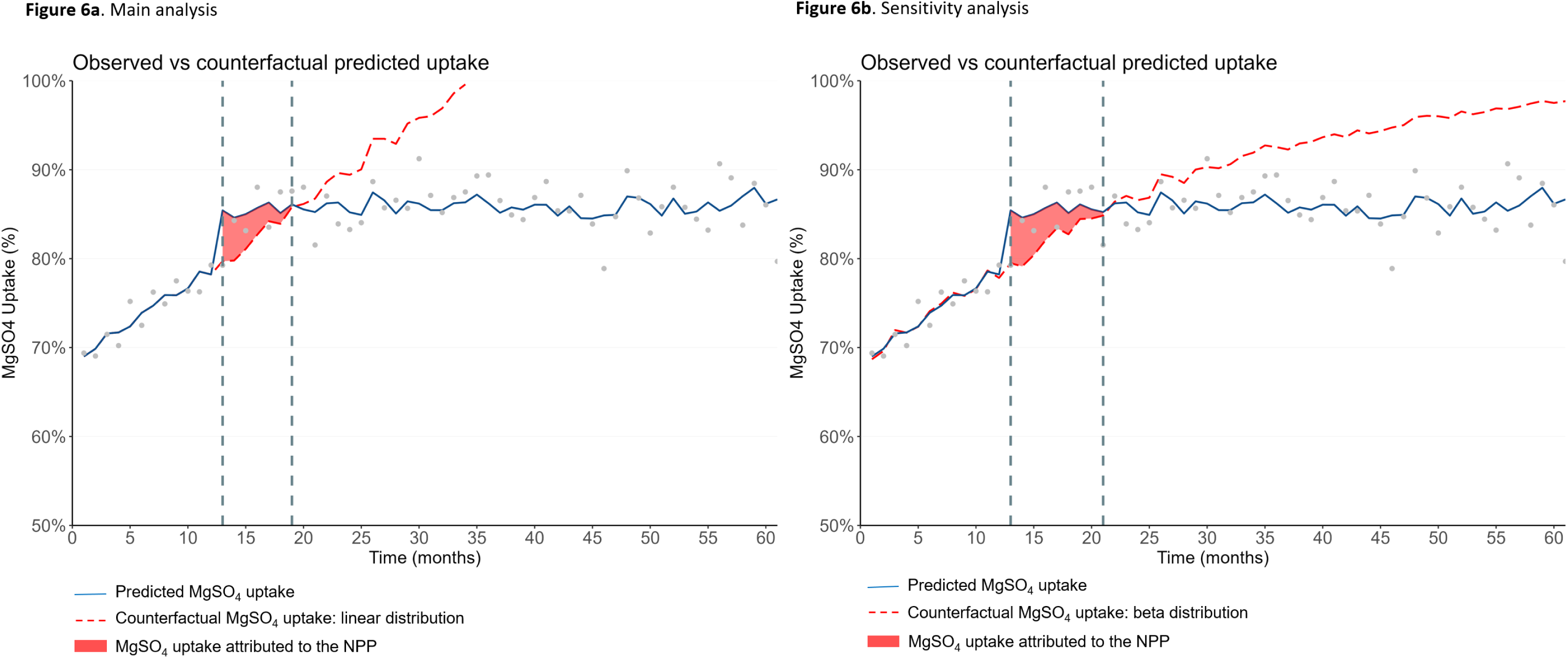
Predicted MgSO4 uptake, Counterfactual and Area-Between-Curves from Interrupted Time Series analyses.

Our probabilistic main analysis estimates that the additional use of MgSO4 attributed to the NPP was equivalent to 3.0% on average over seven months, which equates to an additional 64 of the 2,136 pre-term (<30 weeks gestation) babies receiving treatment (Table 5). Accounting for the total cost of the NPP (£936,747) and the lifetime health gains and cost savings of MgSO4 treatment, the societal NMB of NPP was about £597,000, or £279 per preterm baby, at a willingness-to-pay threshold of £20,000 per QALY. The probability of NPP being cost-effective was 89% at a willingness-to-pay threshold of £20,000 per QALY (Table 5 and Figure 7).

**Figure 7.**
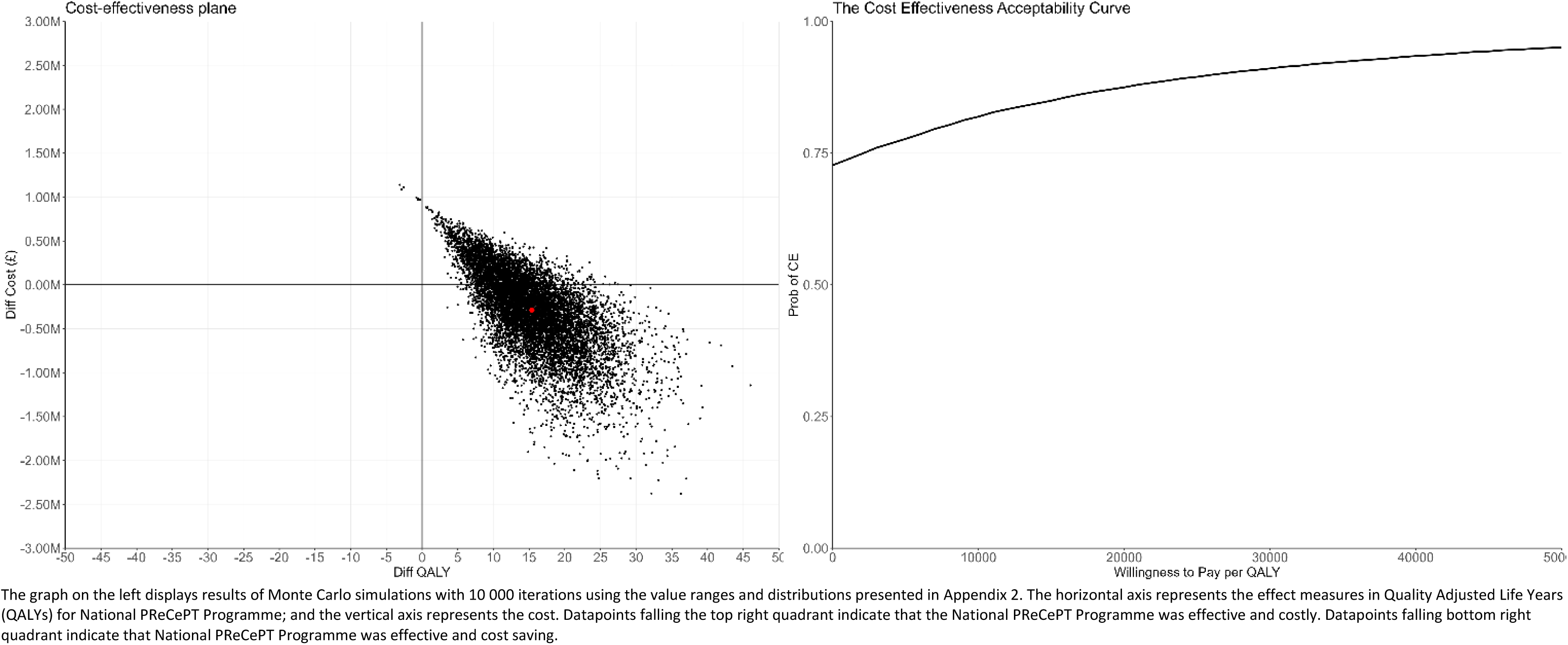
Cost-Effectiveness Plane and Cost-Effectiveness Acceptability Curve from Linear Interrupted Time Series analysis (<30 weeks gestation).

**Table 5.** Probabilistic Cost-Effectiveness Results of the NPP from Interrupted Time Series Analysis (<30 weeks gestation).

|  | Main analysis<br>(linear distribution<br>counterfactual) | Sensitivity analysis<br>(beta distribution<br>counterfactual) |
| --- | --- | --- |
| Period of benefit, months | 7 | 10 |
| Number of pre-term babies ($\leq 3$ weeks), $N$ | 2,136 | 3,129 |
| Change in proportion of pre-term babies treated with MgSO<sub>4</sub>, $\Delta b_i$, % | 3.0% (1.5%; 4.5%) | 2.9% (2.3%; 3.6%) |
| Net Increment of pre-term babies treated with MgSO<sub>4</sub>, $\Delta pat$ | 64 (32; 97) | 92 (72; 112) |
| Net cost of implementation, $\Delta C_i$, £ | 936,747 | 936,747 |
| Implementation cos-effectiveness, $\Delta C_i / \Delta Pat$, £ per additional patient treated | 14,576 (29,284; 9,669) | 10,219 (13,040; 8,386) |
| Lifetime health effect of MgSO<sub>4</sub> treatment per patient, $\Delta bt$, QALY | 0.24 (0.16; 0.33) | 0.24 (0.16; 0.33) |
| Lifetime costs of MgSO<sub>4</sub> treatment per patient, $\Delta ct$, £ | -19,064 (-13,310; -25,648) | -19,064 (-13,310; -25,648) |
| **Net Monetary Benefit of the Policy, $NMB_P$, £<sup>1</sup>** | **596,538 (-221,748; 1,541,786)** | **1,251,511 (558,115; 2,071,244)** |
| **Probability of being cost-effective, %** | **89%** | **100%** |

<sup>1</sup> At a willingness-to-pay threshold of £20,000 per QALY

The sensitivity analysis (with the beta distribution counterfactual) estimates a longer period of impact – an additional three months – over which there was additional use of MgSO4 attributed to the NPP equivalent to 2.9% on average, which equates to an additional 92 of 3,129 pre-term babies received treatment. Accounting for the total cost of the NPP and the lifetime health gains and cost savings of MgSO4 treatment, the societal Net Monetary Benefit of NPP was £1.3m, or £400 per preterm baby. The probability of NPP being cost-effective was 100% (Table 5).

##### Secondary analyses

Expanding the probabilistic analysis to include babies of less than 32 weeks gestation estimates that the additional use of MgSO4 attributed to the NPP was equivalent to 4.4% on average over nine months, which equates to an additional 215 pre-term babies treated. As the total cost of the NPP was fixed and not sensitive to the number of babies treated, the NMB of the NPP was £4.2m, or £853 per pre-term baby, at a willingness-to-pay threshold of £20,000 per QALY. The probability of NPP being cost-effective was 100% at this willingness-to-pay threshold (Table 6 and Figure 8).

**Figure 8.**
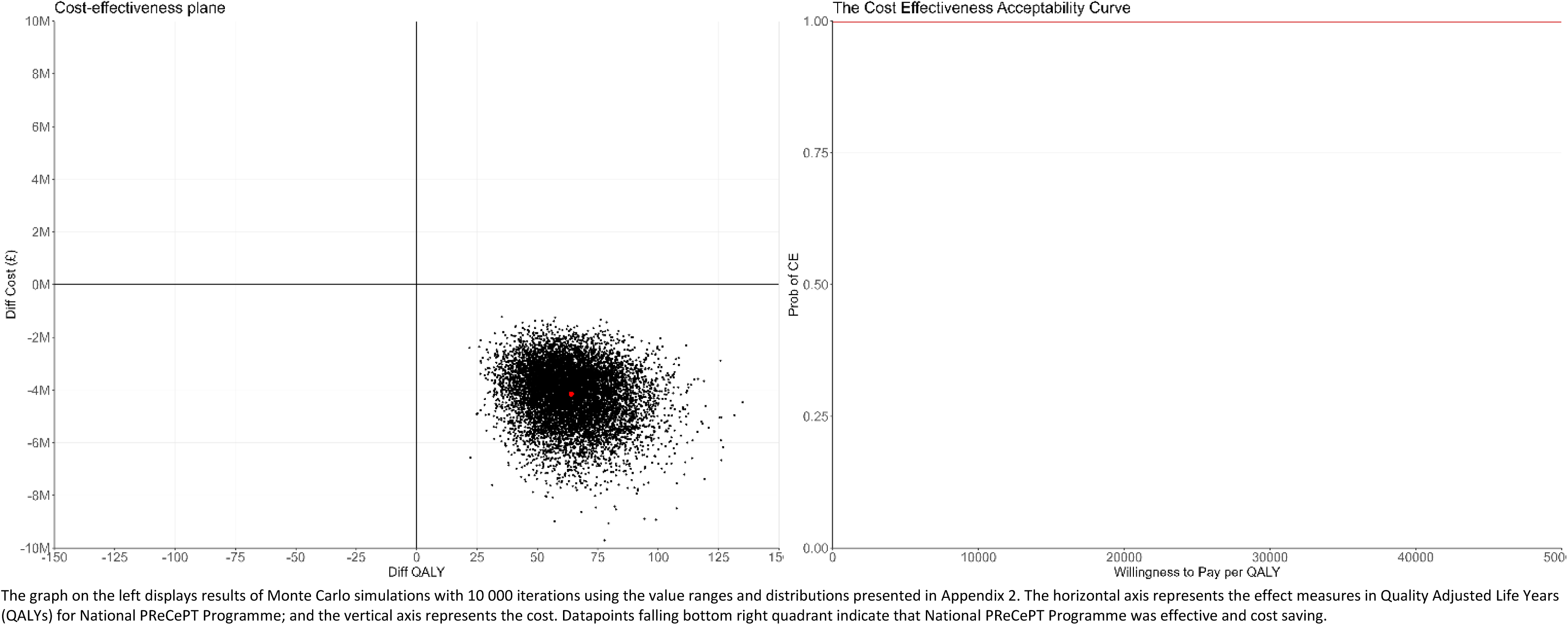
Cost-Effectiveness Plane and Cost-Effectiveness Acceptability Curve from Linear Interrupted Time Series analysis (<32 weeks gestation).

**Table 6.** Probabilistic Cost-Effectiveness Results of the NPP from Interrupted Time Series Analysis (<32 weeks gestation).

|  | Main analysis<br>(linear distribution<br>counterfactual) | Sensitivity analysis<br>(beta distribution<br>counterfactual) |
| --- | --- | --- |
| Period of benefit, months | 9 | 12 |
| Number of pre-term babies ($\leq 3$ weeks), $N$ | 4,923 | 7,768 |
| Change in proportion of pre-term babies treated with MgSO<sub>4</sub>, $\Delta b_i$, % | 4.4% (2.7%; 6.0%) | 3.4% (3.0%; 3.9%) |
| Net Increment of pre-term babies treated with MgSO<sub>4</sub>, $\Delta pat$ | 215 (133; 297) | 267 (230; 303) |
| Net cost of implementation, $\Delta C_i$, £ | 936,747 | 936,747 |
| Implementation cos-effectiveness, $\Delta C_i / \Delta Pat$, £ per additional patient treated | 4,350 (7,031; 3,153) | 3,508 (4,065; 3,087) |
| Lifetime health effect of MgSO<sub>4</sub> treatment per patient, $\Delta bt$, QALY | 0.24 (0.16; 0.33) | 0.24 (0.16; 0.33) |
| Lifetime costs of MgSO<sub>4</sub> treatment per patient, $\Delta ct$, £ | -19,064 (-13,310; -25,648) | -19,064 (-13,310; -25,648) |
| **Net Monetary Benefit of the Policy, $NMB_P$, £<sup>1</sup>** | **4,199,799 (1,986,016; 6,803,391)** | **5,433,488 (3,668,040; 7,508,699)** |
| **Probability of being cost-effective, %** | **100%** | **100%** |

<sup>1</sup> At a willingness-to-pay threshold of £20,000 per QALY

The same analysis above but with the beta distribution counterfactual estimates 12 months as the period of impact (three additional months than the analysis with the linear counterfactual). The increment of MgSO4 uptake attributed to the NPP was 3.4% on average, which equates to an additional 267 of 7,768 pre-term babies receiving treatment. Accounting for the total cost of the NPP and the lifetime health gains and cost savings of MgSO4 treatment, the societal NMB of NPP was £5.4m, or £700 per preterm baby. The probability of NPP being cost-effective was 100% (Table 6).

We also conducted an analysis for the babies with 32 weeks gestation up to 34 weeks gestation included. Our analysis shows that the additional use of MgSO4 attributed to the NPP was equivalent to 7.3% on average over seven months, this means an additional 961 babies received treatment (Table 7). The sensitivity analysis with a beta distribution for the counterfactual shows a 6.5% increment attributed to the NPP over 12 months, this equates to 698 additional patients receiving the treatment. Currently, there is no available estimate of the cost-effectiveness of MgSO4 treatment for babies with more than 31 weeks gestation. For this reason, we did not include these babies in the NMB analysis of the NPP.

**Table 7.** Results of the NPP from Interrupted Time Series Analysis (32-34 weeks gestation).

|  | **Main analysis**<br>(linear distribution<br>counterfactual) | **Sensitivity analysis**<br>(beta distribution<br>counterfactual) |
| --- | --- | --- |
| Period of benefit, months | 15 | 12 |
| Number of pre-term babies ($\leq 3$ weeks), $N$ | 13,243 | 10,665 |
| Change in proportion of pre-term babies treated with $\text{MgSO}_4$, $\Delta b_i$, % | 7.3% (2.9%; 11.6%) | 6.5% (5.6%; 7.5%) |
| Net Increment of pre-term babies treated with $\text{MgSO}_4$, $\Delta pat$ | 961 (387; 1,535) | 698 (593; 803) |

#### 4.2.2 Net monetary benefit of optimal MgSO4 implementation

##### Seeking the level of optimal MgSO4 implementation

MgSO4 uptake has increased over the nine years to December 2022 in England, Scotland and Wales (Figure 9) both for babies of less than 30 weeks gestation (Figure 9a) and those of 30 or 31 weeks gestation (Figure 9b. However, at a national level, a plateau in uptake and even a decrease towards the end of the observed period is apparent. The uptake for babies of 30-31 weeks gestation is lower than that for babies with less than 30 weeks gestation, particularly in Scotland.

**Figure 9.**
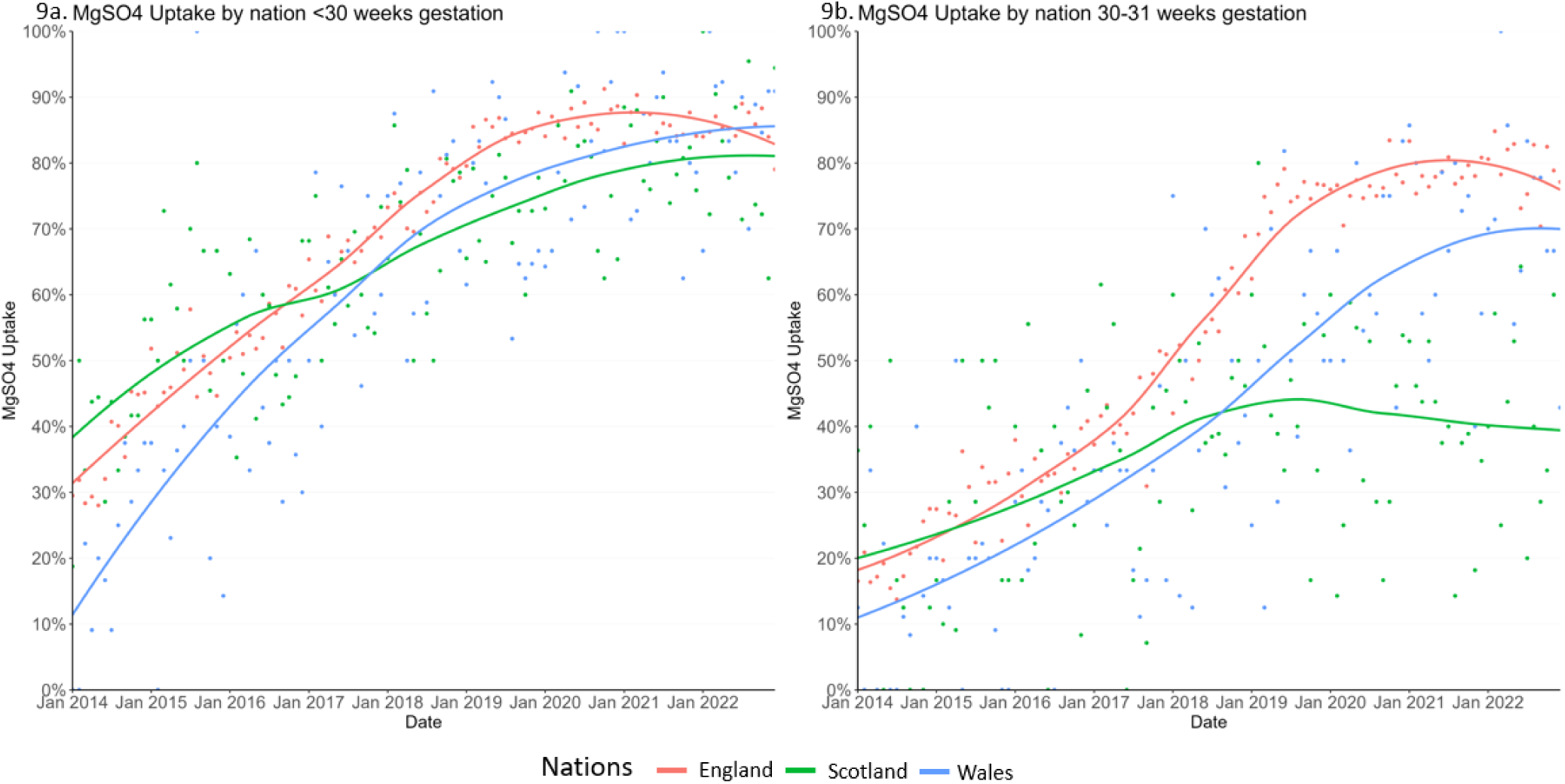
Uptake of MgSO4 in England, Scotland and Wales from Jan 2014 to Dec 2022.

Figure 10 shows the bimonthly uptake from January 2014 to December 2022 for the different ODNs in England for babies of less than 30 weeks gestation. The experience at ODN-level differs: all of them increased MgSO4 uptake over time, and some of them, such as the ODN 200708, have been able to continue to increase uptake after the NPP. However, most of the ODNs were not able to achieve ongoing improvement in MgSO4 uptake over time, beyond around 90%.

**Figure 10.**
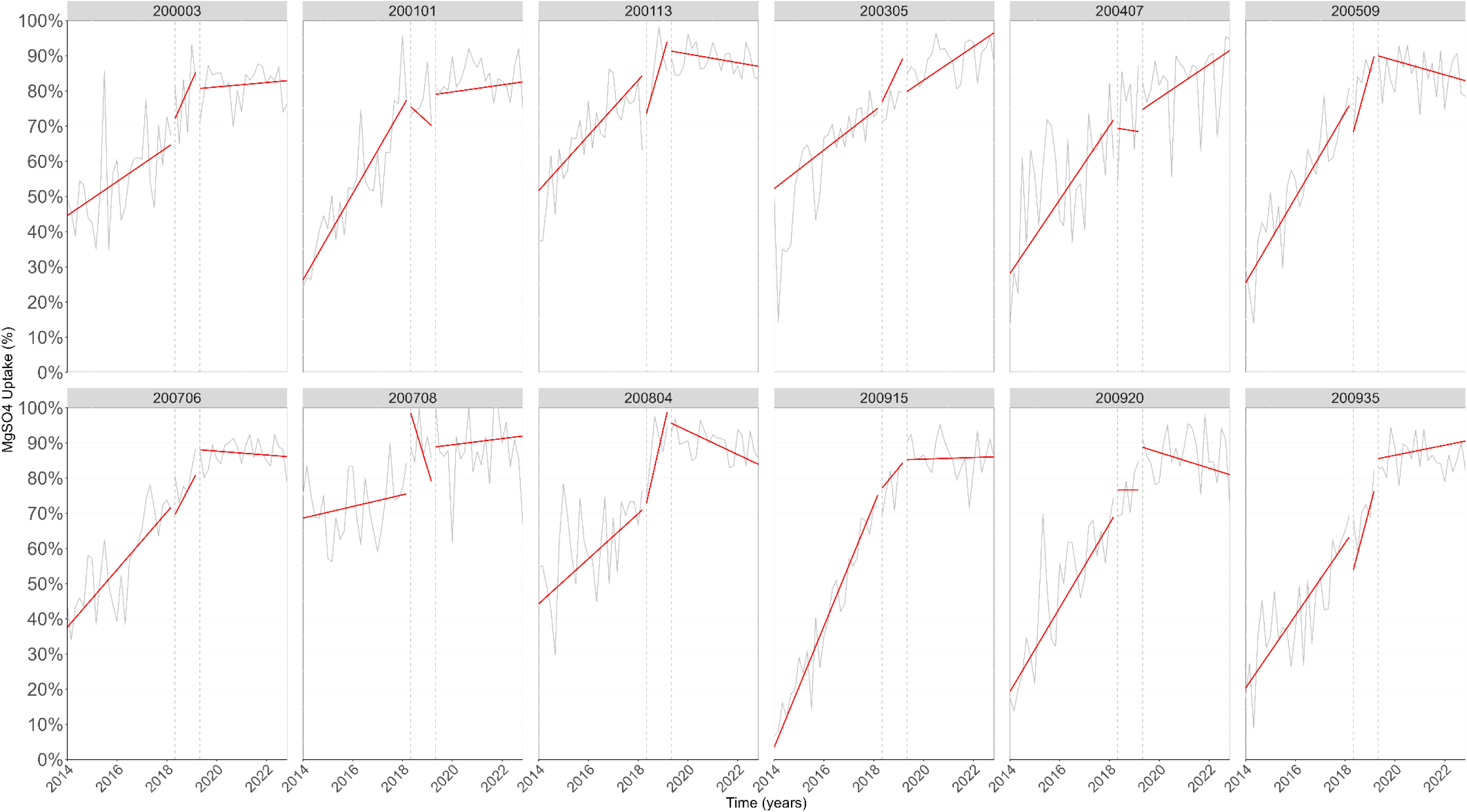
MgSO4 uptake by ODN from Jan 2014 to Dec 2022 (<30 weeks gestation).

We view the optimal implementation of MgSO4 as 95% uptake, as higher uptake may be clinically unrealistic. This upper threshold for MgSO4 uptake will allow us in the next section to estimate the NMB not generated due to MgSO4 not being optimally implemented and the potential value for future implementation initiatives.

##### Net Monetary Benefit of MgSO4 implementation results

For babies of less than 30 weeks’ gestation in each nation, Figure 11 shows the monthly NMB of implementing MgSO4 (green line), the NMB if MgSO4 was optimally implemented (i.e., 95% uptake – blue line), and the red area between these two lines represents the NMB not generated due to MgSO4 not optimally implemented (represented again in the bar charts on the right of the figure). The area in red and consequently the values in the bar charts are reduced progressively (Figure 11). This indicates that the increased MgSO4 uptake over time, closer to the optimal uptake, has generated NMB associated with the lifetime health gains and cost-savings of MgSO4 for the prevention of CP and, therefore, reducing progressively the opportunity cost associated with not administrating this treatment.

**Figure 11.**
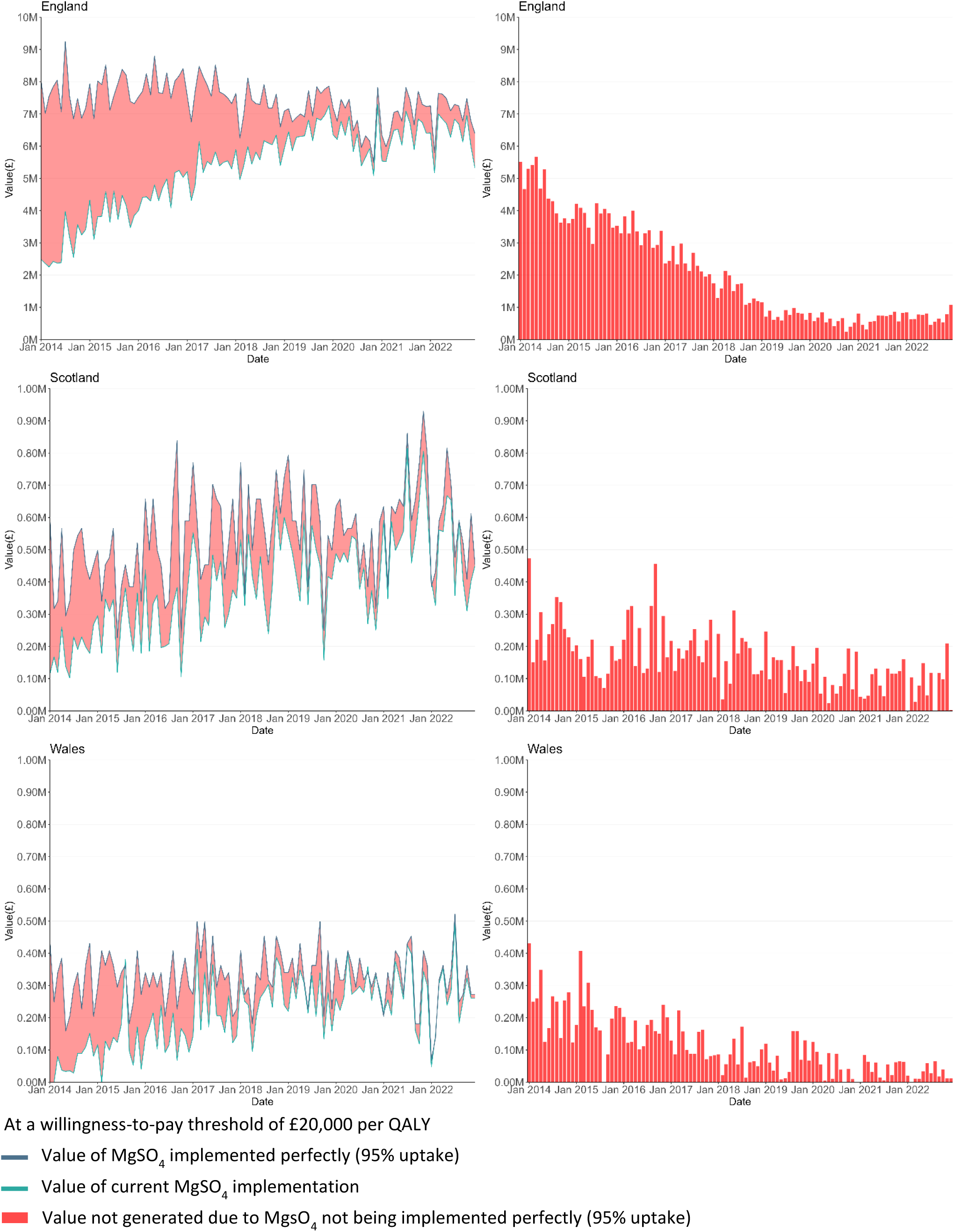
Value of Optimal Implementation of MgSO4 uptake in England, Scotland and Wales from Jan 2014 to Dec 2022 (<30 weeks gestation).

For instance, in England in 2014, the NMB generated for providing this treatment to 36% of babies (N = 4,003) was more than £45m; yet, there was approximately £75m of NMB forgone for the remaining 59% uptake that did not happen (Table 8). This will be equivalent to the area in red and the sum of all bars in Figure 11 for the year 2014. In the same year in Scotland, the 39% uptake (N = 237) generated NMB of approximately £2m, but still around £3m of NMB was not generated. Similarly in Wales, the 20% uptake (N = 160) generated NMB of £0.7m, but still around £2.9m was forgone. In 2022, the 85% uptake (N=3,744) generated NMB of £76m in England, and £8.5m of NMB was forgone due to sub-optimal implementation (Table 8). The NMB forgone equate to £1.0m for Scotland and £0.3m for Wales (Table 8).

**Table 8.** Net Monetary Benefit of MgSO4 implementation in 2014 and 2022 for England, Scotland and Wales (2019 prices).

| Gestation age (weeks) |  | England |  | Scotland |  | Wales |  |
| --- | --- | --- | --- | --- | --- | --- | --- |
|  |  | 2014 | 2022 | 2014 | 2022 | 2014 | 2022 |
| <30 | Number of babies, N | 4,003 | 3,744 | 237 | 292 | 160 | 152 |
|  | Uptake of MgSO<sub>4</sub>, % | 36% | 85% | 39% | 82% | 20% | 86% |
|  | NMB of optimal MgSO<sub>4</sub> implementation (95%), £ | 120,535,455 | 84,843,907 | 5,370,728 | 6,617,100 | 3,625,808 | 3,444,518 |
|  | NMB of current implementation, £ | 45,594,359 | 76,325,340 | 2,199,332 | 5,689,939 | 733,281 | 3,144,234 |
|  | NMB forgone due to suboptimal implementation, £ | 74,941,096 | 8,518,567 | 3,171,396 | 950,255 | 2,892,527 | 300,284 |
| 30 and 31 | Number of babies, N | 3,042 | 2,553 | 139 | 215 | 134 | 117 |
|  | Uptake MgSO<sub>4</sub>, % | 19% | 79% | 19% | 42% | 13% | 72% |
|  | NMB of optimal MgSO<sub>4</sub> implementation (95%), £ | 68,935,675 | 57,854,299 | 3,149,921 | 4,872,180 | 3,036,614 | 2,651,372 |
|  | NMB of current implementation, £ | 14,035,014 | 48,131,724 | 671,309 | 2,142,839 | 430,035 | 2,000,318 |
|  | NMB forgone due to suboptimal implementation, £ | 54,900,660 | 9,722,575 | 2,478,611 | 2,729,340 | 2,606,580 | 651,054 |
| <32 (Total) | **Number of babies, N** | **7,045** | **6,297** | **376** | **507** | **294** | **269** |
|  | **Uptake MgSO<sub>4</sub>, %** | **28%** | **82%** | **29%** | **62%** | **17%** | **79%** |
|  | **NMB of optimal MgSO<sub>4</sub> implementation (95%), £** | **189,471,129** | **142,698,206** | **8,520,649** | **11,489,279** | **6,662,422** | **6,095,890** |
|  | **NMB of current implementation, £** | **59,629,374** | **124,457,064** | **2,870,641** | **7,832,779** | **1,163,315** | **5,144,552** |
|  | **NMB forgone due to suboptimal implementation, £** | **129,841,756** | **18,241,142** | **5,650,007** | **3,679,595** | **5,499,107** | **951,338** |

Net monetary benefit (NMB) was estimated at a willingness-to-pay threshold of £20,000 per QALY

Figure 12 and Table 8 show the results for this analysis for babies of 30 or 31 weeks gestation, and all babies less than 32 weeks gestation. For all babies of less than 32 weeks gestation in 2022 in England, the NMB generated due to MgSO4 uptake was approximately £125m. This number equates to approximately £8m in Scotland and £5m in Wales (Table 8).

**Figure 12.**
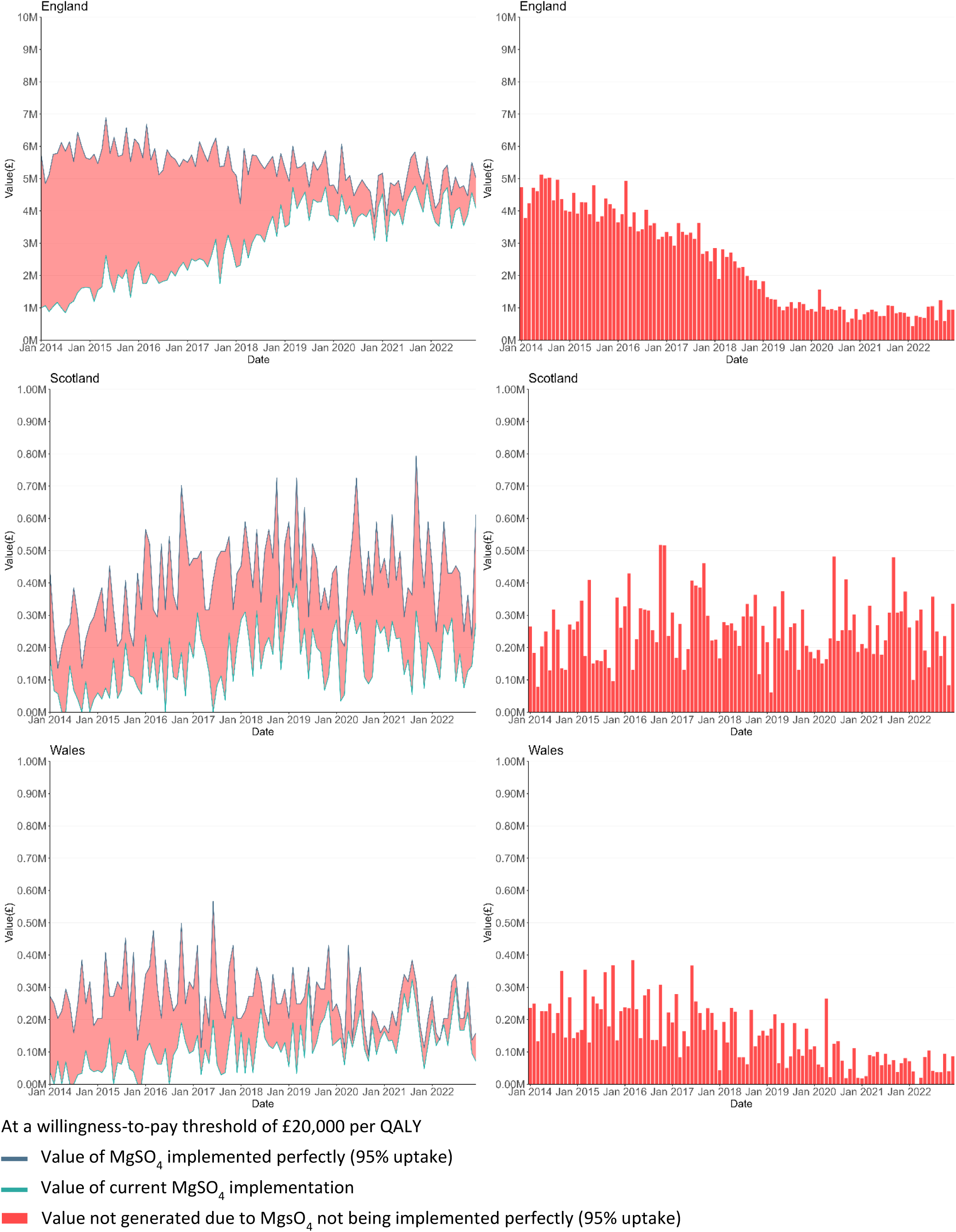
Value of Optimal Implementation of MgSO4 uptake in England, Scotland and Wales from Jan 2014 to Dec 2022 (30 and 31 weeks gestation).

##### The potential cost-effectiveness of future implementation programmes

The NMB not generated due to suboptimal MgSO4 uptake implementation also indicates how much resource could have been invested to achieve the 95% optimal treatment uptake, in other words, the value of optimal implementation. In 2022, this equates to approximately £18.2m in England, £3.7m in Scotland and £1.0m in Wales (for babies of less than 32 weeks gestation) (Table 8). This gives us an upper threshold of how much could be spent on implementation initiatives, and be viewed as representing good value for money when taking a societal lifetime perspective.

We have also estimated the cost-effectiveness of three hypothetical future national implementation programmes for the three nations illustrating different implementation effectiveness scenarios: low performance (1% increase in MgSO4 uptake over 12 months), mid-performance (5%), and high performance (10%). For the purpose of this analysis, the implementation costs were assumed to be similar to those of the NPP in England at £6,044 per unit in each nation (Table 9), although in practice any future implementation programme in England may be focused on units or networks experiencing comparatively weak performance.

**Table 9.** National Net Monetary Benefit of Implementation for potential Initiatives to increase MgSO4 Uptake in England, Scotland and Wales with three different Implementation Effectiveness and Implementation Cost for a single year period (<32 weeks gestation).

|  | England | Scotland | Wales |
| --- | --- | --- | --- |
| Number of babies, N | 6,297 | 507 | 269 |
| Net cost of implementation, $\Delta C_i$, £ | 936,747 | 84,609 | 54,392 |
| Lifetime health effect of MgSO<sub>4</sub> treatment per patient, $\Delta bt$, QALY | 0.24 (0.16; 0.33) | 0.24 (0.16; 0.33) | 0.24 (0.16; 0.33) |
| Lifetime costs of MgSO<sub>4</sub> treatment per patient, $\Delta ct$, £ | -19,054 (-13,310; -25,648) | -19,054 (-13,310; -25,648) | -19,054 (-13,310; -25,648) |
| **Low performance: 1% Implementation effect** |  |  |  |
| Increment of pre-term babies treated with MgSO<sub>4</sub>, $\Delta pat$ | 63 | 5 | 3 |
| Implementation cos-effectiveness, $\Delta Ci / \Delta Pat$, £ per additional patient treated | 14,876 | 16,688 | 20,220 |
| **Net Monetary Benefit of the Policy, $NMB_p$, £** | **565,557 (105,767; 1,087,892)** | **36,348 (-672; 78,403)** | **9,785 (-9,857; 32,098)** |
| **Mid performance: 5% Implementation effect** |  |  |  |
| Increment of pre-term babies treated with MgSO<sub>4</sub>, $\Delta pat$ | 315 | 25 | 13 |
| Implementation cos-effectiveness, $\Delta Ci / \Delta Pat$, £ per additional patient treated | 2,975 | 3,338 | 4,044 |
| **Net Monetary Benefit of the Policy, $NMB_p$, £** | **6,574,774 (4,275,823; 9,186,446)** | **520,177 (335,078; 730,455)** | **266,491 (168,283; 378,058)** |
| **High performance: 10% Implementation effect** |  |  |  |
| Increment of pre-term babies treated with MgSO<sub>4</sub>, $\Delta pat$ | 630 | 51 | 27 |
| Implementation cos-effectiveness, $\Delta Ci / \Delta Pat$, £ per additional patient treated | 1,488 | 1,669 | 2,022 |
| **Net Monetary Benefit of the Policy, $NMB_p$, £** | **14,086,296 (9,488,393; 19,309,638)** | **1,124,964 (754,766; 1,545,519)** | **587,374 (390,957; 810,509)** |

Implementation costs assumed to be similar to unit cost of the National PRECePT Programme
Values were calculated at a willingness-to-pay threshold of £20,000 per QALY
Number of babies (N) are estimated through the last observed period – 2022

The low performance scenario would be cost-effective in England with an estimated NMB of about £566,000, compared to £36,000 in Scotland, and £10,000 in Wales. While there is uncertainty about the impact in terms of NMB in Scotland and Wales at this level of uptake performance, the mid and high performance scenarios are associated with considerable NMB outcomes (Table 9). National implementation programmes achieving a 10% increment of MgSO4 uptake would be likely to be highly cost-effective, with a NMB of £14.1m in England, £1.1m in Scotland and £0.6m in Wales (Table 9).

We have further estimated the cost-effectiveness of hypothetical implementation programme scenarios with different levels of implementation cost and implementation effectiveness, and illustrate this in the ‘heatmaps’ shown in Figure 13. For England the scenarios are based on a range of implementation costs up to £2m and implementation effectiveness up to a 20% increment in MgSO4 uptake. Figure 13 shows that in most of the illustrated scenarios investing in implementing MgSO4 uptake would be cost-effective for the three nations. Programmes achieving even small increments of MgSO4 uptake are likely to be cost-effective, given that MgSO4 treatment is highly cost-effective when viewed from a societal lifetime perspective.

**Figure 13.**
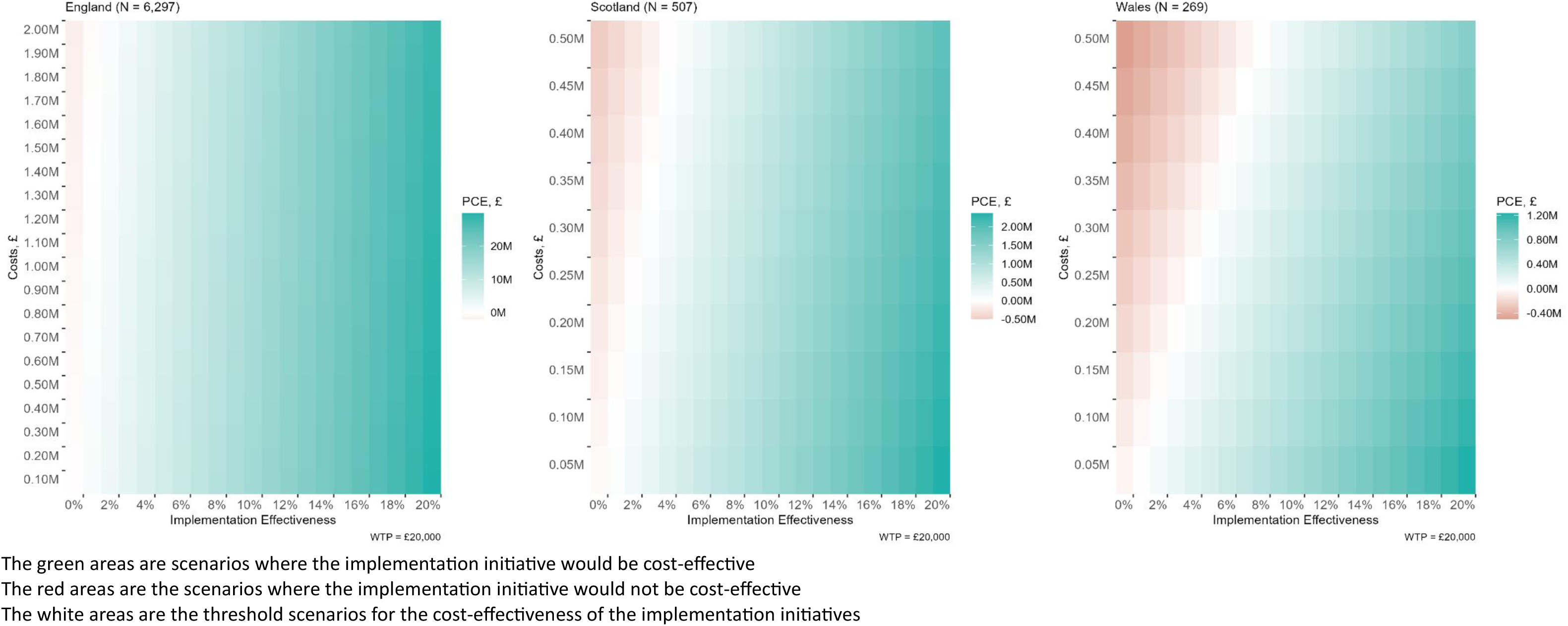
National Value of Implementation for potential Initiatives to increase MgSO4 Uptake in England, Scotland and Wales with different Implementation Effectiveness and Implementation Cost for a single year period (<32 weeks gestation).

### 4.3 Qualitative evaluation

#### 4.3.1 Findings

Thirteen participants were recruited to the study: eight participants from Wales, and five from Scotland. Seven participants were members of the neonatal team (consultant neonatologists and one Advanced Neonatal Nurse Practitioner), and five were members of the maternity team (consultant obstetricians and one midwife). One participant was a QI coach involved in the implementation of the MCQIC Preterm Perinatal Wellbeing Package (PPWP). See Table 10 for participant demographic information

**Table 10.**
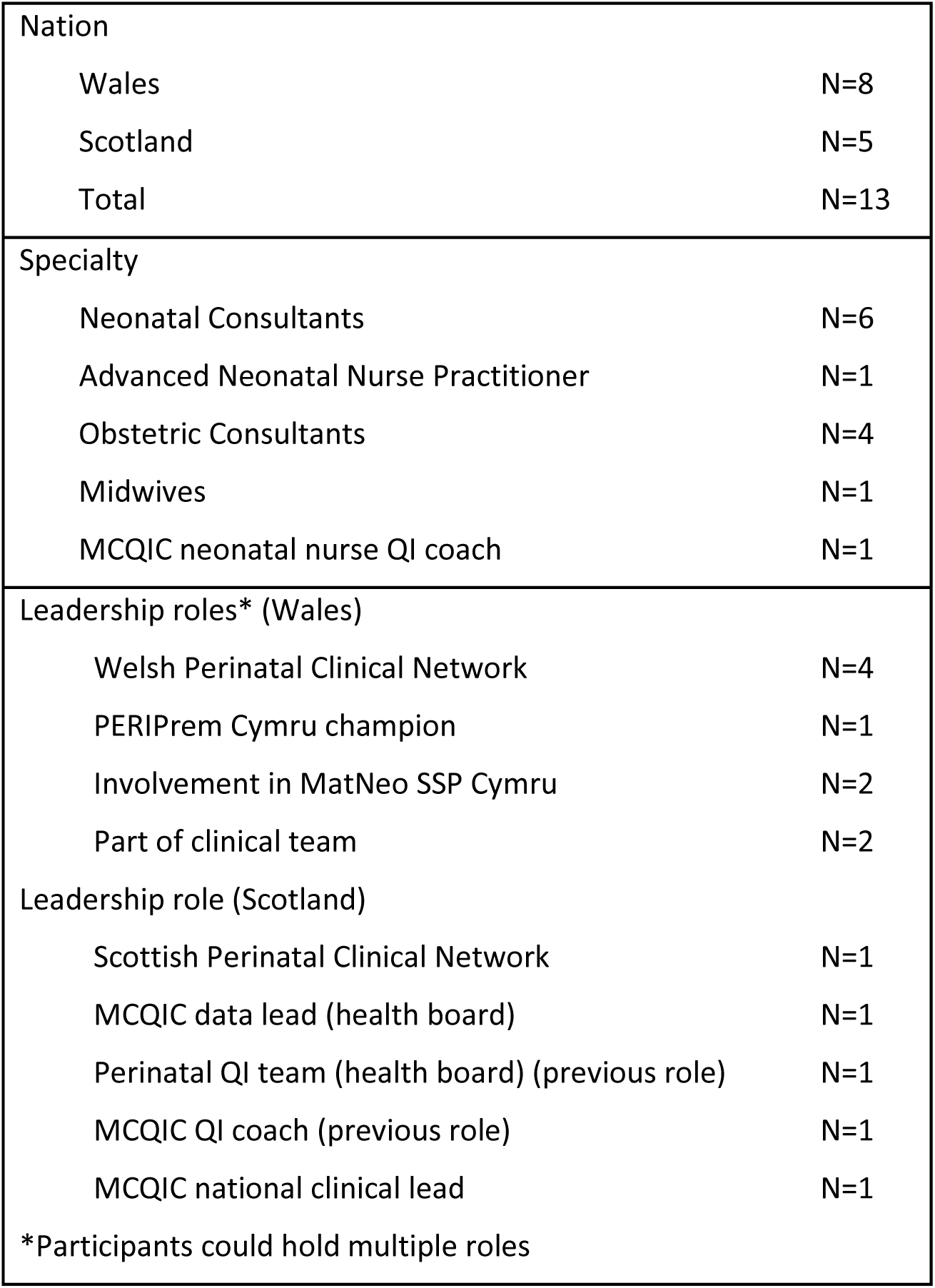
Qualitative interview participants.

#### 4.3.2. Mechanisms of Implementation

We present our findings using the NPT implementation mechanisms coherence building, cognitive participation, collective action and reflexive monitoring, and relating these to the context-specific drivers, barriers and enablers to implementing network- and locally-led improvement strategies and interventions.

##### Coherence-building

Coherence refers to how people make sense of a change to practice. This includes how they think it differs from what they currently do, how they assess the impact of the change on their role and responsibilities, and how they value it. It is important for all actors involved in preterm labour pathways to have a shared understanding of the intervention to ensure the intervention is delivered as intended across time and place. Even though MgSO4 is delivered by maternity teams, including midwives and obstetricians, it is for the therapeutic benefit of the unborn baby for whom neonatal teams are responsible. Therefore, MgSO4 administration is an indicator captured in the National Neonatal Audit Programme with the onus of data capture and audit on neonatal teams rather than obstetric teams. However, if the change in practice is conceptualised as a perinatal team intervention, professional and speciality boundaries are less pronounced.

The building of coherence around a new practice such as the routine administration of MgSO4 to eligible women is achieved through clinical evidence, clinical policies, guidelines, pathways, and processes, as well as local culture i.e., informal practices, intended to create meaning around what MgSO4 is, why, when, how, and to whom it should be administered.

Strong research evidence on the neuroprotective impact of MgSO4 when administered during preterm labour led to NICE guidance calling for all women between 24+0 and 29+6 weeks of pregnancy in established preterm labour or having a planned preterm birth within 24 hours, to be offered MgSO4, with the option to include pregnancies up to 34 weeks’ gestation (NICE 2016). This guidance was reiterated in Royal College of Paediatrics and Child health, and the Royal College of Obstetrics and Gynaecology guidance. Our analysis showed how divisions and boundaries across disciplinary and leadership structures, i.e. royal colleges, clinical leadership groups, and the clinical teams on the ground, and differing clinical priorities, created misalignments in the guidance provided by leadership, which in turn generated conflicts for staff on the ground, compounded by some perceived uncertainties in the evidence base. Because of their audit responsibilities however, neonatal teams were more motivated to change practice, and sense-making activities were concentrated within and led by neonatal teams. Incongruencies in how evidence and guidance were interpreted by the two specialties, and the perceived interference in obstetricians’ clinical practice could strain already difficult perinatal team relationships as the following excerpt illustrates.

> I may not get involved to read RCPCH guidelines. I am sure the neonatologist will not be so familiar with the RCOG guidelines. So it is up to our national bodies, such as RCOG, RCPCH, NICE, that they unify their proposed guidelines. Then how can you blame the obstetrician who says, “Well, I don’t know what your RCPCH guidelines says. My NICE guideline is saying (administer up to) 30 weeks.” (P07W, Obstetrics)

Study participants described national strategic intentions in both nations to unify guidance and create a coherent narrative around perinatal optimisation in general, and MgSO4 specifically through national preterm labour guidelines and care pathways. In Scotland, the MCQIC team had created PPWP which was launched in 2017. More recently both nations used the British Association of Perinatal Medicine (BAPM) Perinatal Optimisation and PERIPrem toolkits; PERIPrem used PReCePT methodologies to implement a perinatal optimisation bundle of interventions including MgSO4. Both of these toolkits adopted a perinatal approach to implementation, and participants from both nations described efforts to include actors from midwifery, obstetric and neonatal teams in implementation activities. In Scotland, PPWP sense-making activities had targeted neonatal teams, but this was perceived as one of the challenges to successful implementation. In relaunching the package, leads wanted to include the whole perinatal team in communication and awareness raising activities, particularly in helping obstetricians understand the evidence and address safety concerns around MgSO4 as the following excerpt illustrates

> A lot of it was understanding the why we want you to change your behaviours, and if you don’t know the evidence why would you change your behaviour? So, I think it’s having those shared common goals across all of our specialties, and building the team from that joined-up approach from the start, and not just working in a silo, that’s the biggest bit of advice. (P13S, Neonatology)

### Comparisons between the devolved nations and English experiences

Data from Wales and Scotland demonstrated how even though national and professional bodies had embedded MgSO4 in guidance, maternity teams were often unsure of how to operationalise the intervention given the nuances and uncertainties around safe administration e.g. eligibility criteria for women over 30 weeks. Strategic improvement intentions and initiatives were initiated by neonatal teams because the change in practice only benefitted their patients, but such efforts were thwarted by poor perinatal team relationships and misalignments in how the two teams understood the intervention. A culture of silo working by obstetric and neonatal teams, evident across the leadership structures, meant that teams attempted to create coherence from different information sources.

These findings echo PReCePT pilot^8^, NPP^9^ and Study evaluations. These evaluations demonstrated how the methodology used by PReCePT (and in particular the ‘enhanced support’ arm in the PReCePT trial^17^) was very effective in creating a collective sense of coherence among the perinatal team; making sure “everyone was singing from the same hymn book” was described as a powerful enabler of implementation. PReCePT strategies which contributed to improved perinatal teamworking and perinatal collective coherence included:

1. Having representatives from midwifery, obstetrics and neonatology in the leadership structure of PReCePT, including on the ground in the form of local champions ensured that voices from all actors were represented when trying to understand the intervention and the factors shaping its implementation. This in turn ensured the quality improvement intervention was a co-creation between all stakeholders.
2. Having a perinatal team of champions, created and opened up communication channels between clinical leadership and staff on the shop floor, and made the embeddedness of sense-making activities in routine meetings and conversations easier. This helped to communicate messages from clinical leads to intervention adopters quickly, and expedited clarifying of emerging clinical ambiguities and questions to ensure MgSO4 was appropriately administered.
3. Training all members of the perinatal team to create awareness of the intervention among all clinical groups involved in the care pathway of pregnant women. The PReCePT process evaluation included data capture of numbers of staff trained, acknowledging the importance of coherence for implementation. Embedding PReCePT in multi-professional training ensured all actors had the same understanding of the intervention and of each other’s roles, and cultivated perinatal team shared governance reinforcing the message that it was everyone’s responsibility. The shared understanding facilitated the redesign of care pathways and processes, and the redistribution of roles and responsibilities required.

Table 11 summarises the challenges to coherence identified, the strategies adopted by PReCePT to address these challenges, and the lessons learned through our research on what a QI intervention implementation framework should include to ensure collective coherence of intervention implementers and adopters.

**Table 11:** Coherence: Overview of findings and lessons learned.

| Coherence | Challenges to coherence-building | PReCePT strategies | Lessons for a QI intervention Implementation Framework |
| --- | --- | --- | --- |
|  | Operationalising national guidance & evidence ambiguous ≥30wks/complex cases<br>Misalignment between neonatal and obstetric understandings of what needs to be done<br>Coherence not spread across all members of the team providing care (e.g. midwives, juniors etc)<br>Poor perinatal teamworking culture a blocker to co-creating coherence | *“Everyone singing from the same hymnbook”*<br>PReCePT co-produced toolkit to define intervention & operationalisation (pathways, guidance, evidence etc)<br>National, regional, and local communication strategy and activities e.g. launch events, social media campaigns targeting all stakeholders (including patients/service users)<br>Perinatal clinical leadership providing MgSO4 steer and clarity among adopters (maternity & neonatal teams)<br>Coaching implementers to ensure clarity around the intervention, its components, and implementation roles | Coherence around the QI and clinical intervention needs to be co-produced by all stakeholders i.e. multi- disciplinary/- professional and service user consensus<br>Evidence and clinical guidance specific to the intervention need to be brought together in a single cross-disciplinary guideline<br>Intervention needs to be communicated in a clear and concise message<br>Clinical (horizontal) leadership is needed for steering operationalisation of guidance across time and space (implementation settings)<br>Toolkits, coaching & training of implementers and adopters help create and maintain coherence across time and space i.e through multi-professional training |

#### Cognitive Participation

Cognitive participation is the relational work that people do to build and sustain a community of practice around a new technology or complex intervention. It relates to people’s understanding of their role in implementing the intervention and that of their teams. In this study it refers to networks with a vested interest in MgSO4 guidance adherence such as the perinatal strategic clinical networks, perinatal optimisation and quality improvement networks e.g. BAPM, MQIC, as well as local clinical teams, and the activities organised by these networks to bring people together to understand and plan implementation. Activities can take place within and across micro (the midwifery, obstetric, and neonatal teams), meso (all the teams that make up the perinatal service), and macro (Trust/Health Board and national bodies) levels of the healthcare system, formally e.g. as part of huddle and handover meetings, network-level meetings, or webinars and learning events, or informally between members of the team at the point of care delivery.

Creating perinatal and multi-organisational networks of participation was a strategic intention of both Welsh and Scottish perinatal networks and of the MCQIC team. Participants from both nations discussed horizontal networks of participation which linked strategic, clinical and managerial leadership, with staff on the shop floor expected to deliver the clinical intervention. In Wales, a national drive for perinatal optimisation was informed by PERIPrem and PReCePT methodology emphasising perinatal communities of practice, as evident in the activities of the Perinatal Optimisation Task and Finish Group, a sub-group of the National Strategic Clinical Network for Maternity and Neonatal Services, which aimed to deliver PERIPrem components in Wales, albeit without any allocated funding or operational capacity. This group enrolled local clinical, nursing and midwifery leads as perinatal optimisation champions tasked to drive change and improvement in their units. Local champions had the opportunity to share learning and access support during three monthly meetings. Annual network audit and QI meetings also provided platforms for local teams to discuss the intervention. Quality improvement capacity-building and support was the remit of Improvement Cymru; in 2023 funding was secured and actors with a vested interest in quality improvement and perinatal optimisation (i.e. the Clinical Network, Welsh Government, and Improvement Cymru, among others) were brought together to implement PERIPrem Cymru, and MATNEO SSP Cymru but because of the early stages of these networks’ activities, the relational work taking place within them could not be identified in this study.

In Scotland, cognitive participation activities specific to MgSO4 were led by the Maternity and Children Quality Improvement Collaborative as part of PPWP implementation, and these included MCQIC team visits to Health boards, webinars, and national meetings, organised during the first phase of the programme but discontinued during a later phase. Responsibility for overseeing implementation was then taken over by Health Boards. In recent years, the Scottish Perinatal Network is another platform bringing together maternity and neonatal staff, but without a quality improvement remit; other actors such as NHS Education for Scotland also have a quality and improvement role.

The merging of the maternity and neonatal networks in both nations demonstrated a cultural shift in perinatal teamworking. Both nations’ current strategies aimed to address collaboration between all stakeholders, influenced by the perinatal teamworking approach demonstrated to be an effective improvement strategy by PERIPrem and PReCePT, and which was also a BAPM policy priority. All study participants acknowledged the need for collaboration, aligning strategic priorities and clarifying roles and responsibilities for all organisations, networks, and teams with a perinatal optimisation agenda, and these aims featured in the activities of their networks, as the following excerpt illustrates.

> There’s a Scottish Perinatal Network which has been in place for about three years, four years now, […]MCQIC or SPSP Perinatal now are really trying to bring us all together. […] that’s a big priority, aligning the national stakeholders, within and out of Scotland to make sure we’re singing from the same hymn sheet, that these are MCQIC priorities and with individual variations if need be. But most definite alignment to be had there. (P13S, Neonatology)

Even though actors now had opportunities to make connections as part of perinatal networks, they still faced challenges in engaging in meaningful relational work to develop these into perinatal communities of practice where MgSO4 interventions could take shape. An ingrained culture of disciplinary boundaries in how care was organised and delivered exacerbated fractured communication and information channels and hindered relational work, as the following excerpt illustrates.

> It’s the culture thing, that this is your job. I don’t need to know this. This is my job. You don’t interfere. I don’t interfere. But that is a full pathway for the patients who have all experienced the journey for the mother and the baby from coming to the hospital to leaving the hospital and it is touching obstetrics, midwifery, neonatal, nursing, everything. People are not mature to see it from that point of view. We are all putting our blinkers on and this is what I need to focus on. Everything else is someone else’s job (P07W, Obstetrics)

The dominant culture shaping perinatal teamworking in the clinical area was mirrored in the obstetric-neonatal relationship dynamics within clinical leadership of the networks; Welsh participants believed the network was driven by neonatologists, and addressed neonatal priorities and conventions. These relational dynamics shaped obstetric motivation to participate and engage in these networks. In practice, obstetric participation was shaped by capacity, as well as culture. Staffing pressures and difficult working conditions exacerbated by the COVID-19 pandemic resulted in heavy workloads, and clinicians’ time away from clinical work was limited, which inevitably resulted in rationing engagement with and participation in relational work, such as network meetings. When obstetricians felt perinatal meeting agendas were shaped by neonatologists and addressed neonatal interests, participation became even more challenging, as illustrated in the following excerpt.

> Time that we can give to attending meetings, is limited. […]So we have to pick and choose which one is the most important where I definitely am needed and I will have to prioritise those meetings. So if I go to a PERIPrem meeting where there is only two obstetricians and 12 neonatal consultants and 12 advanced neonatal practitioners and they are talking in terms which I am not familiar with certain things I’ve never heard of, so next time I will think maybe I am not needed there. So next time, when I have to prioritise, I will say, okay, I am probably not needed there. (P07W, Obstetrics)

In Wales, participants also raised maternity and neonatal service commissioning structures as another hindering factor for perinatal collaboration as the participant in the following excerpt reports:

> Neonatal services are a commissioned service and the commissioning body in Wales is something called WHSC, so it’s the Welsh Health Services Commissioning and they commission specialist services but that doesn’t include maternity services. So, there is a complication with that, as a starting point. (P01W, Neonatology)

Maternity and Neonatal services are funded through different mechanisms and neonatal services are not allocated funds according to case load or size. Units with larger numbers of and/or more expensive service users would then inevitably have less funds to implement interventions that would help with quality improvement work, such as data managers. This was thought to inequitably allocate resources between services since units caring for more complex cases received the same funds as smaller ones. At the same time, because maternity received funding centrally, through Health board allocated funds, it was constantly competing with other departments with a higher profile, such as surgery and cancer services which might make it a lower priority for funding compared to these high profile services. Wales was reported to be going through a process of reevaluating commissioning systems and processes which was hoped to result in more appropriate allocation of resources that would enable meeting optimisation and care standards.

Most importantly, participation in implementation activities was reliant on individual enthusiasm. Organisations had no financial incentives for improving performance and quality improvement was led by individuals rather than implementation networks; clinical networks have no scrutiny or enforcement powers, and change is reliant on individual vision and leadership. Settings differed in their improvement capacity and capability (actors’ access and ability to draw from resources necessary to effect improvement), which further created disparities as smaller units were believed to always be at a disadvantage. Enrolling champions responsible for local implementation, and mentored by implementation leads, with protected funded time was raised by all participants as a core enabler, and an explanatory factor for the success of PReCePT and PERIPrem. A strategic intention of both nations was to enrol champions with funded time to deliver on optimisation initiatives, mirroring PReCePT methodology:

> The difference with PERIPrem is that we were doing all this unfunded through the network, so people were doing it as additional roles, and the main challenge that we have had is that the champions have been slightly inconsistent. It’s almost been different people on different meetings, and that’s been one of the biggest challenges. (P03W, Neonatology)

> When MCQIC was in first commission we used to have MCQIC midwives, and we used to have people that had paid time to be able to do the MCQIC work (but this was phased out), there may be people within the medical team that might have a bit of time for QI, but there’s so little time given to anyone on the front-line to be able to do any of the work, or any of the QI work at all. […] if only we had funding because I know that the PERIPrem model would maybe work similarly funded midwives, champions, and named leads within the centres. (P13S, Neonatology)

### Comparisons between the devolved nations and English experiences

A shift in perinatal teamworking culture was demonstrated by the merger of the maternity and neonatal clinical networks in both nations, which provided a platform where midwifery, obstetric and neonatal teams could interact and discuss the intervention. Even though other actors such as improvement agencies and in the case of Scotland the MCQIC team were also invested in improvement and perinatal optimisation, it is only in recent years, and following the evident success of PERIPrem and the PReCePT methodology, and the publication of BAPMs perinatal optimisation and perinatal teamworking toolkits, that both nations’ strategic intentions materialised into activities aiming to create perinatal communities of practice around the intervention. In practice, an ingrained culture of silo working, capacity issues i.e. staffing pressures and limited time away from front-line clinical duties, as well as motivation and competing priorities, intersecting with perceptions of neonatologists controlling the narrative, challenged equal participation by all actors in these communities of practice. Commissioning mechanisms and organising structures further ingrained boundaries and divisions between specialties. Lack of protected time also impeded champion continuity, with the role being occupied by different clinicians over time which hampered meaningful relational work and the development of communities of practice.

In England, the Clinical Negligence Scheme for Trusts, and the operational and delivery remits of clinical networks provided strong incentives for all actors, including local organisations and teams, to prioritise and commit to MgSO4 uptake improvements, and provided platforms for networks of participation. Their importance for driving commitment and participation were evident in the PReCePT evaluation findings. Benefiting from these drivers, the PReCePT methodology supported cognitive participation through organising implementation around PReCePT networks of participation and communities of practice which spanned all layers of the system through:

**1. Funded backfill time and protected time away from clinical duties** PReCePT evaluation highlighted how the provision of adequate backfill time and protected time away from clinical duties for champions was one of the most important implementation strategies. This allowed champions time and space away from clinical duties to participate in network activities and engage in relational work with other champions, (especially obstetric and neonatal champions in the case of ES) which allowed communities of practice to take shape.
**2. Champion communities, which allowed relational work to take place between actors from different Trusts and in the case of ES, maternity and neonatal teams** In the case of the NPP, these involved midwife champion networks, where champions within each AHSN were linked to each other, and to AHSN QI leads, and regional clinical leads for implementation support. The most successful form of these networks were those taking place digitally in WhatsApp groups. All implementers were also part of Facebook and Twitter (now X) online communities where they shared news, learning, and success stories. ES units had perinatal local implementation teams, and in addition to having a named clinical champion, clinical champions were allocated backfill funding and protected time to participate in learning events alongside midwife champions. Significant relational work took place during these events which resulted in stronger perinatal teamworking in these settings, compared to those not receiving the ES package.
**3. The clinical micro- and meso-systems as communities of practice:** A clinical microsystem is understood as a small group of professionals working together to provide care to discrete populations of patients, e.g. the perinatal team, whereas the clinical mesosystem is combination of these teams which make up the service e.g. the maternity care service. PReCePT methodology encouraged these groups to work together and improved communication which contributed to relational work to come together as a community of practice. Champions were encouraged to use PDSA cycles to effect change in their settings, and this required involvement from all members of the perinatal team. Embedding PReCePT in inter-professional training helped establish conversations around the intervention as part of routine opportunities coming together formally, for example in perinatal mortality and morbidity meetings, handover and huddle meetings, or informally as part of informal conversations on the ward.

Table 12 summarises the challenges to cognitive participation, the strategies adopted by PReCePT to address these challenges, and the lessons learned through our research on what a QI intervention implementation framework should include to ensure collective coherence of intervention implementers and adopters.

**Table 12:** Cognitive participation: Overview of findings and lessons learned.

|  | Challenges to relational work | PReCePT strategies | Lessons for a QI intervention Implementation Framework |
| --- | --- | --- | --- |
| Cognitive participation | Multiple QI & optimisation actors working in isolation<br>Midwifery, obstetric & neonatal team relationships & teamworking culture across micro, meso, and macro levels of the system (silo working, different specialty cultures & hierarchies)<br>National & local implementation led by neonatal teams means obstetricians not always motivated to participate in implementation networks<br>Commissioning mechanisms, immense staffing pressures, poor working conditions | An Engagement and Awareness Raising Strategy :<br>Enrol implementers with the right roles, experience, and motivation from across the system (e.g. AHSNs, clinical networks, local teams)<br>Clarity of implementation roles<br>A perinatal team of champions<br>Connect PReCePT to all MgSO4 stakeholders & networks e.g. MATNEO SIP networks<br>Participation in social media platforms<br>Align PReCePT with national context & use other networks and collaboratives as vehicles for communication<br>Bring implementers together as part of communities of practice (physical and online)<br>Engage all teams across the care pathway in relational work e.g. community midwives, pharmacy, anaesthesiology<br>Backfill funding for champions | Building relationships around the intervention a core part of set-up phase to create networks:<br>Regional scaling structures/teams (QI and clinical leads)<br>Local implementation teams (midwifery, obstetric & neonatal leads)<br>A network of participation linking regional and local teams<br>Linking implementation teams to other wider associated networks & executive sponsorship in each setting<br>Enrolling key organisation staff in supporting implementation roles (e.g. clinical leads, strategic and managerial leads)<br>Use networks to communicate intervention message<br>Multimodality platforms where networks can socialise<br>**Shared coherence needed for commitment to participation**<br>**Champions with funded protected time** |

#### Collective action

Collective Action refers to the operational work that people do to support a new practice. Collective action in this study refers to what members of the perinatal team do individually and collectively to ensure MgSO4 is administered and data are captured accurately. Crucially, it requires the appropriate allocation of tasks. MgSO4 is prescribed and administered by members of the maternity team, but is recorded and captured in neonatal patient record systems i.e. Neonatal Badgernet. For these two activities to happen, midwifery, obstetric and neonatal staff need to engage in a series of articulated tasks to identify women at risk of, and diagnose preterm labour, prescribe and administer MgSo4 within a given time window, and capture its administration in the neonatal patient record system Badgernet:

> The administration of the magnesium sulphate is done by the midwifery staff on the instructions of the obstetricians. Those conversations (are had) between the obstetricians and the neonatal team. So, it might be me that the obstetric team are dealing with. So, when it looks like delivery is imminent, as well as talking about antenatal steroids I will talk to the obstetric team about whether they think there’s time and whether they think it’s indicated to give magnesium sulphate to a pregnant lady. (P06W, Advanced Neonatal Nurse Practitioner)

Our study suggests that commissioning decision-making mechanisms, and policies determined the resources available to units, for example access to neonatal expertise and cots, equipment e.g. prefilled syringes, number of staff etc. Access to resources shaped the challenges teams faced in improving their uptake rates. Smaller units were believed to be at particular disadvantage because of lacking in neonatal resource, and having to organise in utero transfers; a time consuming and logistically challenging task, in addition to delivering care to women. Cross-organisational communication and collaboration to clarify referring units’ concerns and working to the same guidance were important for ensuring MgSO4 was administered before transfer, but in most cases participants agreed MgSO4 was not administered. Challenges included the uncertainties in diagnosing preterm labour as the following excerpt illustrates:

> We have a gestational cut-off that we can accept, there sometimes is a difficult conversation with the obstetricians where they have got a lady on the antenatal ward. Say she’s 24 weeks. And they say, ‘She’s got a urinary tract infection. She’s not in labour.’ Or, ‘She’s got diarrhoea and vomiting. She’s not in labour. She will be going home tomorrow.’ So, then that lady doesn’t get steroids necessarily. She doesn’t necessarily get magnesium sulphate and nor does she get transferred to a unit who could cope more effectively if the baby was delivered. And then what happens is that that lady’s UTI was early labour and this lady is now in established labour and of course then they do think to give steroids and to give magnesium sulphate but it might be a little bit too late. And it’s definitely too late to try and do an in-utero transfer. (P06W Advanced Neonatal Nurse Practitioner)

But the competing priorities staff had to navigate when trying to organise transfers also affected administration during in utero transfers. One Welsh participant described how magnesium is usually missed during in utero transfers because their team lacked in confidence to decide whether the intervention was warranted, and administering it appropriately. During high pressured scenarios with competing demands on staff, more immediate, familiar tasks took precedence as the experience of the following participant illustrates:

> We don’t want to be transferring people to tertiary units having given them the loading dose of magnesium sulphate with our decision for delivery when potentially they may get to the tertiary unit and that isn’t the case, they don’t think that delivery is necessary. […] I just think that we maybe haven’t been very good at it because our biggest thing would be to transfer them to the best place for their birth and that obviously takes priority along with steroids, antibiotics, etc, etc. (P08 W, midwife)

Individual and collective coherence as well as skills and competencies were essential. High staff turnaround however had a negative impact on capacity building among staff, and necessitated more intensive and structured learning opportunities to ensure every new member of staff was trained; again smaller units were believed to be at a disadvantage:

> I think the differences in sizes of units is sometimes always a bit tricky because you have so much more resource throughput in the big tertiary centres and the smaller centres may just be lacking in some… I don’t know, I don’t want to say skills, but just the resources to be able to do everything as efficiently as a bigger unit who’s doing it much more frequently. The smaller units where it’s infrequent, it’s harder to get it all as ingrained as in tertiary units..(P03W, Neonatology)

Even if senior members of the obstetric and neonatal teams were familiar with the intervention, it was junior staff and other members of the multi-disciplinary team who determined whether women received the intervention, and without individual skills and collective coherence, this was jeopardised during highly pressured and stressful situations. Clarity in the guidance as was the case for births below 30 weeks were more likely to receive MgSO4, but more complex cases, particularly in pregnancies over 30 weeks where individual clinician judgement was required made it more challenging, as the following excerpts illustrate:

> I feel that baby can still benefit from MgSO4 but because people on the ground sometimes interpret guidelines literally and that can sometimes lead to loss of the bigger picture and where it is said give, they give. Where it says consider, especially junior staff, and remember a lot of the time junior staff will be involved in the emergency management […] because sometimes in emergencies it’s a bit stressful and multitasking and other things and you can lose the bigger picture. (P07W, Obstetrics)

> Sometimes our anaesthetist or midwives will stop the magnesium when a woman’s going to theatre. And then it might be 45 minutes later when you’re like, ‘Where’s the magnesium? Should be running.’(P05W, Obstetrics)

Cognitive load therefore, i.e. taking decisions in the context of clinical uncertainty, and trying to find information and equipment at the same time was a crucial factor impacting on administration, making smoothly articulated clinical workflows important. Study participants discussed the role of aligning national and local guidance to ensure all units, teams and clinicians worked to the same protocols which decreased behaviour variation. In addition to policies, easy access to workflow charts, and equipment such as pre-filled syringes were also instrumental as the following excerpt illustrates:

> We have to make it as easy as possible for them. We have very straightforward guidance to follow. We have very straightforward flowcharts that show you exactly how to make up magnesium sulphate and what the dose should be and what the dilutant should be and what rate you put the pump on at and all these very straightforward things that just make people’s lives easier (P09S, Obstetrics)

On a strategic level, existing policies and programmes championed by professional associations, particularly the British Association of Perinatal Medicine were mentioned by all participants to inform implementation and improvement. Such programmes and toolkits were backed by either evidence of effectiveness such as the PERIPrem and PReCePT intervention, or had the sponsorship and expert input of a perinatal network as in the case of the BAPM perinatal optimisation and perinatal teamworking toolkits. Participants from both nations referred to using these resources to make it easier for teams to remember and administer MgSO4, as the following excerpt illustrates:

> People saw the difference in performance in Wales compared to England and the obvious difference was that PReCePT happened […] that’s why we’ve just used the Peri-Prem resources because they’re available. If someone’s done the work of creating things, then we should be using them. (P02W, Neonatology)

### Comparisons between the devolved nations and English experiences

Study participants believed women below 30 weeks were in the large majority of cases receiving MgSO4, and maternity teams had the knowledge, skills and competencies needed to ensure adherence. There was more likelihood for pregnancies over 30 weeks, in utero transfers, difficult to diagnose preterm labour, and emergency C-sections not receiving MgSO4. Analysis highlighted specific challenges and enablers to administering MgSO4 which, in combination with the PReCePT evaluation findings, helped create a profile of factors and setting-specific attributes which jeopardised adherence.

Maternity units with access to neonatal resource emerged as more likely to administer MgSO4 because of a higher concentration of socio-cognitive (competencies, skills, confidence, and positive attitudes towards the intervention and its components) and socio-structural (e.g. clinical guidance, workflows and a perinatal optimisation culture) resources.

National preterm care pathways and policies such as “birth in the right place”, and commissioning mechanisms, defined the resources concentrated in and available to units e.g. distribution and number of NICU units, number of maternity staff, neonatal beds, and access to equipment such as pre-filled syringes. They also defined the sequence of tasks and activities taking priority in units e.g. mobilising maternity teams into organising in-utero transfer, rather than delivering perinatal optimisation interventions. Knowledge of, and positive attitudes towards the intervention are more common in neonatal teams who take responsibility for mobilising and advise maternity teams particularly when encountering challenging cases. There were more opportunities therefore for, at least informal, learning to take place due to high exposure to preterm labour which necessitated maternity and neonatal teams to work together. Without intervention awareness and clarity of process among all those involved in the care of women in preterm labour, MgSO4 was more likely to be forgotten during stressful and complex situations such as transfer of mothers to the operating theatre or other units. Good communication and information exchange, and collaboration between the members of the perinatal team within and across settings, for example in cases of in utero transfers, were crucial for overcoming specific challenges, but analysis suggested perinatal teamworking might not be as strong in smaller units.

The PReCePT evaluations demonstrated that the following intervention components were able to address barriers to administration:

- **The PReCePT Toolkit resources**: this was a coproduced and piloted resource described as flexible and easy-to-use. Champions used the included resources to e.g. embed MgSO4 in local protocols and pathways and other documentation, train all members of the perinatal team, and use resources such as posters and proformas to ensure MgSO4 was a perinatal team priority and at the forefront of everyone’s minds.
- **PReCePT Training and staff capacity building**: training and raising awareness among all members of the perinatal team. PReCePT enriched settings’ socio-cognitive resources by creating competencies and improved MgSO4 self-efficacy among all members of the perinatal team. It gave permission to all members of staff to take responsibility for MgSO4 administration decision-making (shared governance).
- **Clinical communities of practice**: Its cross-organisational and perinatal communities of practice meant that teams in smaller units with limited access to resources were able to draw from the resource and capital of larger units through diffusion of innovation, peer to peer support, and links to contacts in other hospitals they could use to coordinate and discuss difficult cases such as during in-utero transfers.
- **QI communities of practice**: Champions from poor-resource settings benefited from the support and mentoring of others to utilise QI techniques such as PDSA cycles, and use learning from missed case analysis to effect changes in their system which made it easier for their teams to administer MgSO4 e.g. PReCePT grab boxes. ES was more effective in creating QI resource within settings.

Table 13 summarises the challenges to collective action, the strategies adopted by PReCePT to address these challenges, and the lessons learned through our research on what a QI intervention implementation framework should include to ensure collective coherence of intervention implementers and adopters.

**Table 13:** Collective Action: Overview of findings and lessons learned.

| Collective action | Challenges to collective action | PReCePT strategies | Lessons for a QI intervention Implementation Framework |
| --- | --- | --- | --- |
|  | • Ambiguous clinical protocols<br>• No defined & articulated preterm labour pathways and workflows<br>• Awareness and knowledge of intervention among staff providing care<br>• Clinical skills and competencies associated with preterm labour e.g. diagnosis<br>• High pressure scenarios with competing priorities e.g. transfers<br>• Staff exposure to intervention, staff turnover are challenges to building critical mass<br>• Information flow within & between clinical teams involved in preterm labour | • Make decision-making & administration as easy and quick as possible<br>• PReCePT pathways and other resources in Toolkit<br>• MgSO4 message linked to steroids<br>• Aide Memoirs & Posters<br>• Open communication channels between staff on the floor, champions & clinical leads for “live” direction, support and liaising to organise administration<br>• Training all perinatal team staff<br>• Perinatal teambuilding as part of cognitive participation<br>• Highest impact for units low in resources | • Understand points in the care pathway where missed opportunities cluster<br>• Understand barriers and blockers to administration to inform intervention targets<br>• Provide resources to make decision-making and administration as easy and quick as possible (pathways, equipment, etc)<br>• Interprofessional training of all intended adopters |

#### Reflective monitoring

Reflexive Monitoring is the appraisal work that people do to assess and understand the ways that a new set of practices affect them and others around them, and how well they think they are working. In this case, appraisal work refers to outcome measurement and audit activities taking place to assess adherence to clinical guidance. Even though MgSO4 is administered by the maternity team, outcome measurement and audit is carried out by the neonatal team who are responsible for submitting data to the National Neonatal Audit Programme (NNAP); this creates a split between those at the centre of operationalising MgSO4 guidance, and those tasked with monitoring adherence to the guidance.

NNAP emerged as one of the strongest drivers of improvement initiatives. Up to that point MgSO4 was captured in neonatal patient records but these data were not routinely audited. The publication of annual NNAP data provided information to clinical leads on their unit’s performance, and enabled benchmarking against other settings. It worked synergistically with NICE clinical guidance and the existing evidence base to drive change initiatives; NICE guidance and clinical evidence provided direction on what units should be doing, whereas NNAP data provided information on adherence. That information was used by the Welsh perinatal clinical network to identify outliers and provide recommendations to Health Boards. The Scottish Government withdrew funding and contractual arrangements with NNAP in 2020 but has now re-entered the programme. Appraisal work is carried out by the MCQIC team using the data submitted by Health Boards participating in the PPWP.

Neonatal staff had responsibility for MgSO4 data entry, but all participants agreed the quality of data was often poor which presented challenges to appraisal work and quality improvement. Maternity and neonatal teams used multiple paper and digital patient record systems which lacked interfacing capabilities, and data imputing into neonatal Badgernet relied either on manual data transfer, or communication between maternity and neonatal teams, for example during handover meetings. In addition to Badgernet, QI programmes such as the PPWP required neonatal teams to enter the same data in a separate data system (part of the Core Measurement Plan), which added further complexities to data capture, as the following excerpt illustrates:

> When we admit the babies it will go into our Badgernet Maternity, pulled into Badgernet Neonatal — actually it’s not an automatic process, at the moment it’s a manual process.

> However, what SPSP looks at is an external toolkit spreadsheet that sits in a dashboard on our computers, that we transcribe into a spreadsheet manually, so user error or input error and all of these sort of things they definitely play a part. (P13S, Neonatology)

Because NNAP and PPWP used neonatal data, appraisal work was routinely concentrated within neonatal teams. However, the findings needed to be communicated to maternity teams for change to happen, and the nature of this communication often defined its success. A shared strategy between the two nations was to create opportunities for maternity and neonatal teams to share learning on existing platforms e.g. perinatal network activities. Participants described opportunities for maternity and obstetric teams to discuss MgSO4 adherence for example perinatal mortality and morbidity meetings, but feeding back data to all members of the perinatal team in a meaningful way, was not the norm and relied on local initiative and capability including perinatal teamworking culture and QI resource. Another challenge was that NNAP represented data obtained from the year before, which was not as meaningful and informative for local teams because of the time delay. Performance may have improved at the time of publication, but staff reverted back to the normal routines when MgSO4 was superseded by other priorities. Feeding “live” data to actors across teams and hierarchical structures was thought of as important. Again, tools and resources aligning with PReCePT methodology were adopted to achieve that, as the following excerpt illustrates:

> I still feel as if there’s a lack of sharing (MCQIC) data, in a bit more of a meaningful way to front-line teams […] what we started to do locally is generate a […] poster. […] it’s found to be a bit more meaningful to teams. They definitely respond a little bit more to something a bit more visual than a run chart. (P13S, Neonatology)

Data capture and performance monitoring were described as time consuming and labour intensive, which presented additional challenges to quality improvement work, particularly when QI programmes required teams to enter the same data into two different databases or prepare detailed progress reports. As in the case of administering magnesium, data capture needed to be easy in order for teams to carry out the task successfully. This was a strong learning point emerging from the Scottish interviews when reflecting on the challenges faced by the SPSP team. Scottish participants described how the MCQIC team was redesigning and simplifying their data collection processes in light of the lessons learned from the first phase of the programme, as the following excerpt illustrates:

> Something that we’re working on within the new programme is to get rid of this duplication of measurement, and actually pull it off from the one source that it’s all going into, and that’s definitely a priority that we’ll hopefully take forward with the new part of the programme. (P13S, Neonatology)

All participants agreed that resource and capacity were required for such activities to take place. Analysis highlighted how quality improvement activities, including audit and missed case analysis, were led by individuals on a voluntary basis, which jeopardised continuity and embeddedness of the work, for example when staff moved on to different roles. Access to QI training was open to all interested staff, but this did not come with protected time. Participants agreed that without protected time and space, and support from their teams and organisations, such efforts would not be as successful or sustained, as illustrated by the following excerpt.

> Most of the QI are led by individuals depending on their initiative, their motivation, their enthusiasm and depending on the support they receive from their own department,[…] QI cannot flourish amidst hostility. If I am trying to do a QI and all of my colleagues think that is rubbish, I don’t think it will get off the ground. So that is why it’s very, very important. […] to have the time. If I’m over burdened with my clinical work I won’t have the time. (P07W, Obstetrics)

### Comparisons between the devolved nations and English experiences

Findings confirmed PReCePT evaluation findings that existing outcome measurement and performance monitoring processes are not conducive to reflexive monitoring activities and quality improvement work. Badgernet data quality is low, and data are not fed back to the perinatal team in a way that promotes learning and behaviour change. NNAP data target a neonatal audience, and the annual feedback mechanisms is not as meaningful and informative for driving improvement. The quality of perinatal team communication and information flow between the two teams further compound the challenges presented by multiple and distinct maternity and neonatal patient records. Audit and feedback, and missed case analysis are reliant on individual clinical capacity and capability, local culture and processes, which puts implementation and sustainment of these activities at risk.

PReCePT addressed these issues through its emphasis on “using data for improvement”, continuous monitoring of improvement and evaluation, of process, as well as outcome. It was generally agreed that improved quality of data through improvements in data capture and missed case reviews was one of the factors underpinning improved uptake rates; in many of the cases triangulating neonatal and obstetric data illustrated that MgSO4 had in fact been given though not reported. PReCePT components which enabled reflexive monitoring included:

- **Improving data quality:** providing tools such as proformas, which improved data capture by maternity, and made data entry easier, and more accurate for neonatal staff. Data quality was also improved through perinatal team building
- **Audit and Feedback:**

`o` Baseline measurement was used to gauge improvement
`o` Dashboards and posters made it easier to translate data into easy to understand visual representations of performance presented in wards weekly, so all staff and patients could see “live” performance information
`o` Champions embedded MgSO4 discussions in all informal learning encounters in the ward, discussing performance and learning from missed case analysis with staff
`o` Champions and clinical leads also presented this data in routine meetings where strategic decisions on improvement needs could be made
- **Champions** with protected time and space to engage in reflexive monitoring activities
- **Continuous quality improvement:**

- Encouraging units to continually monitor and review performance to understand cases likely to not be given MgSO4 and use the learning to guide further improvement activities
- **QI collaboratives:** being part of communities providing opportunities for QI coaching, sharing learning and innovation, and accessing feedback and support helped champions create competencies, boosted morale, maintain momentum, and supported local implementation. PReCePT communities of practice, in combination with other QI programmes such as MatNeo SIP and operational delivery networks were platforms were PReCePT learning was discussed, which helped embed the intervention in improvement activities across the system further helping with sustainment beyond the end of PReCePT.

Table 14 summarises the challenges to collective action, the strategies adopted by PReCePT to address these challenges, and the lessons learned through our research on what a QI intervention implementation framework should include to ensure collective coherence of intervention implementers and adopters.

**Table 14:** Reflexive monitoring: Overview of findings and lessons learned.

|  | Challenges reflexive monitoring | PRCePT strategies | Lessons for a QI intervention Implementation Framework |
| --- | --- | --- | --- |
|  | • Intervention operationalised by maternity teams, but performance appraised by neonatal teams<br>• Badgernet data quality and meaningfulness<br>• Problematic IT and communication systems (informal ways of acquiring MgSO<sub>4</sub> data)<br>• Missed case analysis not a requirement<br>• NNAP publishes “outdated” data not useful for improvement<br>• Data discussed primarily among neonatal teams; do not trickle down<br>• Individual initiative behind activities; precariousness of QI work<br>• Collecting data for NNAP and other QI programs complex and time consuming, relying on individual initiative | • Improving quality of data/information flow between mat&neo teams<br>• Toolkit resources e.g. dashboard, stickers, proformas etc<br>• Teambuilding<br>• Audit and feedback;<br>• Champions<br>• Taking advantage of or creating opportunities for discussing intervention within settings e.g. handover (cognitive participation)<br>• Opportunities to discuss the intervention across all levels of the system<br>• PRCePT communities of practice, MatNeo SIP, learning systems & within organisation<br>• QI capacity building among champions how to use data for improvement | • Building relationships around the intervention a core part of set-up phase to create networks:<br>▪ *Regional scaling structures/teams (QI and clinical leads)*<br>▪ *Local implementation teams (midwifery, obstetric & neonatal leads)*<br>▪ *A network of participation linking regional and local teams*<br>▪ *Linking implementation teams to other wider associated networks & executive sponsorship in each setting*<br>▪ *Enrolling key organisation staff in supporting implementation roles (e.g. clinical leads, strategic and managerial leads)*<br>• Use networks to communicate intervention message<br>• Multimodality platforms where networks can socialise<br>• **Shared coherence needed for commitment to participation**<br>• **Champions with funded protected time** |

### 4.4. Implications for future perinatal optimisation activities: A blueprint for a QI implementation framework

The PReCePT Devolved Nations findings add to our understanding of the process of implementation and scaling of QI interventions, challenges and enablers of implementation, and implementation outcomes ^8^ ^22–24^. Our comparisons between two different packages of implementation support delivered to units as part of the PReCePT study enabled us to understand what components of the support packages are essential as a minimum, and when additional support input might be needed to create capacity and capability equitably within units to drive behaviour change across the board i.e. address disparities between units in the quality of care delivered. Disparities between units made evident in pronounced differences in uptake rates between units can go unnoticed when performance monitoring focuses on national average rates, but the implication is a postcode lottery of quality of care which compounds regional inequalities in health outcomes. Our findings have shed light on the main contextual enablers shaping settings’ readiness to implement framework activities (Figure 14). By comparing findings from these three evaluations, we can propose a framework for future national perinatal optimisation programmes to accelerate getting research evidence into clinical practice. This framework, presented in Table 15, includes activities and resources that should be considered across all levels of the perinatal ecosystem (national context, regional health system, organisations, multidisciplinary care team, the perinatal clinical team, and patients/service users) to address four primary drivers of implementation as described in the PReCePT implementation guide^25^:

1. **Engagement with all actors and Awareness Raising**: This is the preparatory work that needs to be done to prepare the ground for implementation. It requires communicating a concise, clear message to all stakeholders including service users, enrolling implementers and champions with protected, funded time, and understanding implementation readiness and implementation support needs across settings. Creating communication networks that link clinical leadership to staff on the shop floor is part of this phase.
2. **Knowledge mobilisation:** This involves creating capacity and capability among implementers so they can design local implementation plans to scale the intervention of interest. Creating improvement capacity in the system, and competencies specific to the clinical intervention among adopters are part of this phase.
3. **Operational and system enablers:** Adopting, adapting and developing practical resources from piloted and tested Toolkits to make decision-making and engaging in different behaviours as easy as possible for staff e.g. clinical pathways, workflows, clinical proformas, equipment etc.
4. **Driving Behaviour Change by embedding knowledge into practice:** Using national and local data and information systems to continuously monitor performance, and feeding back the learning into the system through audit and feedback mechanisms, educational activities, and governance processes.

**Figure 14:**
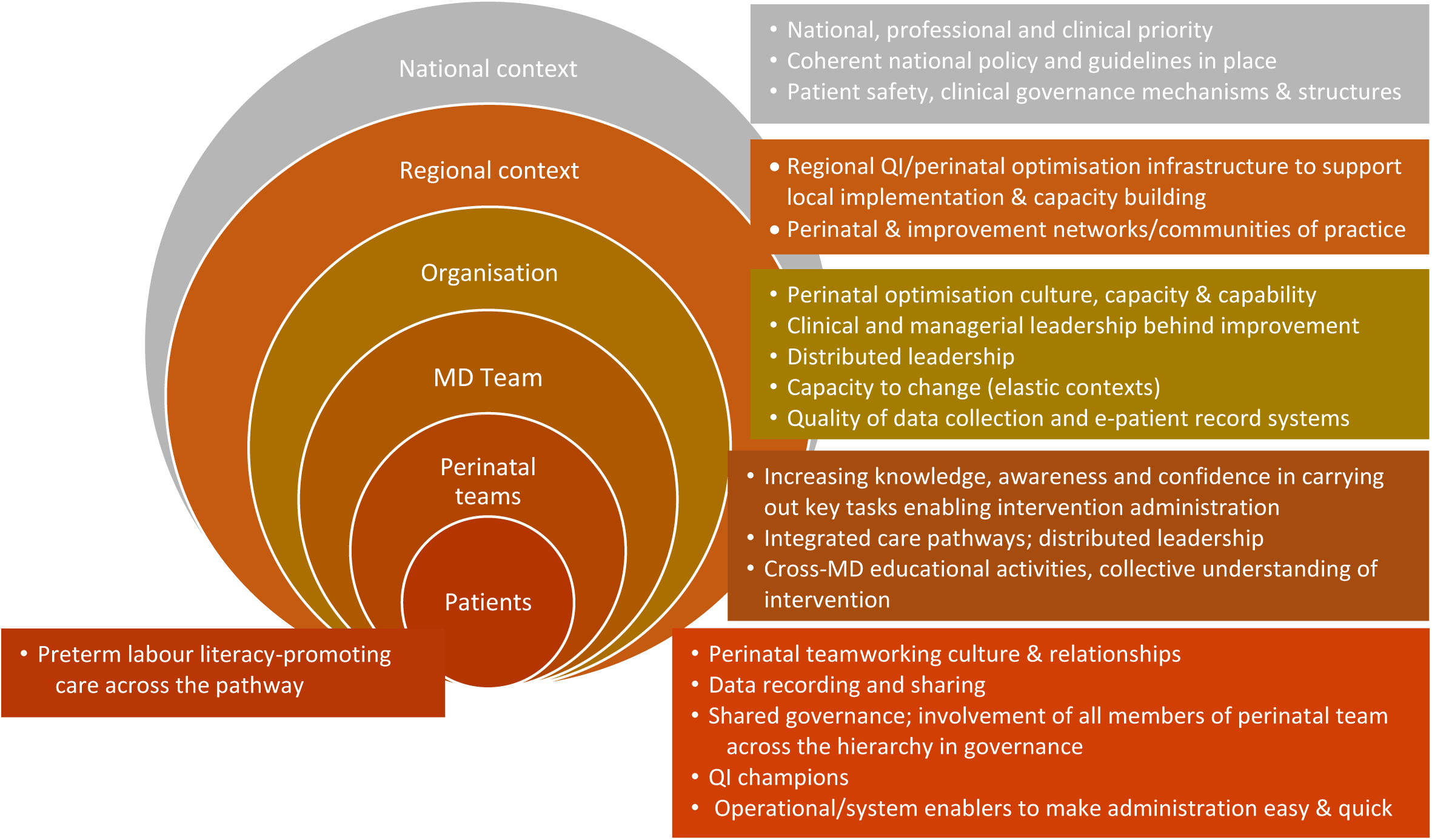
Contextual drivers and enablers which foster improvement across the perinatal ecological system

**Table 15:**
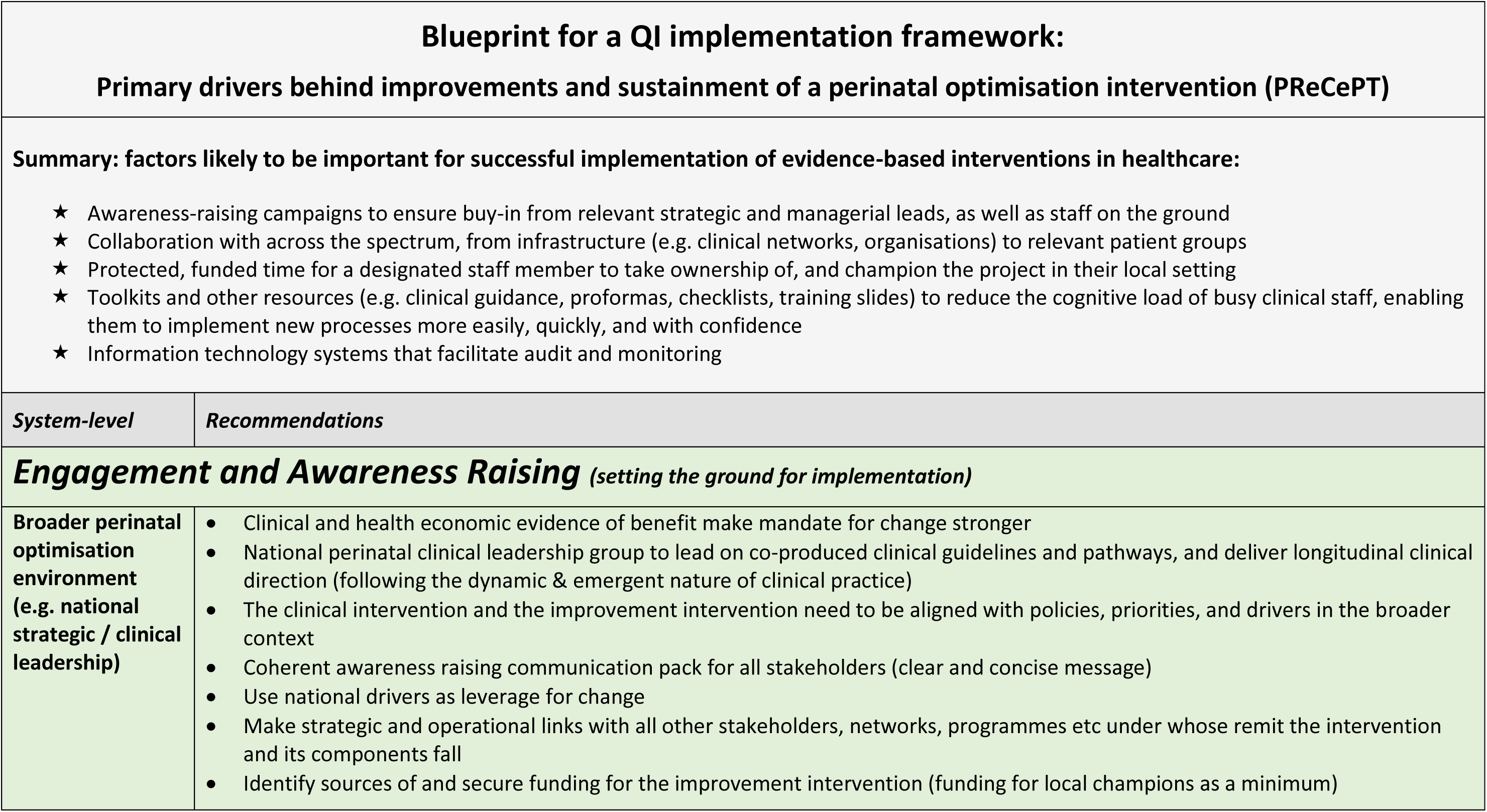
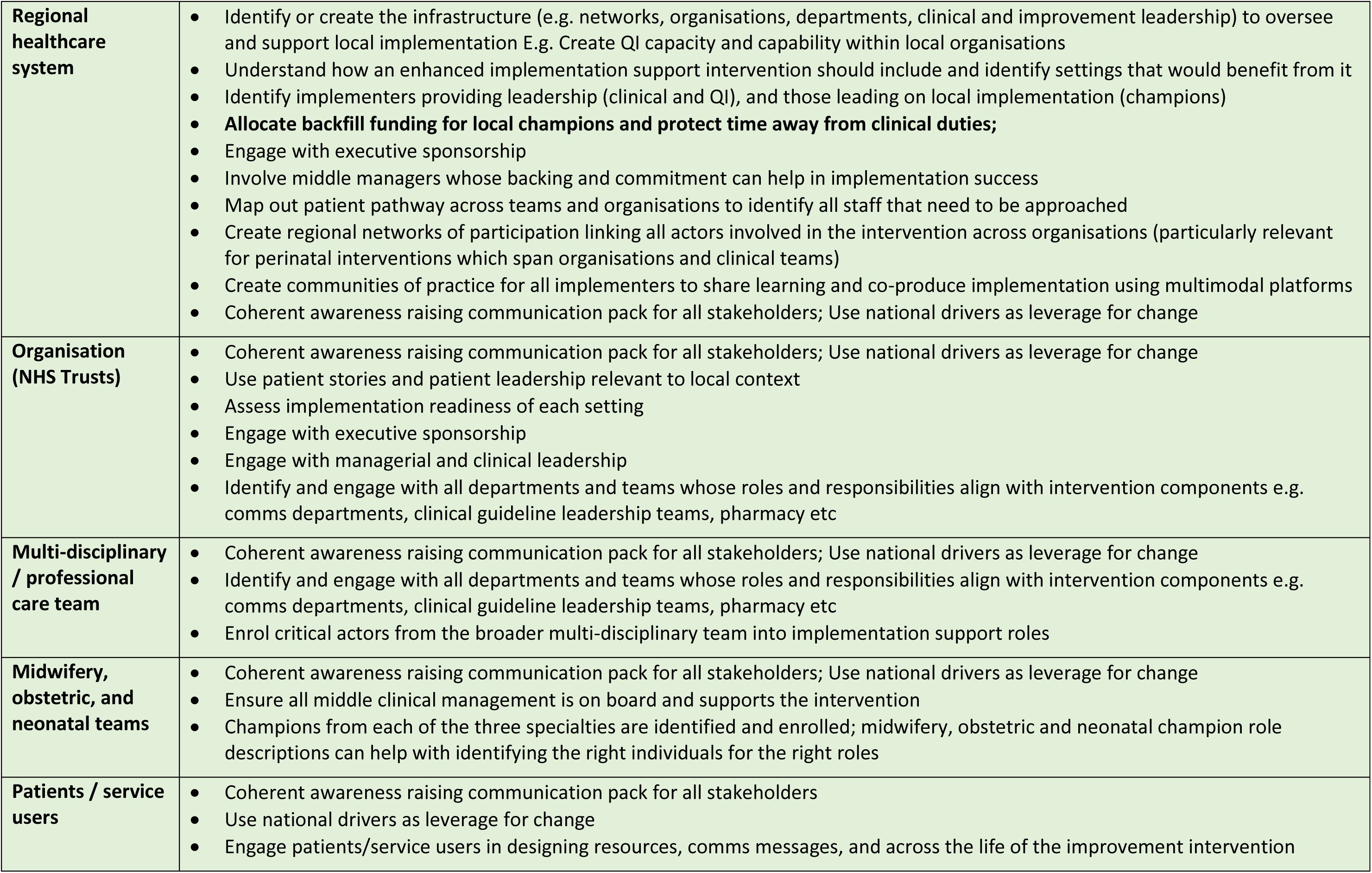
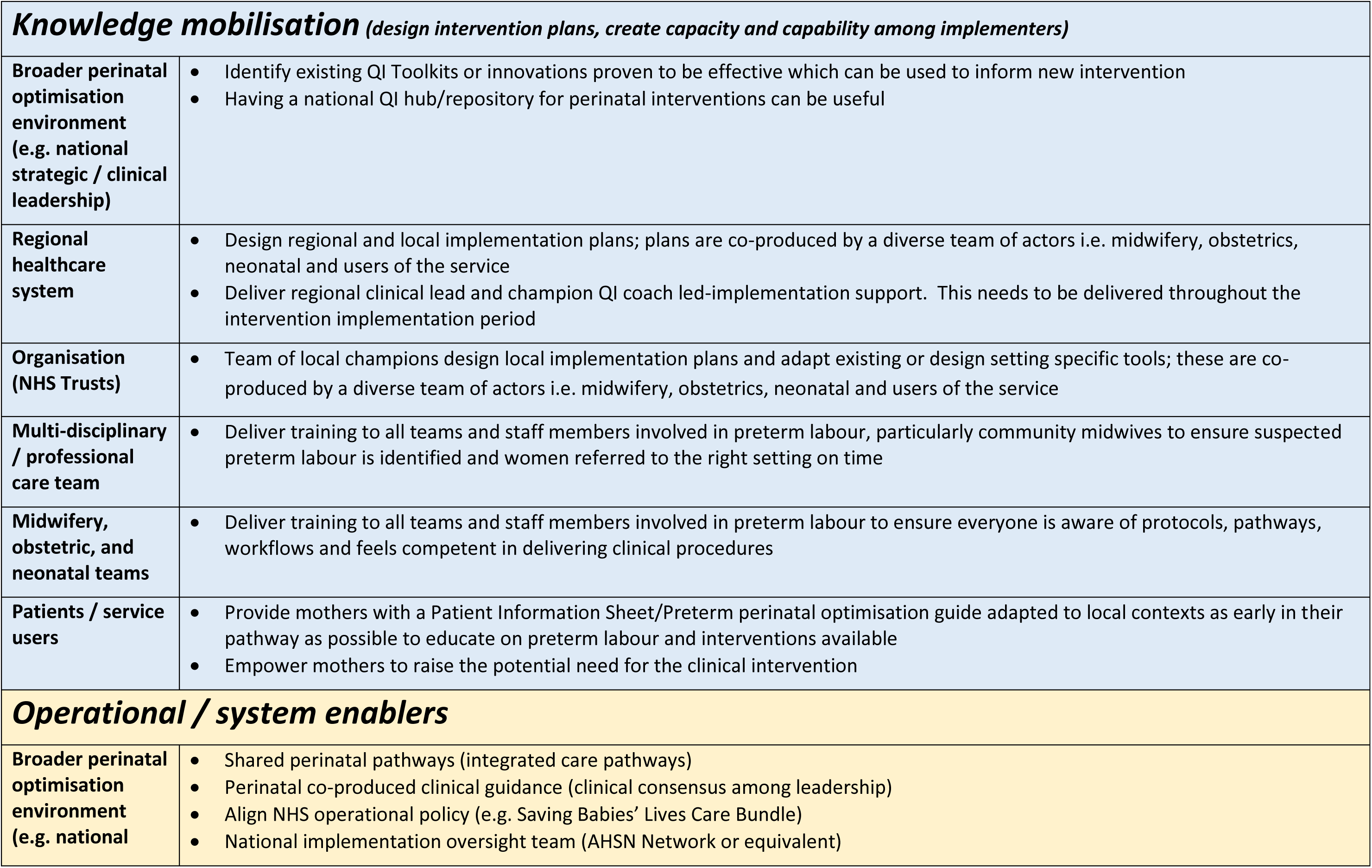
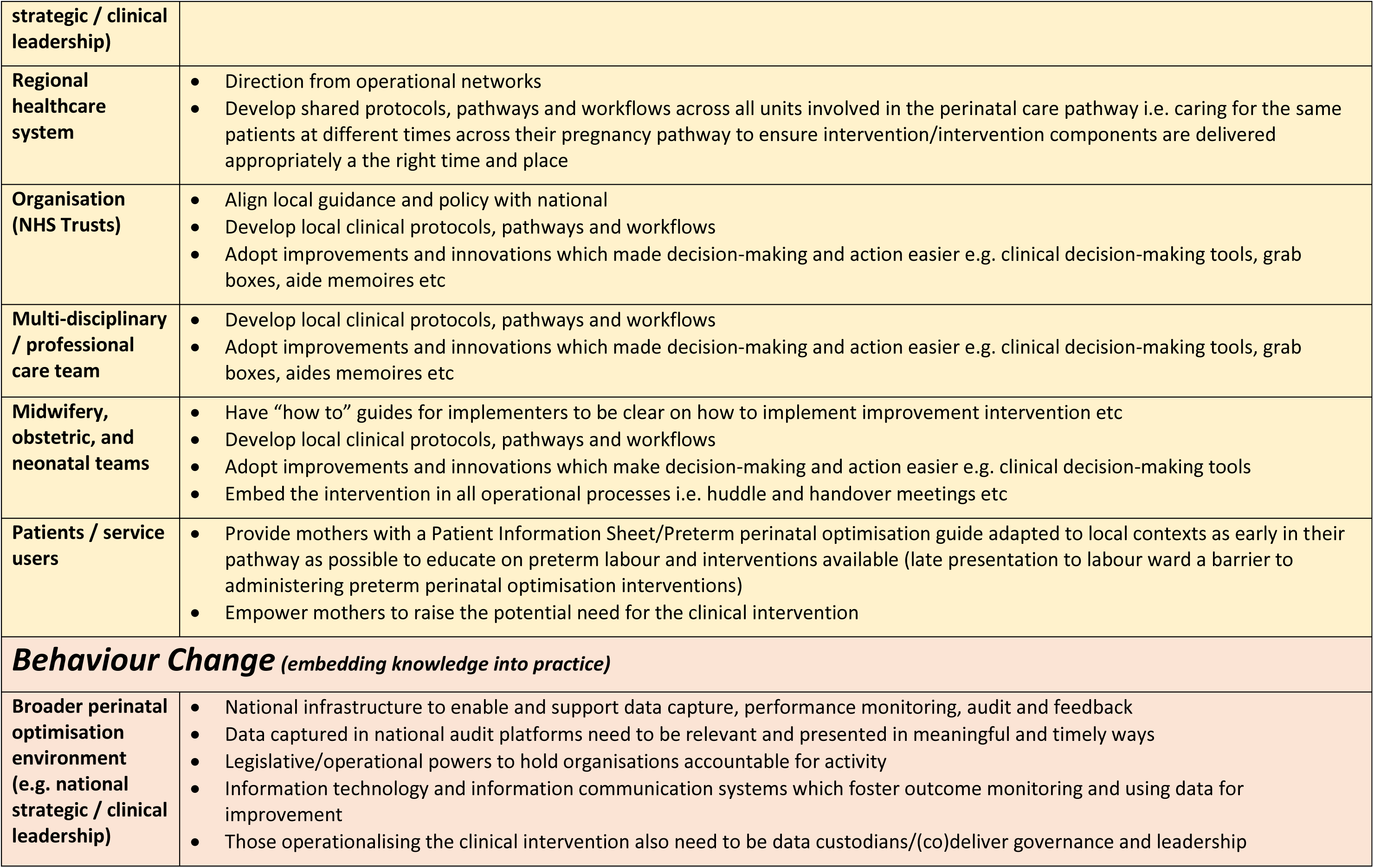
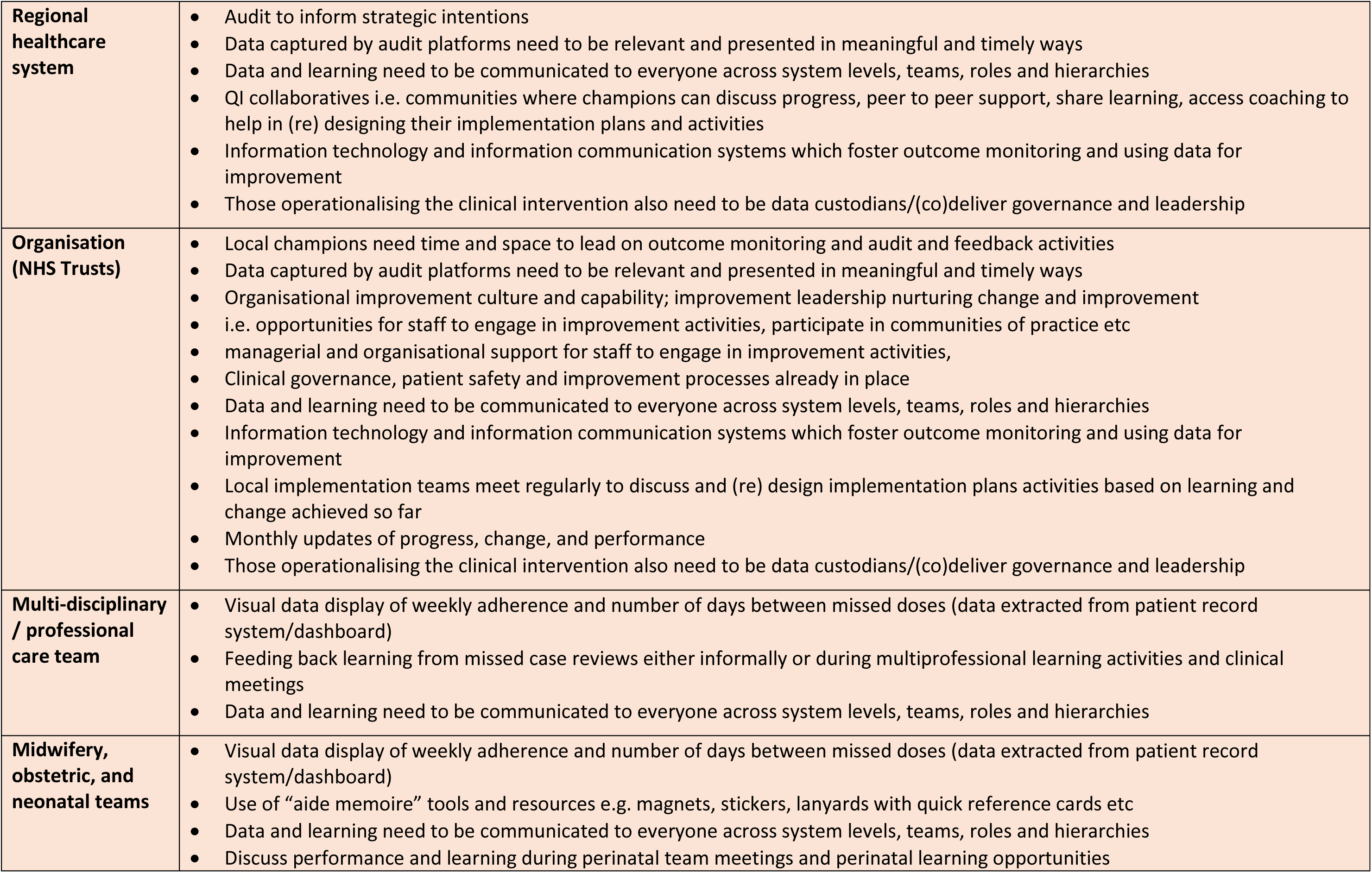
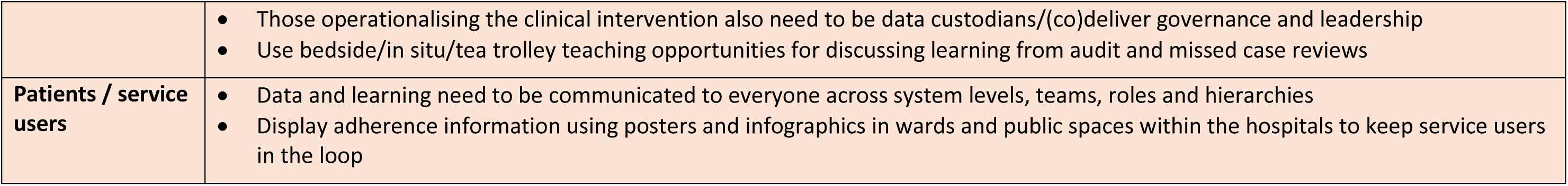
A blueprint for a QI implementation framework.

## 5. Discussion

### 5.1 Summary of results

In England, there was evidence of an increase in MgSO4 use following the launch of the NPP, with the majority of gains appearing to take place in the first year or two. The improvements achieved were broadly sustained over the four-year follow-up, although there is indication of a more recent decline in use, consistent with the COVID-19 pandemic period. Completeness of MgSO4 data in hospital records has been sustained with under 1% missing MgSO4 data.

Nationally, the greatest overall improvement over time was seen in Wales (due to their lower starting levels). Compared to the devolved nations, uptake appeared to improve faster in England after the NPP launch. In the latest 2022 data, the three nations were broadly comparable with delivery of MgSO4 around 81-87%.

At the individual level in England, historically, mothers in the North of England, or with a history of smoking, were less likely to receive MgSO4 compared to mothers in the South or with no smoking history. While the North/South disparity in treatment appears to have attenuated, with little difference soon after the launch of the NPP, smokers remain less likely to receive MgSO4 than non-smoking mothers. It would be interesting in future analyses to compare sociodemographic risk factors for treatment between nations.

The NPP was associated with around £597,000 net monetary benefit from a lifetime societal perspective, with 89% probability of being cost-effective accounting for babies with less than 30 weeks’ gestation.

Including babies <32 weeks’ gestation, the NPP was associated with £4.2m of net monetary benefit. These results evidence that the NPP was cost-effective. The impact could be higher as there was evidence of the effect of the NPP among babies between 32 and 34 weeks gestation. We have not sought to estimate the cost-effectiveness of the NPP for this group of patients, given the limited evidence of life-time costs associated with CP. Our estimate of the increase in MgSO4 uptake attributable to the NPP entails estimating how MgSO4 uptake may have changed over time in the absence of the NPP. Our sensitivity analysis illustrates an alternative assumption for estimating the counterfactual, which indicates a longer-term impact and consequently a larger associated NMB of £5.4m for babies with less than 32 weeks gestation.

MgSO4 implementation generates health gains and cost savings over time. The three nations have increased MgSO4 uptake over the nine years to 2022 for those babies of less than 30 weeks gestation, such that the associated net monetary benefit has increased by 67% to £76m in England, by 159% to £5.7m in Scotland, and by 329% to £3.1m in Wales. Consequently, the benefit forgone due to suboptimal MgSO4 uptake (95% uptake), has reduced in the three nations. MgSO4 uptake for babies of 30 or 31 weeks gestation has also improved, but the uptake of MgSO4 is further away from the optimal uptake, hence, there is a greater benefit forgone due to suboptimal uptake. There remains considerable scope for further investment in the implementation of MgSO4 to achieve optimal uptake across all three nations.

We have shown that investing in new QI programmes to support increasing MgSO4 uptake further are very likely to be cost-effective in the three nations. For instance, a hypothetical QI programme costing £6,000 per unit, similar in cost to NPP, achieving a 5% increment of MgSO4 uptake in a single year would generate NMB of £6m in England, £520,000 in Scotland and £266,000 in Wales. For the purpose of this analysis, we have assumed funding allocated to all units in each nation. In practice, any future implementation programmes may target specific units or networks experiencing comparatively weak performance. Our analysis supports the prioritisation of funding to increase MgSO4 uptake further in the three nations.

Our qualitative study shed light on corresponding implementation activities taking place in Scotland and Wales (Scotland’s Maternity and Children Quality Improvement Collaborative (MCQIC) leading on the implementation of the Preterm Perinatal Wellbeing Package (PPWP), and the Welsh Joint Maternity and Neonatal Strategic network roll-out of PERIPrem Cymru. The challenges and enablers raised by participants reflect those emerging from the main PReCePT evaluations, and both nations reported being influenced by PReCePT methodology. Our analysis highlighted specific challenges and enablers to driving improvement across settings, but also to sustaining improvement over time. This was a challenge for all units as reported by participants, the reason being the fact that MgSO4 was administered by maternity teams, but uptake data were recorded and audited by neonatal teams who also provided governance and leadership. These factors related to:

- Discordant conceptualisations of the intervention, and of improvement intentions between maternity and neonatology;
- Perinatal and multiprofessional teamworking and collaboration;
- Individual rather than collective concerted action to enable timely and appropriate administration of MgSO4, constrained by lack of resources (e.g. staffing issues, knowledge and competencies, integrated pathways, and access to equipment);
- Problematic data collection, audit, and feedback processes.

National policy and audit mechanisms such as the National Neonatal Audit programme were mobilising forces behind improvement strategic intentions by providing national direction on how clinical practice should look like, and providing data on actual performance allowing for benchmarking. This demonstrates that without national audit and feedback mechanisms to benchmark performance on existing clinical guidelines, it is difficult for clinical leadership to identify areas needing improvement, and for local teams to understand why improvement interventions are needed. Improvements however were driven by local initiative, and differences in improvement capacity between settings resulted in significant variations in performance, indicating how some settings might require additional support input and resource to push up improvement capability, as also indicated by PReCePT findings. Improvements were believed to be concentrated in units with strong perinatal team resource such as perinatal optimisation and improvement culture and capacity, but such improvements, typically in NICU units, did not spread across settings. Particular components such as local champions, perinatal teamworking, audit and feedback, and diverse multi-disciplinary and multi-organisational perinatal networks of participation, were believed to be important for perinatal optimisation and spreading good practice.

### 5.2 Interpretation

Uptake of new evidence on (cost-)effectiveness of clinical interventions in healthcare practice is known to be slow and may take 10 to 20 years before it reaches optimal level of implementation. During this period of suboptimal implementation, major health benefits and/or cost savings associated with the cost-effective intervention are foregone, as not all eligible patients are provided with the cost-effective intervention.

During this period, there is opportunity to accelerate the uptake of the intervention, through active implementation efforts. These efforts and associated costs may be justified by the incremental health gains generated by increased uptake, as well as offset by potential cost savings associated with the intervention. The extent to which such an implementation effort is deemed cost-effective will therefore depend both on the cost-effectiveness of the clinical intervention itself, and the cost-effectiveness of the implementation programme, i.e. how well uptake is improved in relation to the cost of the intervention.

MgSO4 has been demonstrated to be a very cost-effective intervention in threatening preterm birth to reduce the risk of neurological brain damage including cerebral palsy, and thus long-term societal costs. Therefore MgSO4 was considered likely to benefit from a national programme to accelerate implementation across maternity units across England. The various analyses throughout this report illustrate the methodological complexity and multitude of quantitative, qualitative and health economic perspectives involved in such evaluation. And the challenge to carry out robust methodology to demonstrate the additional benefit of this programme, against the background of national and local policies, professional communication, and mutual influencing amongst regions or even nations, and the diminishing gap between current and optimal implementation.

Typically, new evidence-based interventions take around 10-15 years to be fully implemented into clinical practice. Additional support for implementation may accelerate this process, thereby generating health benefits and cost savings, depending on the cost-effectiveness of the clinical intervention. A key question is what the window of opportunity is to generate health benefits and cost savings, in relation to the costs required for implementation support.

Our results provide further evidence that the NPP helped improve uptake in England, and that the benefit has been largely sustained over time. The estimate of improvement of around 5% across the four years of follow-up was robust to sensitivity analyses and multiple analytic approaches.

The original Bayesian analysis using Scotland and Wales as a synthetic control group should be interpreted with caution, due to the departure from the parallel trends assumption in the pre-intervention period. While this analysis may overestimate the effect size, in the context of the overall convergence of results within confidence limits, may indicate that the real effect is at the higher end of the confidence limits indicated by the other analyses; with up to 9% improvement.

The reduction in missing data is likely to reflect one of the PReCePT aims of improving data quality. MgSO4 was also included in the maternity clinical negligence scheme incentive in England, so if uptake was above 80%, Trusts received insurance reductions.

The economic evaluation shows clear evidence of the NPP being cost-effective. The NPP generated around £597,000 net monetary benefit through an increase of MgSO4 uptake for babies of less than 30 weeks’ gestation. When babies of 31 or 32 weeks’ gestation are included, the net monetary benefit increases to £4.2m. This benefit represents the lifetime societal impact of increasing MgSO4 uptake on health gains and cost savings associated with the prevention of CP in preterm babies associated with the additional increase MgSO4 uptake due to NPP.

Although the trends in the devolved nations are less clear due to smaller numbers (and so higher variability) it appears that they have largely caught up with England uptake; converging at high levels in all three nations. Scotland and Wales did not have a NPP, but did implement other local and national initiatives to improve MgSO4 use. We also know that hospitals in Scotland and Wales were accessing the online PReCePT resources, with 66 downloads from Wales and 32 from Scotland from 2018-2022 (AHSN data, unpublished). Although this is a small amount in comparison to the total recorded 4434 downloads, it does indicate that there may have been indirect benefits of PReCePT in the devolved nations – and this diffusion of knowledge (which would be considered ‘contamination’ in a trial) could dilute comparisons between the nations.

The observed plateau in uptake may indicate that the ‘easy’ gains (or “low hanging fruit”) have now been made, and getting the national average beyond the current high levels may be a challenge without further concerted effort directed, perhaps, at the lower-performing units. In 2022, across the three nations, 10-15% of eligible mothers were recorded as not receiving MgSO4 due to imminent delivery. We know from qualitative data that this may involve mothers not being diagnosed as being in preterm labour by obstetricians until it is too late to administer MgSO4 or transfer mothers to a larger unit. PReCePT evaluations demonstrated how improvements across the system such as training and awareness raising among teams involved in prenatal and perinatal care e.g. community midwives and staff in smaller units can improve response timings which can in turn improve adherence. Assuming the reported reasons are accurate, and the proportion of mothers with imminent deliveries is unlikely to change, it may indeed be difficult to further improve average national uptake beyond 85-90%. Current uptake levels in England, Scotland and Wales are comparable to (and at the higher end of) levels reported internationally (69% to 87%^26–29^) following guidelines and/or interventions to increase MgSO4 uptake. One the other hand, some individual units do report higher (>90%) uptake, and arguably these exceptional units demonstrate that it is possible to achieve better (potentially through better triaging and monitoring of symptoms) and so should be used as the benchmark for quality of care.

Implementing MgSO4 generates long-term health gains and cost savings associated with the prevention of brain injury including CP in preterm babies. We have used estimates of these to calculate net monetary benefit generated over time in the three nations due to the increase of MgSO4, and the benefit forgone associated with suboptimal uptake. This analysis should encourage further investment to support optimal MgSO4 implementation.

It is plausible that the COVID-19 pandemic negatively impacted MgSO4 use via its impact on staffing and overall quality of care. Mothers may also have been presenting at hospital later, due to concerns about infection, or fear of giving birth without a birthing partner; factors that could mean missed opportunities to give MgSO4. Analysis of future data will be essential to monitor this trend and hopefully pre-empt further declines in use. Concerningly, follow-up of a comparable national programme in New Zealand, also found a recent decline in use following an initially successful intervention (unpublished finding, conference presentation).

Another factor, that could potentially be impacting results, is a change in gestational age demographics due to a larger proportion of babies at extremely low (22 and 23 weeks’) gestation being given survival focused care. Numbers of the most immature babies under 24 weeks gestation admitted to neonatal units has been gradually increasing over time. As this group have lower odds of being treated, their increasing numbers could be artefactually pulling down the average uptake over time. We explored this in the data, and there was a small increase in babies up to 22 weeks gestation (92 in 2021-2022 versus 15 in 2018-19). Numbers in other gestational ages showed little difference. However, these babies made up <0.5% of the dataset, which is very unlikely to impact on results. It also seems questionable that any impact from increasing numbers in this age group this would only co-occur with the onset of the COVID-19 pandemic, which is where we do observe a slight decline in uptake. Future reporting should if possible adjust for this evolving gestational age demographic, and the influence of evolving guidance on their management. Health economic implications will also be affected by increasing numbers of survivors in these extremely preterm babies.

The suggestion of greater improvements in uptake when including older preterm births (all babies up to 34 weeks of gestation at birth) is interesting. While these infants represent a much larger proportion of preterm births than the more extreme preterm infants, interpretation is complex; their profile of underpinning antenatal disease, and hence their presentation to healthcare, may vary from the more preterm presentation, and this may impact on the subsequent ease of delivering antenatal optimisation. Indeed, thresholds for earlier delivery may be different across the gestational range, while the units, and the staff, looking after them may also be patterned by gestational age. However, evidence on the protective effect of MgSO4 in babies 30-34 weeks is unclear^3^ ^30^ ^31^, although long-term neurological impacts remain higher in these groups than term-born peers^32^, and due to their greater numbers, even small shifts in risks may have substantial population benefits^33^. Qualitative analysis showed that even though administration to babies below 30 weeks was considered routine, more ambiguity and therefore variation was described in MgSO4 administration to those over 30 weeks, with some participants reporting that babies were most often not being given it.

These results are consistent with what we have seen in those babies between 30 and 31 weeks gestation. Our cost-effectiveness results extended up to less than 32 weeks gestation show that the NPP was effective and cost-effective in increasing the uptake of MgSO4. Extending the gestational age range impacted positively on the number of babies treated, and thus the NPP was estimated as even more cost-effective.

From a health economic perspective, we have not included in the cost-effectiveness babies between 32 and 34 weeks gestation as there is not sufficient evidence of MgSO4 cost-effectiveness for this group of patients. The uptake of MgSO4 for babies between 31 and 32 weeks gestation has increased in the three nations but to lower rates in comparison to babies with lower gestation. Therefore, the NMB forgone for not implementing MgSO4 uptake is higher. We estimated that treating this group of patients to optimal uptake (i.e., 95%) could generate an additional £10m in England, £2.7m in Scotland and £651k in Wales. Therefore, we suggest revising the evidence to include more specific instructions about deliveries with babies between 31 and 32 weeks’ gestation.

History of smoking may be associated with a lower chance for not receiving MgSO4 due to unmeasured socio-demographic patterning (e.g. it is more frequent in younger white women from more deprived backgrounds, who may be more likely to have unplanned pregnancies and present at hospital later). Analysis was adjusted for available sociodemographic factors, although residual and uncontrolled confounding is still possible. Alternatively, another explanation, may be that women who smoke may spend some of their treatment window smoking, which reduces their overall opportunity to receive the drug. The decline of the historical North/South disparity in use in England is a positive finding, suggesting improvements in equality of healthcare access across the country. This is consistent with the advantages of a universal national programme, rather than separate localised efforts.

Qualitative findings demonstrated that scaling, spreading, and sustaining evidence-based interventions over time and across settings requires strong perinatal clinical leadership to provide clear guidance on what the intervention is, why it should be administered, when, and how, in a way that is meaningful to all teams and actors involved in its administration. Co-production of the narrative, communication strategy, policy, pathways, processes, and workflows is essential to ensure the “right people at the right place” are identified and engaged, implementers and adopters are clear of their and each others’ roles. Clear practical clinical guidance, documentation, and equipment (e.g. prefilled syringes, use of aide memoirs) reduce uncertainly, build confidence, and make decision-making and administration easier and quicker.

For sustaining uptake and driving improvement, national performance audit mechanisms (e.g. NNAP) are essential for making teams aware of what is happening in practice through benchmarking, but such data need to be disseminated to all perinatal stakeholders. Easy to use data capture systems and IT systems with interfacing capabilities are also needed to make data capture and audit as easy as possible. Even though performance audit data are essential drivers for improvement strategic intentions, for behaviour change to happen on the ground, explanatory as well as descriptive data are needed in the form of missed case reviews. Learning needs to be fed back to teams and individuals in meaningful ways. Local champions can facilitate and drive improvement but they need funded and protected time to engage in local implementation activities and mobilise teams, but also to participate in communities of practice and networks where capacity building, access to clinical leadership, diffusion of innovation and knowledge, and continued learning takes place. It is also clear that improvement activities are fragile and reliant on systemic forces and enablers to continue over time as well as local action. Without the necessary investment to ensure safe staffing levels, policy and leadership backing for improvement, and continuous capacity building targeting units with less improvement and safety capability, local teams struggle to divert resources from addressing immediate care needs of their patients, to improvement activity. For this reason we recommend that quality improvement activities need to be embedded in all levels of the perinatal ecosystem, rather than be the responsibility of individual clinicians or teams.

### 5.3 Strengths and limitations

This study benefitted from high quality, routinely collected, longitudinal patient-level data. Two analytical approaches (both with sensitivity analyses) were used to evaluate the impact of the NPP, in order to check and triangulate results, and counteract the individual (but different) potential bias structures of each method. Results from these multiple approaches were very similar, which strengthens the overall picture of a beneficial effect of the programme.

As data covered almost all maternity units in England, Scotland, and Wales (excepting the five in England that comprised the PReCePT pilot), this represents a highly generalisable cohort. Data covered a period of eight years, four before and four after launch of the NPP, which gives adequate time for analysis of trends and assessment of the medium-term impact and sustainability of the NPP.

A limitation is that this is by necessity a non-randomised study, and so it is not possible to conclusively attribute the observed increase in uptake to the NPP alone. In this case it was neither possible, nor arguably ethical to conduct an RCT (strongly positive results from the pilot study mean clinical equipoise was lost). We have tried to minimise the impact of confounding through analytic methods, and interpret findings with appropriate caution. The total observed increases in MgSO4 use are likely to be due to many factors.

Definitive evidence on the protective effective of the drug, from systematic review and meta-analysis, was published in 2009^34^. Use started being reliably recorded as a Neonatal Data Analysis Unit (NDAU) audit metric for maternity units in 2014-15, and became a recommendation in the NICE Guidance in 2015^2^. The PReCePT pilot study was also started in 2015, and published positive results in 2017^8^. Neonatal Audit Programme (NNAP) themselves concluded in their report on 2020 data that “This rapid improvement [in MgSO4 use], particularly seen in England, is likely to result from the targeted approach of the PReCePT quality improvement initiative”.^35^

Ability to draw comparisons between nations was limited. Although perinatal care is in many ways delivered comparably in England, Scotland, and Wales, there are also important differences that could confound comparisons (for example, Scotland and Wales have relatively more tertiary units (NICUs) than England). The smaller numbers and so higher variability in the devolved nations MgSO4 data additionally limit precise distinctions in uptake levels. As noted above, Scotland and Wales also had their own interventions to improve uptake, and staff there were also accessing PReCePT resources, which are all likely to dilute comparisons between the groups.

The COVID-19 pandemic was a significant external event affecting all aspects of healthcare. This can hamper assessment of the effects of other activities taking place in the same time period. It is hard to disentangle what might be a temporary negative effect of the pandemic, versus what might be a natural waning of the initially positive effect of the NPP. From the observation that antenatal steroid use (historically well-established at high levels) declined almost identically to MgSO4 use over the pandemic period, we suggest that the declines may be most likely associated with the pandemic, but further follow up is important to monitor whether levels of both treatments recover, and guide further intervention if not. One could also speculate that Brexit was another external factor that may have impacted on NHS workforce and thereby quality of care^36^. The NNAP data dashboard indicates that since the beginning of the pandemic the lowest neonatal nursing to patient ratios were experienced in the second half of 2022.

The total observed increases in MgSO4 use since 2014 (when records became sufficiently reliable) are likely to be due to many factors. Definitive evidence on the protective effective of the drug, from systematic review and meta-analysis, was published in 2009^34^. Use started being reliably recorded as a Neonatal Data Analysis Unit (NDAU) audit metric in 2014-15, and became a recommendation in the NICE Guidance in 2015^2^. The PReCePT pilot study in five units in South West England was also stared in 2015, and published positive results in 2017^8^. Initial discussions with unit leads, about the proposed NPP, also started in 2017. The MCQIC PPWP was also launched in 2017. All of these factors are likely to have had a role in the observed improvements. It is worth noting that the National Neonatal Audit Programme (NNAP) themselves concluded in their report on 2020 data that “This rapid improvement [in MgSO4 use], particularly seen in England, is likely to result from the targeted approach of the PReCePT quality improvement initiative.”^35^

Finally, we were limited to analysis of infants born, and admitted to a neonatal unit, and interpretation should bear this in mind. While there is little to suggest that MgSO4 influences early survival (and thereby chance of admission)^34^, we were unable to assess the number of women who received MgSO4, but then did not precede to deliver an eligible preterm infant.

Health economic analyses have the strength of being able to combine the implementation evidence (cost and implementation effects) with the evidence of MgSO4 treatment (health gains and cost savings associated with the prevention of CP). This is an innovative approach and appropriate in implementation science to assess the long-term impact of QI programmes such as the NPP. Yet, there are some limitations associated with the available evidence. As noted above, evidence on the protective effect of MgSO4 in premature babies is limited, and a wide range of costs associated with CP have been reported^37^. The evidence of the MgSO4 cost-effectiveness is limited to a small number of studies^2^ ^14–16^. In this study, we have used analysis by Bickford et al^14^, which used Danish resource data from 2006 estimated from a combination of register data, published literature, and expert opinion^6^. These CP costs were then adjusted to 2011 Canadian dollars before we converted them to pounds sterling inflated to 2019 prices^14^ ^38^. More recent estimates in Australia determined the cost of CP of $145,632 per person per annum^39^. However, these costs were estimated for the year 2018 alone and at a national level and, therefore, difficult to extrapolate to other settings such as England. The treatment cost-effectiveness analysis reported in the 2022 update of the NICE guideline^2^ also uses the lifetime cost data used by Bickford et al^14^. It is clear that UK-based and up-to-date estimates of lifetime CP impact would strengthen the evidence for the evaluation and prioritisation of effective strategies to prevent CP in the future, such as, MgSO4.

The qualitative methods components of our study provided invaluable contextual information to the quantitative and health economic data, enriching the interpretation of findings. A limitation to the qualitative data is the lack of representation of clinicians from smaller units in Scotland, whose experience may be different from larger NICUs. All participants however had current or past involvement in the design and implementation of improvement activities in their respective nations as part of national strategic clinical networks and quality improvement programmes, and/or as part of their local clinical leadership teams. They were able to provide an in-depth and expert description of implementation activities specific to MgSO4. Qualitative findings also enabled a comparison between the experiences of the devolved nations and England, and helped elucidate contextual and intervention specific drivers and enablers important for any improvement intervention attempting to improve uptake of evidence-based clinical perinatal interventions.

### 5.4 Conclusions, and implications for practice/NHS service provision

Improving clinical practice can be a slow process, shaped by forces spanning all levels of the perinatal care system including conclusive and coherent clinical guidance, audit and feedback mechanisms, clinical leadership, as well as organisation capacity, and team and individual clinician capabilities. Following evidence of improved preterm survival with antenatal steroids, it took decades for their use to become standard care for mothers at risk of preterm labour. The journey for MgSO4 has been more rapid, which is likely due, at least in part, to the benefits of dedicated national programmes. The recent suggestion of a downturn in use of both of these valuable treatments is concerning, and illustrates the fragility of complex healthcare systems and the need for quality improvement efforts to be embedded across all levels of the system.

Our findings stress the importance of improvement efforts which engage across the perinatal ecosystem for improvement to be equitably spread and sustained. Professional networks and communities speed up diffusion of innovation. Adequate resourcing, including workforce numbers and staff competencies is essential.

In view of this complexity, simple methods are likely insufficient to adequately evaluate processes, outcomes and impacts of improvement programs such as PReCePT. A mixed-method approach is essential to address clinical, health economic and qualitative aspects of such programmes. Innovative approaches are needed for appropriate modelling and exploration of the mechanisms of change, and of health and economic implications, both at the clinical and policy level.

Failure to deliver MgSO4 to eligible mothers should be considered inadequate care, and not financially sustainable for the NHS. MgSO4 as a quality metric should continue to be closely monitored, and further intervention (possibly targeting the lowest-performing units) may be warranted to achieve optimal treatment levels. The essential next step in this quality improvement journey is to quantify, in this same population, the health and societal benefits – i.e. cases of cerebral palsy prevented – resulting from the improvements we have achieved in use of MgSO4.

## Data Availability

Anonymised individual-level data for this study are from the NNRD. Our data sharing agreement with the NNRD prohibits sharing data extracts outside of the University of Bristol research team.

## Acknowledgements

This report relates to NIHR ARC West project number P367b. The evaluation reported here would not have been possible without the efforts and assistance of perinatal teams across England, Scotland and Wales; the QI coaches; the West of England AHSN (now Health Innovation West of England) the AHSN network (now the Health Innovation Network); and the parent collaborators who have helped steer the PReCePT programme of work. We would also like to acknowledge the helpful reflection and advice received from the study steering committee (Prof. Nigel Simpson, Monica Bridge, Elly Salisbury, Anna Burhouse, Tasha Swinscoe, Elisa Smit, Julie-Clare Becher, and Dr Colin Peters).

## Funding and declaration

This research was jointly funded by The Health Foundation, The Academic Health Science Network, The West of England AHSN and the National Institute for Health and Care Research (NIHR) Applied Research Collaboration (ARC West) at University Hospitals Bristol and Weston NHS Foundation Trust. The views expressed are those of the authors and not necessarily those of the NHS, the NIHR or the Department of Health.

## APPENDICES

### Appendix 1: Estimated lifetime costs and QALYs per patient associated with MgSO4 treatment (2019 prices)

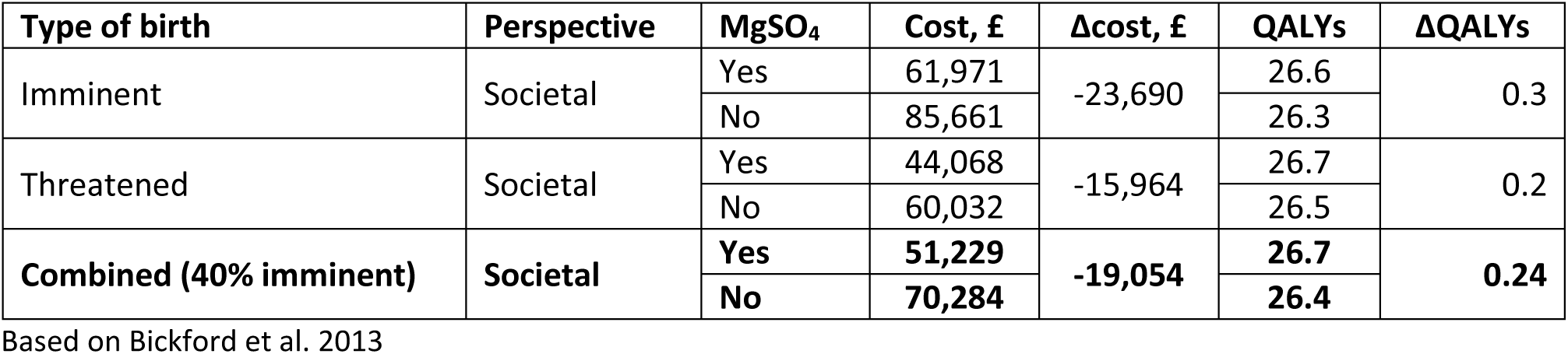

| Type of birth | Perspective | MgSO<sub>4</sub> | Cost, £ | Δcost, £ | QALYs | ΔQALYs |
| --- | --- | --- | --- | --- | --- | --- |
| Imminent | Societal | Yes | 61,971 | -23,690 | 26.6 | 0.3 |
|  |  | No | 85,661 |  | 26.3 |  |
| Threatened | Societal | Yes | 44,068 | -15,964 | 26.7 | 0.2 |
|  |  | No | 60,032 |  | 26.5 |  |
| **Combined (40% imminent)** | **Societal** | **Yes** | **51,229** | **-19,054** | **26.7** | **0.24** |
|  |  | **No** | **70,284** |  | **26.4** |  |

Based on Bickford et al. 2013

### Appendix 2: Point estimates, probability distributions, and source of parameter estimates used in the probabilistic analysis

Appendix 2: Point estimates, probability distributions, and source of parameter estimates used in the probabilistic analysis.
| Analysis | Element | Statistics | Linear | Distribution | Source |
| --- | --- | --- | --- | --- | --- |
| Common estimates | Health utility, QALYs | Mean (SE *) | 0.24 (0.05) | Beta distribution | Bickford et al. (1) |
|  | Lifetime costs, £ | Mean (SE *) | -19,054 (-3,811) | Gamma distribution | Bickford et al. (1) |
|  | Cost NPP, £ | Total | 936,747 | N/A | PReCePT Study – Cost analysis |
| Linear ITS >30 weeks gestation | Pre-term babies, *N* | Sum | 2,136 | N/A | NNRD data |
|  | Change in the proportion of MgSO<sub>4</sub> | Mean (SE) | 3.0% (0.9%) | Normal distribution | Area-Between-Curves |
| Beta ITS >30 weeks gestation | Pre-term babies, *N* | Sum | 3,129 | N/A | NNRD data |
|  | Change in the proportion of MgSO<sub>4</sub> | Mean (SE) | 3.0% (0.4%) | Normal distribution | Area-Between-Curves |
| Linear ITS >32 weeks gestation | Pre-term babies, *N* | Sum | 4,923 | N/A | NNRD data |
|  | Change in the proportion of MgSO<sub>4</sub> | Mean (SE) | 3.4% (1.0%) | Normal distribution | Area-Between-Curves |
| Beta ITS >32 weeks gestation | Pre-term babies, *N* | Sum | 7,768 | N/A | NNRD data |
|  | Change in the proportion of MgSO<sub>4</sub> | Mean (SE) | 3.4% (0.3%) | Normal distribution | Area-Between-Curves |

\*Standard errors are calculated as the 20% of the point estimates as in Bickford et al. (1)

### Appendix 3: PReCePT QI Devolved Nations Staff Interview Topic Guide

#### 1. Introductions

- Participants’ job title, time in post, role and responsibilities

#### 2. Opening

What are your thoughts on the NICE clinical guidance and its implementation in your region? Background information to your nation’s/region’s journey in introducing MgSO4 administration for neuroprotection, including your current policy?

#### 3. Commitment to the implementation of the NICE guidance

- Who is responsible for implementing this clinical guidance?
- Do you think there has been buy in and commitment by all stakeholders to implementing the guidance in your region? Why/why not?
- How are they engaging with clinical managers/perinatal teams/PPI groups?
- Is there still support from individual staff/management/the organisation/system for MgSO4 neuroprotection?

#### 4. What has taken place: implementation activities

- What strategies were used to implement the NICE guidance? On a national/regional/policy level? On a unit level?
- Where do you think you are now on your nation’s implementation trajectory/journey?
- Who does what and when in implementing these strategies and ensure adherence to the guidance
- What factors may have been responsible for increases/decreases/variation in uptake?
- Has the implementation of the guidance been supported through actions or other tangible support by management and other stakeholders e.g. policy, money, material resources
- What has been put in place to make it possible or easier for people on the ground to administer MgSO4

#### 5. Reflecting on implementation

- Overall, how would you assess the success of the implementation of this clinical guidance?
- Has your organisation been evaluating the implementation of the guidance?
- Did these result in any changes in your strategies/practice? How?
- Can you tell us about your reflections, observations, views on sustainability and learning for future national adoption and spread programmes?

#### 6. Closing

- Thank you for taking part in this interview. Is there anything else you would like to tell me for the evaluation?

### Appendix 4: The Normalisation Process Theory implementation mechanisms and how they overlap with the four primary drivers of PReCePT

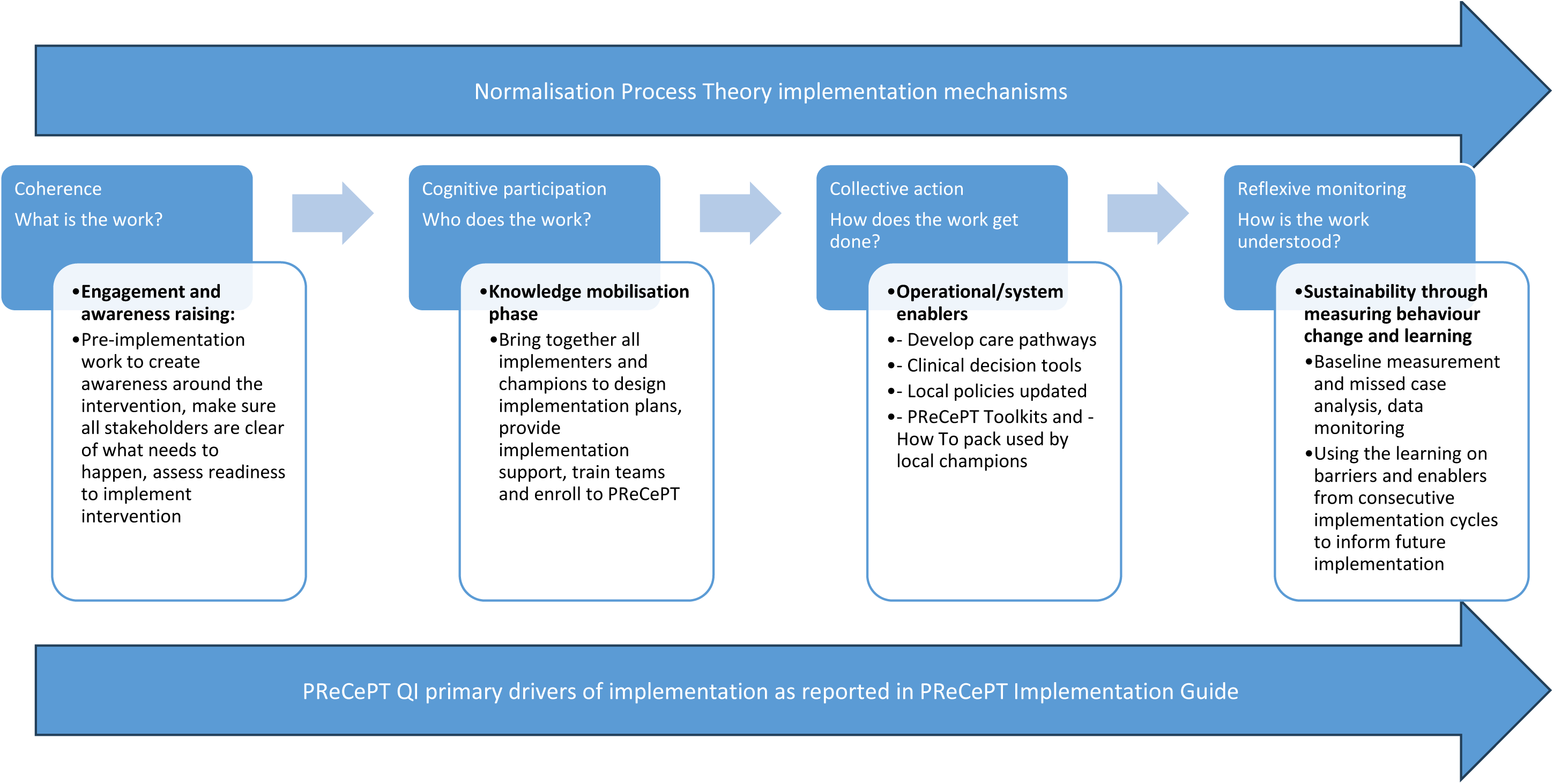

## Notes

### Competing Interest Statement

The authors have declared no competing interest.

### Funding Statement

This study was jointly funded by The Health Foundation (funders reference 557668), the National Institute for Health and Care Research Applied Research Collaboration West (NIHR ARC West, core NIHR infrastructure funded: NIHR200181), and Health Innovation West of England (formerly the West of England Academic Health Science Network). The views expressed are those of the authors and not necessarily those of NHS England, NHS Improvement, the NIHR or the Department of Health and Social Care.

### Author Declarations

The PReCePT Programme Evaluation was granted a favourable ethical opinion by the UK National Health Service Health Research Authority (HRA project ID: 260504) and the University of Bristol Faculty of Health Sciences Research Ethics Committee (FREC Ref: 84582).

